# Efficacy and safety of GLP-1 receptor agonists versus SGLT-2 inhibitors in overweight/obese patients with or without diabetes mellitus: a systematic review and network meta-analysis

**DOI:** 10.1101/2022.02.06.22270446

**Authors:** Hong Ma, Yu-Hao Lin, Li-Zhen Dai, Chen-Shi Lin, Yanling Huang, Shu-Yuan Liu

## Abstract

**Objective:** To compare the efficacy and safety between and within glucagon-like peptide-1 receptor agonists (GLP-1RAs) and sodium-glucose co-transporter 2 inhibitors (SGLT-2is) in overweight or obese adults with or without diabetes mellitus.

**Methods:** PubMed, ISI Web of Science, Embase and Cochrane Central Register of Controlled Trials (CENTRAL) database were comprehensively searched to identify randomised controlled trials (RCTs) of effects of GLP-1RAs and SGLT-2is in overweight or obese participants from inception to Jan 16, 2022. The efficacy outcomes were the changes of body weight, glucose level and blood pressure. The safety outcomes were serious adverse events and discontinuation due to adverse events. The mean differences (MDs), odds ratios (ORs), 95% credible intervals (95% CI), the surface under the cumulative ranking (SUCRA) were evaluated for each outcome by network meta-analysis.

**Results:** Sixty-one RCTs were included in our analysis. Both GLP-1RAs and SGLT-2is conferred greater extents in body weight reduction, achieving at least 5% weight loss, HbA1c and fasting plasma glucose decrease compared with placebo. GLP-1RAs was superior to SGLT-2is in HbA1c reduction (MD: -0.39%, 95% CI: -0.70 to -0.08). GLP-1RAs had high risk of adverse events, while SGLT-2is were relatively safe. Based on intraclass comparison, semaglutide 2.4mg was among the most effective interventions in losing body weight (MD: -11.51kg, 95% CI: -12.83 to -10.21), decreasing HbA1c (MD: -1.49%, 95% CI: -2.07 to -0.92) and fasting plasma glucose (MD: -2.15mmol/L, 95% CI: -2.83 to -1.59), reducing systolic blood pressure (MD: -4.89mmHg, 95% CI: -6.04 to -3.71) and diastolic blood pressure (MD: -1.59mmHg, 95% CI: -2.37 to -0.86) with moderate certainty evidences, while it was associated with high risk of adverse events.

**Conclusions:** Semaglutide 2.4mg showed greatest effects on losing body weight, controlling glycemic level and reducing blood pressure while it was associated with high risk of adverse events.

**ARTICLE SUMMARY:** 

**Strengths and limitations of this study:**

▸ The RoB 2.0 and GRADE methods were used to assess the risk of bias and evaluate the confidence in effect estimates.
▸ Study selection, data extraction, quality of study and evidence assessment were conducted independently by two authors and audited by the third author.
▸ The network meta-analysis was mostly based on indirect comparisons due to limited reported comparisons in available literatures.
▸ The small number of randomised controlled trials available in some comparisons.

## INTRODUCTION

The World Health Organization (WHO) claimed that obesity has become a major public health challenge of the 21st century.^[1]^ Obesity can cause considerable harm to affect individuals notable, including physical disabilities, psychological problems, increasing the risk of expanding sets of chronic diseases, such as diabetes mellitus, cardiovascular diseases (CVD), certain cancers and depression.^[2 3]^

Glucagon-like peptide-1 receptor agonists (GLP-1RAs) and Sodium-glucose co-transporter 2 inhibitors (SGLT-2is) are both novel glucose-lowering agents. These two categories of drugs can not only improve glycemic control, but also reduce body weight and decrease the risk of cardiovascular disease by different mechanisms.

As we know, both GLP-1RAs and SGLT-2is are recommend as medication options in type 2 diabetes mellitus patients with overweight or obesity by the American Diabetes Association (ADA). Further, the US Food and Drug Administration (FDA) has approved two kinds of GLP-1RAs: liraglutide 3.0mg and semaglutide 2.4mg for obese or overweight individuals with at least one weight-related comorbidity.^[4]^ Recently, there are many randomised controlled trials (RCTs) comparing GLP-1RAs or SGLT-2is with placebo in overweight/obese patients with or without diabetes. But, the direct comparisons between GLP-1RAs and SGLT-2is is few. In the absence of direct ‘head-to-head’ data, network meta-analysis is considered as the methodology of choice to obtain a relatively scientific evaluation of the results.^[5]^

Thus, we conducted a systematic review and network meta-analysis to estimate the difference of the efficacy and safety between and within GLP-1RAs and SGLT-2is treatments in overweight or obesity patients with or without diabetes by synthesizing the direct and indirect data of RCTs. Different GLP-1RAs and SGLT-2is and different dosage of liraglutide and semaglutide were separately analyzed including exenatide 10ug, dulaglutide 1.5mg, liraglutide 1.8mg, liraglutide 3.0mg, semaglutide 1.0mg, semaglutide 2.4mg, canagliflozin 300mg, dapagliflozin 10mg and empagliflozin 10mg to explore for potential differences among individual GLP-1RAs and SGLT-2is. The aim of this study was to provide useful insights to support clinical decision-making.

## METHODS

This systematic review and network meta-analysis had been registered on PROSPERO (No. CRD42021258103). The methods and results of this review in accordance with the Preferred Reporting Items for Systematic Reviews and Meta-Analyses for Network Meta-Analyses.^[6]^

### Data sources and search strategy

A comprehensive search of PubMed, ISI Web of Science, Embase and Cochrane Central Register of Controlled Trials (CENTRAL) was conducted in each database from inception to Jan 16, 2022 for RCTs in any language. The search strategy designed by an experienced librarian used Medical SubjectHeadings and relevant text words that consisted of terms relating to “Glucagon-Like Peptide 1”, “Sodium-Glucose Transporter 2 Inhibitors”, “obesity” and “overweight”. Full search strategy is presented in online Supplemental Table 1. In addition, other relevantly published and unpublished trials were conducted by manual search.

### Study selection and eligibility criteria

Two authors independently selected the studies that followed the PICO (population, intervention, comparator, outcome) framework, with any discrepancies resolved through a third author. The ‘liberal accelerated’ method was used to duplicate screening, which was first applied by Khangura *et al*.^[7]^ Studies were deemed eligible if they: (a) design of RCTs; (b) participants were overweight (BMI ≥25 kg/m^2^, Asian ≥23 kg/m^2^) or obesity (BMI ≥30 kg/m^2^) adults (≥18 years) ; (c) interventions were long-acting, short-acting GLP-1RAs or SGLT-2is; (d) compared intervention(s) with placebo or GLP-1RAs or SGLT-2is; (e) outcomes considered were at least one of the following: changes in body weight, proportion of participants reaching at least 5% weight loss, hemoglobin A1c, fasting plasma glucose (FPG), systolic blood pressure (SBP), diastolic blood pressure (DBP), serious adverse events (SAE) or discontinuation due to adverse events. Further, the RCTs with the results presented on ClinicalTrials.gov were also included. Studies were excluded if they: (a) patients with renal transplant, undergoing bariatric surgery, clozapine-and olanzapine-treated schizophrenia overweight; (b) recruited single gender populations (e.g., entirely females); (c) duration of interventions less than 12 weeks; (d) medications that have been withdrawn from the market. Pre-defined screening and extraction forms were piloted to ensure inter-rater reliability. And the inter-rater agreement calculated by the Cohen’s K coefficient was 0.81(95% CI: 0.44 to 1.18).

### Data extraction

Two authors independently extracted data using a standardized pre-defined spreadsheet from each included study; discrepancies were resolved by a third author. The following parameters were extracted: first author name; year of publication; clinicaltrials.gov trial number or UMIN-CTR search clinical number; intervention(s) and comparator(s); duration of follow-up; baseline characteristics of participants and background therapy. The data of number of participants, mean differences (MDs), standard deviation were extracted for continuous outcomes, total number of participants; numbers of participants with events were extracted for dichotomous outcomes. If standard deviation data were not reported, the SD was calculated as SD=SE×SQRT (n), where SE was standard error and n was the total number of participants; if the SE does not hold, the following formula was applied to compute the SD: SD=SQRT (n) ×(upper 95%CI -lower 95%CI)/3.92) (CI means credible intervals).^[8 9]^ For trials assessing more than one dose of dulaglutide, only data for the dosage of 1.5mg were included. If the same population were involved in more than one published studies, only the primary study was considered.

### Quality of evidence

The quality of evidence for the network meta-analysis was evaluated based on the GRADE (The Grading of Recommendations Assessment, Development, and Evaluation) approach.^[10]^ In this approach, the evidences were rated as high, moderate, low, or very low levels. Rating of direct estimates was based on risk of bias, inconsistency (or heterogeneity), indirectness, imprecision and publication bias. For each indirect estimate, the lower rating from the two direct estimates forming the dominant first order loop provided the starting point for certainty ratings, with further downward rating for imprecision or intransitivity. If evidence of direct and indirect estimates were coherent, the higher of their ratings were as the best estimate of the treatment effect. If direct and indirect estimates were incoherent, the evidences of network estimates were downgraded and the higher of their ratings was assigned to treatment effect estimates.

### Risk of bias

The Cochrane Collaboration Risk of Bias 2.0 tool was used to assess the risk of bias of individual studies in regard to the following aspects: randomization process, intended interventions, missing outcome data, measurement of the outcome and selection of the reported result.^[11]^ Publication bias was assessed with comparison-adjusted funnel plots.^[12]^ Any discrepancies were resolved by a third author.

### Statistical analysis

In this study, we calculated MDs for continuous outcomes and odds ratios (ORs) for dichotomous outcomes, along with 95% credible intervals (95% CI) for efficacy and safety outcomes. Direct pairwise meta-analysis for treatment comparisons was performed using DerSimonian-Laird random-effects model.^[13]^ Heterogeneity was assessed with the *I*^2^ and *I*^2^>50% indicated substantial heterogeneity.^[14]^ Based on Markov-chain Monte Carlo method, we performed random-effects Bayesian network meta-analyses to analyze direct and indirect comparisons of different treatments.^[15]^ And all treatment contrasts were assumed to have the same heterogeneity variance. The ranking probabilities of all treatments on efficacy and safety outcomes were displayed on their surface under the cumulative ranking (SUCRA) probabilities. The larger the SUCRA value is, the better is the rank of the treatment for outcomes.^[16]^ In each closed loop, inconsistency of the model was assessed by the node splitting method. Sensitivity analysis was based on trials with low risk of bias to assess the robustness of the findings. And subgroup analysis was performed for studies that participants with or without diabetes. All two-sided *p*-values of <0.05 was considered statistically significant. Statistical analyses were conducted using R with “gemtc” and “meta” packages, Stata 16 MP were used for comparison-adjusted funnel plots.

### Patient and public involvement

Patients were not involved in this design or conduct of this systematic review and network meta-analysis.

## RESULTS

We identified 12825 trials using the search strategy, and 7346 duplicates were eliminated. After 5479 trials were screened for titles and abstracts, 936 trials were received for full-text screening. Finally, 61 RCTs with 17281 participants were included for the analyses. Details of trials selection were shown in Figure 1. The available networks of evidence were displayed in Figure 2. In 61 RCTs, 60 RCTs were parallel design, except 1 RCT was across-over design; and 56 RCTs were blinded design, remaining 5 RCTs were open-label design. The participants of 35 RCTs were reported in overweight/obese patients with diabetes, whereas 20 RCTs were conducted in overweight/obese patients without diabetes, remaining 6 RCTs performed in overweight/obese patients with or without diabetes. The duration of all RCTs ranged from 12 weeks to 72 weeks, with the median of 24 weeks (interquartile range [IQR] 16 weeks to 36 weeks).

**Figure 1.**
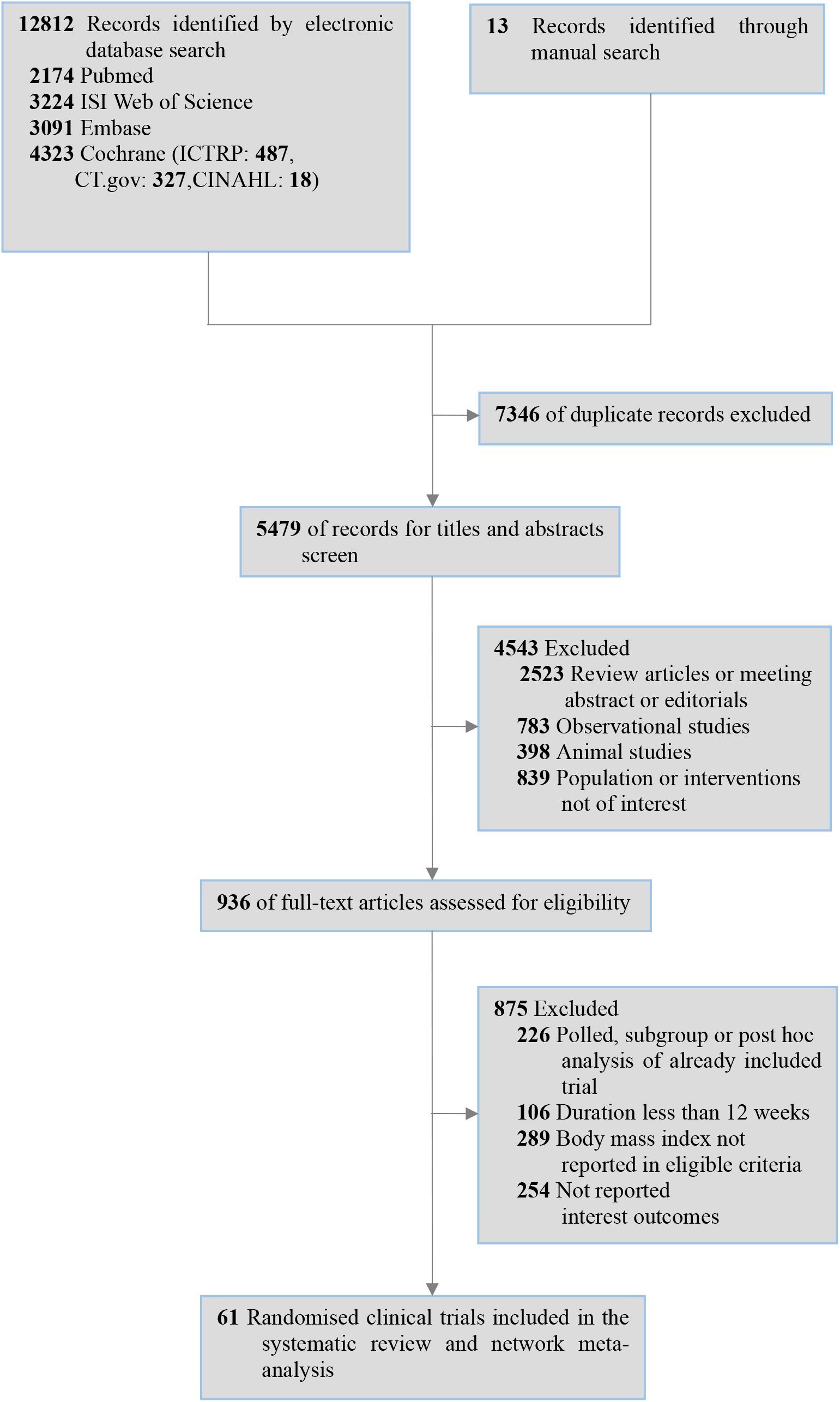
Summary of study search and selection.

**Figure 2.**
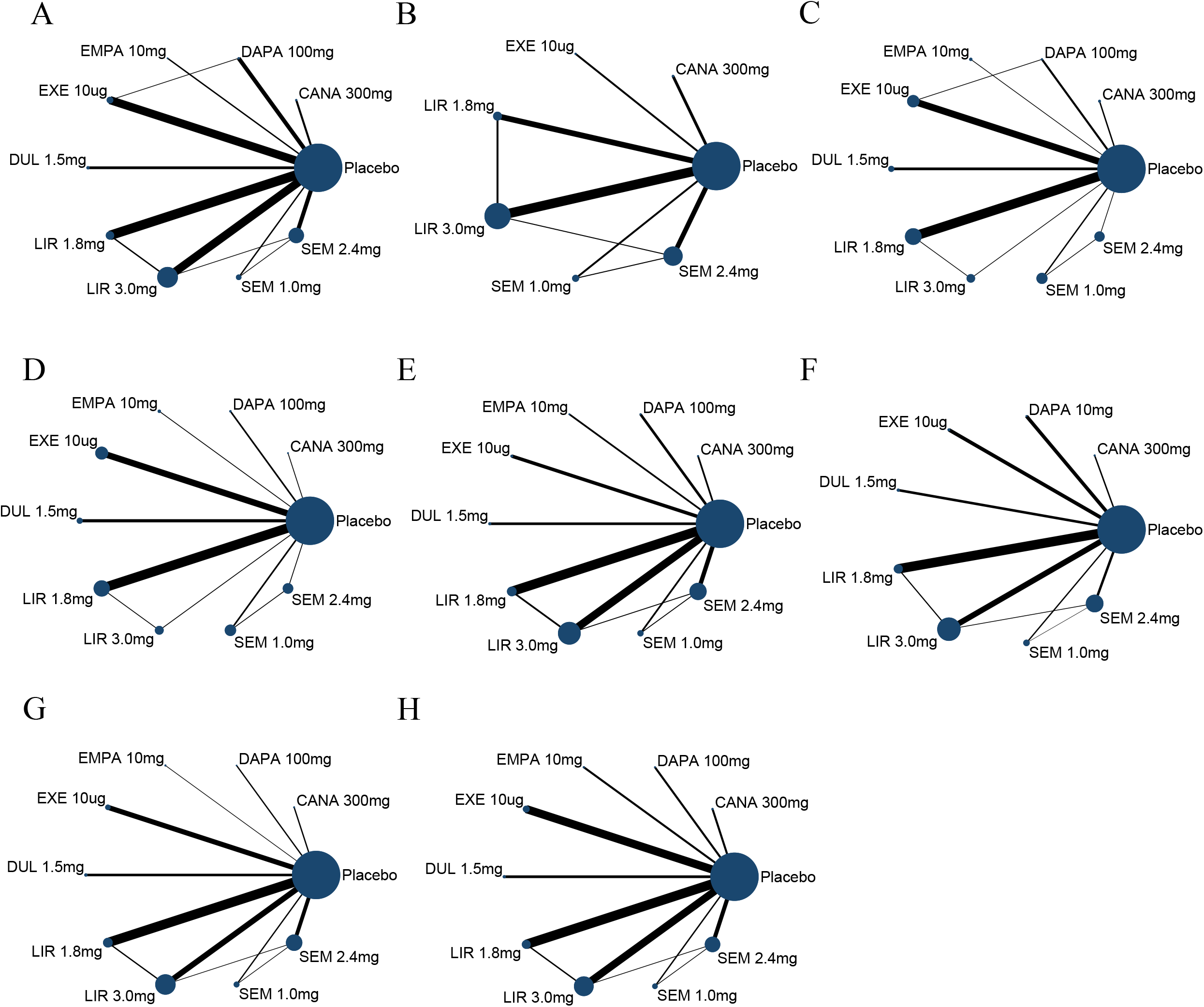
The network diagrams of all eligible comparisons for the primary outcomes of efficacy and safety. (A) Body weight; (B) Achieving ≥5% weight loss; (C) Systolic blood pressure; (D) Diastolic blood pressure; (E) HbA1c; (F) Fasting plasma glucose; (G) Serious adverse events; (H) Discontinuation due to adverse events. CANA, canagliflozin; DAPA, dapagliflozin; EMPA, empagliflozin; Exe, exenatide; DUL, dulaglutide; LIR, liraglutide; SEM, semaglutide.

The median of average age of patients across RCTs was 54.1 years (range, 24.0-68.0 years). Patients’ median of average BMI was 33.6 kg/m^2^ (range, 27.3-40.1 kg/m^2^), median of average body weight was 96.4 kg (range, 68.0-118.7 kg). The mean duration of diabetes was 8.72 years (SD 4.93 years), mean of baseline HbA1c was 7.92 % (SD 0.54 %) in participants with diabetes. In most RCTs, participants received lifestyle modification, including hypocaloric diet and regular physical activity. The characteristics of included RCTs were summarized in online Supplemental Table 2.

### Certainty of evidence and risk of bias

Among 61 RCTs, 47 RCTs with low risk, 2 RCTs with high risk, and 12 RCTs were judged to have some concerns in overall risk of bias (online Supplemental Table 3). The study “Schiavon, 2021” was classified as high risk of bias due to that it was judged to have some concerns for domains of “Bias arising from the randomization process” and have high risk for domains of “Bias due to deviations from intended interventions” that substantially lowered confidence in the result in a way. The study “Matikainen, 2018” was classified as high risk of bias due to that it was judged to have some concerns for domains of “Bias due to deviations from intended interventions” and “Bias due to deviations from intended interventions” in a way that substantially lowered confidence in the result. The quality of evidence was low to moderate in most comparisons and the confidence downgraded mainly due to study limitation, inconsistency, indirectness and imprecision (online Supplemental Table 4 and Supplemental Table 5). Further, comparison-adjusted funnel plots were relatively symmetric and did not suggest the presence of publication bias (online Supplemental Figure 1).

### Body weight

With respect to the change in body weight, 56 RCTs were analyzed. In direct pairwise meta-analyses, compared with placebo, reductions in body weight were observed in GLP-1RAs and SGLT-2is, range from -1.47kg to -11.47kg (online Supplemental Figure 2). Results of network meta-analysis demonstrated that both GLP-1RAs and SGLT-2is had effects on body weight reduction, and these two agents conferred similar body weight reductions (online Supplemental Table 6). Compared to placebo, semaglutide 2.4mg (MD: -11.51kg, 95% CI: -12.83 to -10.21, SUCRA: 0.99) showed greatest reduction in body weight, followed by semaglutide 1.0mg (MD: -5.67kg, 95% CI: -7.84 to -3.52, SUCRA: 0.86), liraglutide 3.0mg (MD: -4.65kg, 95% CI: -5.60 to - 3.69, SUCRA: 0.79). Liraglutide 1.8mg, exenatide 10ug, dulaglutide 1.5mg and dapagliflozin 10mg reduced body weight from -3.14kg to -1.28kg. Canagliflozin 300mg and empagliflozin 10mg had modest effects in body weight reduction.

Compared with other medications, semaglutide 2.4mg showed greater benefits in body weight, ranged from -10.23kg to -5.84kg (Figure 3, online Supplemental Figure 3). In the subgroup analysis of trials with diabetic patients, compared with placebo, all treatments had greater effects on body weight reduction and semaglutide 2.4mg showed greatest reduction in body weight. In the subgroup analysis of trials with non-diabetic patients, dulaglutide 1.5mg and semaglutide 1.0mg were not analyzed due to limited RCTs. And dapagliflozin 10mg and exenatide 10ug had modest effects on body weight reduction compared with placebo (online Supplemental Table 7).

**Figure 3.**
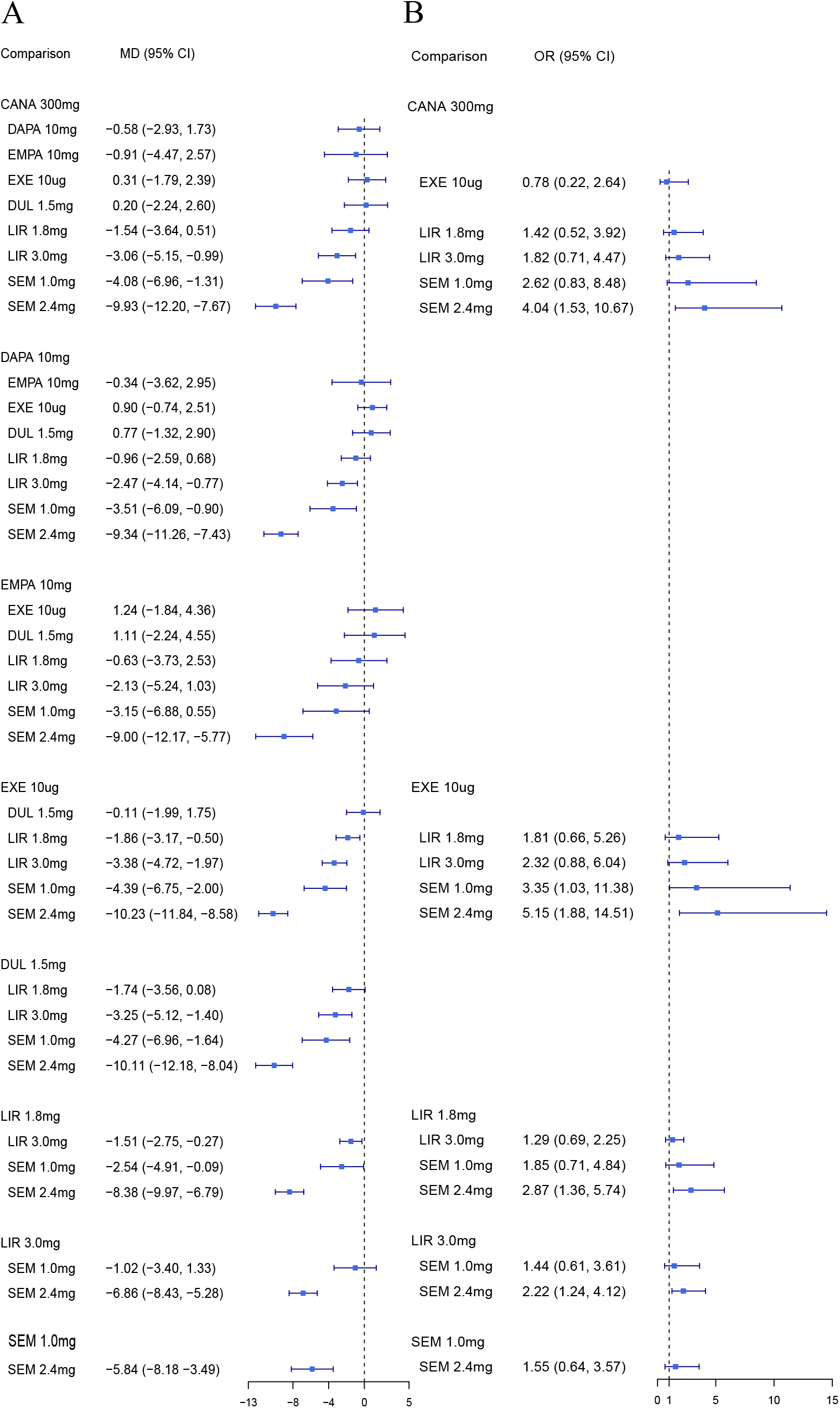
Network meta-analysis results among active treatment. (A) body weight (kg); (B) achieving ≥ 5% weight loss (%). MD, mean difference; OR, odds ratio; 95% CI, 95% credible interval; CANA, canagliflozin; DAPA, dapagliflozin; EMPA, empagliflozin; Exe, exenatide; DUL, dulaglutide; LIR, liraglutide; SEM, semaglutide.

### Reaching ≥ 5% weight loss

With respect to reaching at least 5% weight loss, 24 RCTs were analyzed. Due to limited RCTs, outcome of reaching at least 5% weight loss were not analyzed in dulaglutide 1.5mg, dapagliflozin 10mg and empagliflozin 10mg. Pairwise meta-analysis results are presented in online Supplemental Figure 2. Results of network meta-analyses demonstrated that compared with placebo, both GLP-1RAs and SGLT-2is was associated with higher odds of reaching at least 5% weight loss. While no notable differences were evident between GLP-1RAs and SGLT-2is. Compared to placebo, semaglutide 2.4mg (OR: 10.88, 95% CI: 6.69 to 18.39, SUCRA: 0.97) was associated with the highest odds in reaching weight reductions of at least 5%, followed by semaglutide 1.0mg (OR: 7.02, 95% CI: 3.25 to 16.50, SUCRA: 0.80), liraglutide 3.0mg (OR: 4.90, 95% CI: 3.37 to 7.13, SUCRA: 0.64), liraglutide 1.8mg (OR: 3.81, 95% CI: 2.31 to 6.70, SUCRA: 0.49), canagliflozin 300mg (OR: 2.70, 95% CI: 1.19 to 6.40, SUCRA: 0.51), no difference was found in exenatide 10ug. Based on intraclass comparisons, semaglutide 2.4mg was superior to liraglutide 3.0mg, liraglutide 1.8mg, canagliflozin 300mg and exenatide 10ug; semaglutide 1.0mg was superior to exenatide 10ug (Figure 3, online Supplemental Figure 3). In the subgroup analysis of patients with diabetes, compared with placebo, only liraglutide 1.8mg and semaglutide 1.0mg were associated with higher odds in reaching weight reduction of at least 5%. In the subgroup analysis of non-diabetic patients, due to limited RCTs, the analysis was not conducted in semaglutide 1.0mg. And canagliflozin 300mg no longer associated with higher odds in reaching weight reduction of at least 5% compared with placebo (online Supplemental Table 7).

### Glycemic control

Outcomes of HbA1c were analyzed in diabetic patients with 35 RCTs. Direct pairwise meta-analyses showed that GLP-1RAs and SGLT-2is were associated with reduction in HbA1c compared with placebo (online Supplemental Figure 2). Network meta-analysis results showed that both GLP-1RAs and SGLT-2is were associated with greater extent in HbA1c reduction compared with placebo. Further, GLP-1RAs was superior to SGLT-2is (MD: -0.39%, 95% CI: -0.70 to -0.08) in HbA1c reduction (online Supplemental Table 6). Compared with placebo, semaglutide 2.4mg showed the greatest reduction in HbA1c (MD: -1.49%, 95% CI: -2.07 to -0.92, SUCRA: 0.93), followed by semaglutide 1.0mg (MD: -1.38%, 95% CI: -1.83 to -0.96, SUCRA: 0.89) and dulaglutide 1.5mg (MD: -1.11%, 95% CI: -1.44 to -0.79, SUCRA: 0.75).

Liraglutide 3.0mg, exenatide 10ug, liraglutide 1.8mg, canagliflozin 300mg and dapagliflozin 10mg were conferred HbA1c reductions ranging from -0.98% to -0.48% (Figure 4, online Supplemental Figure 2). The outcomes of fasting plasma glucose were also analyzed in diabetic patients with 30 RCTs. Pairwise meta-analysis results were presented in online Supplemental Figure 2. Network meta-analysis results showed that both GLP-1RAs and SGLT-2is showed greater efficacy in fasting plasma glucose reduction compared with placebo. While no notable differences were found between GLP-1RAs and SGLT-2is. The most efficacy in a mean fasting plasma glucose reduction was semaglutide 2.4mg (MD: -2.15mmol/L, 95% CI: -2.83 to -1.59, SUCRA: 0.92), followed by semaglutide 1.0mg (MD: -2.01mmol/L, 95% CI: -2.67 to -1.54, SUCRA: 0.84) and liraglutide 3.0mg (MD: -1.86mmol/L, 95% CI: -2.45 to - 1.26, SUCRA: 0.75) compared with placebo. Dulaglutide 1.5mg, liraglutide 1.8mg, exenatide 10ug, canagliflozin 300mg, dapagliflozin 10mg and empagliflozin 10mg were conferred fasting plasma glucose reductions ranging from -1.79mmol/L to - 0.81mmol/L (Figure 4, online Supplemental Figure 3).

**Figure 4.**
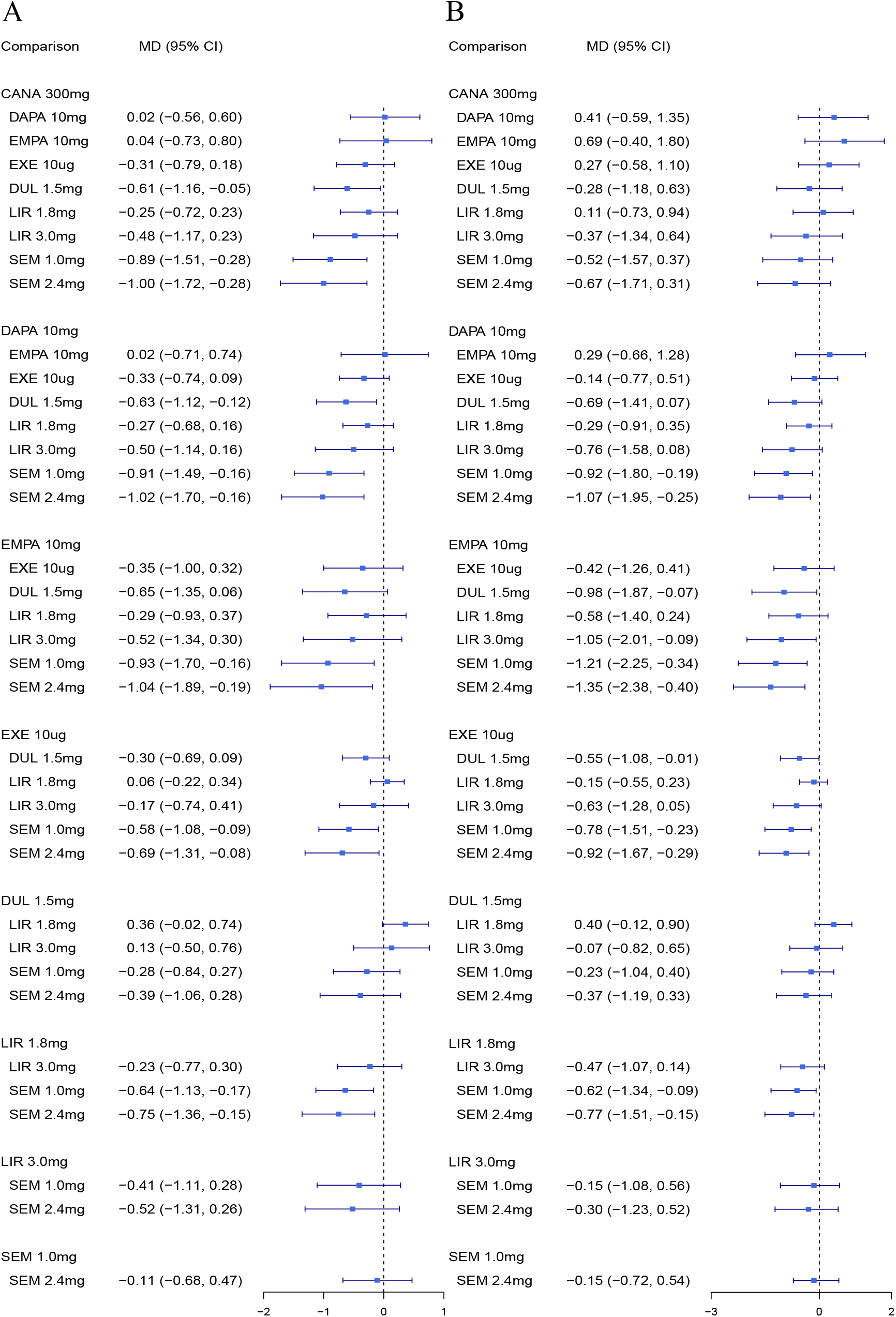
Network meta-analysis results among active treatment. (A) HbA1c (%); (B) Fasting plasma glucose (mmol/L). MD, mean difference; 95% CI, 95% credible interval; CANA, canagliflozin; DAPA, dapagliflozin; EMPA, empagliflozin; Exe, exenatide; DUL, dulaglutide; LIR, liraglutide; SEM, semaglutide.

### Blood pressure

#### Systolic blood pressure

Thirty-seven RCTs were included in the analysis of systolic blood pressure. Pairwise meta-analysis results were presented in online Supplemental Figure 2. In network meta- analysis, GLP-1RAs had significant effects on systolic blood pressure decrease, while SGLT-2is had modest effects. Compared with placebo, semaglutide 2.4mg (MD: -4.89mmHg, 95% CI: -6.04 to -3.71, SUCRA: 0.96), exenatide 10ug (MD: - 3.86mmHg, 95% CI: -6.29 to -1.47, SUCRA: 0.78) and semaglutide 1.0mg (MD: - 3.25mmHg, 95% CI: -5.46 to -1.04, SUCRA: 0.68) ranked as the top three in systolic blood pressure reduction, followed by liraglutide 1.8mg and liraglutide 3.0mg. However, dulaglutide 1.5mg, canagliflozin 300mg, dapagliflozin 300mg and empagliflozin 10mg had a neutral effect. Further, based on intraclass comparison, semaglutide 2.4mg was more efficacious than liraglutide 3.0mg, liraglutide 1.8mg, dulaglutide 1.5mg, canagliflozin 300mg and dapagliflozin 10mg in reducing systolic blood pressure, while no significant differences were found between other individual GLP-1RAs and SGLT-2is (Figure 5, online Supplemental Figure 3). In the subgroup analysis of trials with diabetic patients, canagliflozin 300mg were not analyzed due to limited RCTs. Compared with placebo, exenatide 10ug, liraglutide 1.8mg and semaglutide 2.4mg showed greater reduction in systolic blood pressure (online Supplemental Table 7).

**Figure 5.**
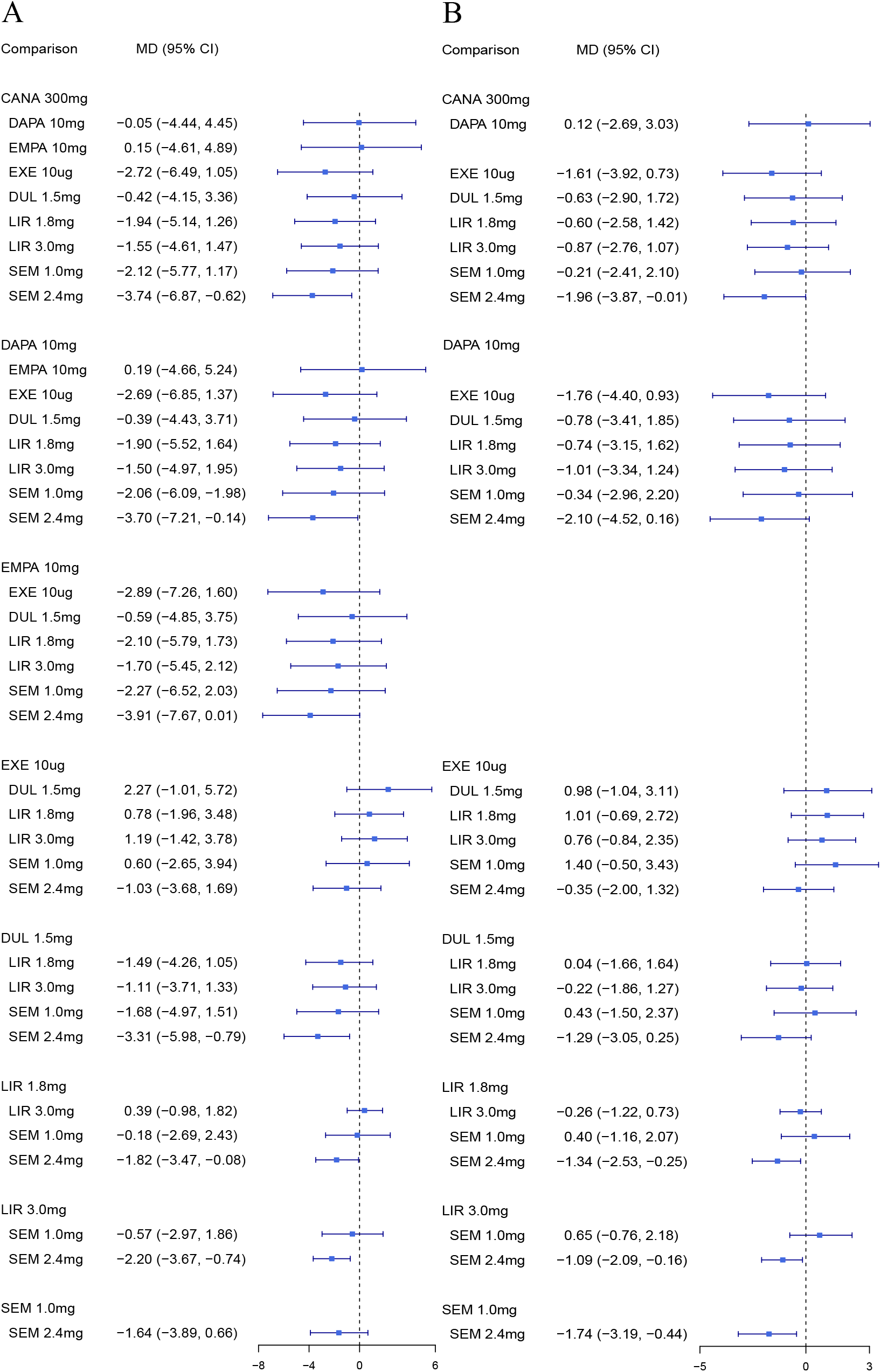
Network meta-analysis results among active treatment. (A) Systolic blood pressure (mmHg); (B) Diastolic blood pressure (mmHg). MD, mean difference; 95% CI, 95% credible interval; CANA, canagliflozin; DAPA, dapagliflozin; EMPA, empagliflozin; Exe, exenatide; DUL, dulaglutide; LIR, liraglutide; SEM, semaglutide.

#### Diastolic blood pressure

Thirty-four RCTs were included in the analysis of diastolic blood pressure. Data on diastolic blood pressure of empagliflozin 10mg was not available because of limited RCTs. Pairwise meta-analysis results were presented in online Supplemental Figure 2. In network meta-analysis, GLP-1RAs significantly reduced diastolic blood pressure in comparison with placebo, while SGLT-2is had neutral effects in diastolic blood pressure reduction. In addition, semaglutide 2.4mg (MD: -1.59mmHg, 95% CI: -2.37 to -0.86) had greater extent in diastolic blood pressure reduction, while SGLT-2is and other kinds of GLP-1RAs had neutral effects compared with placebo. Further, based on intraclass comparison, semaglutide 2.4mg was more effective than liraglutide 3.0mg, liraglutide 1.8mg, semaglutide 1.0mg and dapagliflozin 10mg in diastolic blood pressure reduction, while no significant differences were found between other individual GLP-1RAs and SGLT-2is (Figure 5, online Supplemental Figure 3). In the subgroup analysis of patients with diabetes, due to limited RCTs, the analysis was not conducted in canagliflozin 300mg. And semaglutide 2.4mg no longer reduced diastolic blood pressure compared with placebo (online Supplemental Table 7).

### Adverse event outcomes

#### Serious adverse events

Pairwise meta-analysis results showed higher odds of serious adverse events for semaglutide 2.4mg and liraglutide 3.0mg compared with placebo (online Supplemental Figure 2). In network meta-analysis, compared with placebo, GLP-1RAs was associated with higher risk of serious adverse events, while SGLT-2is had no significant difference. Semaglutide 2.4mg (OR: 1.42, 95% CI: 1.02 to 1.99) increased the risk of serious adverse events in comparison with placebo. Whereas, no differences of serious adverse events were found in other kinds of GLP-1Ras and SGLT-2is (Figure 6, online Supplemental Figure 3). The subgroup result of diabetic patients showed that semaglutide 2.4mg was no longer associated with higher risk of serious adverse events (online Supplemental Table 7). The most frequent serious adverse events with SGLT-2is were genital mycotic infection and urinary tract infection; while the most frequent serious adverse events with GLP-1RAs were pancreatitis and acute gallbladder disease.

**Figure 6.**
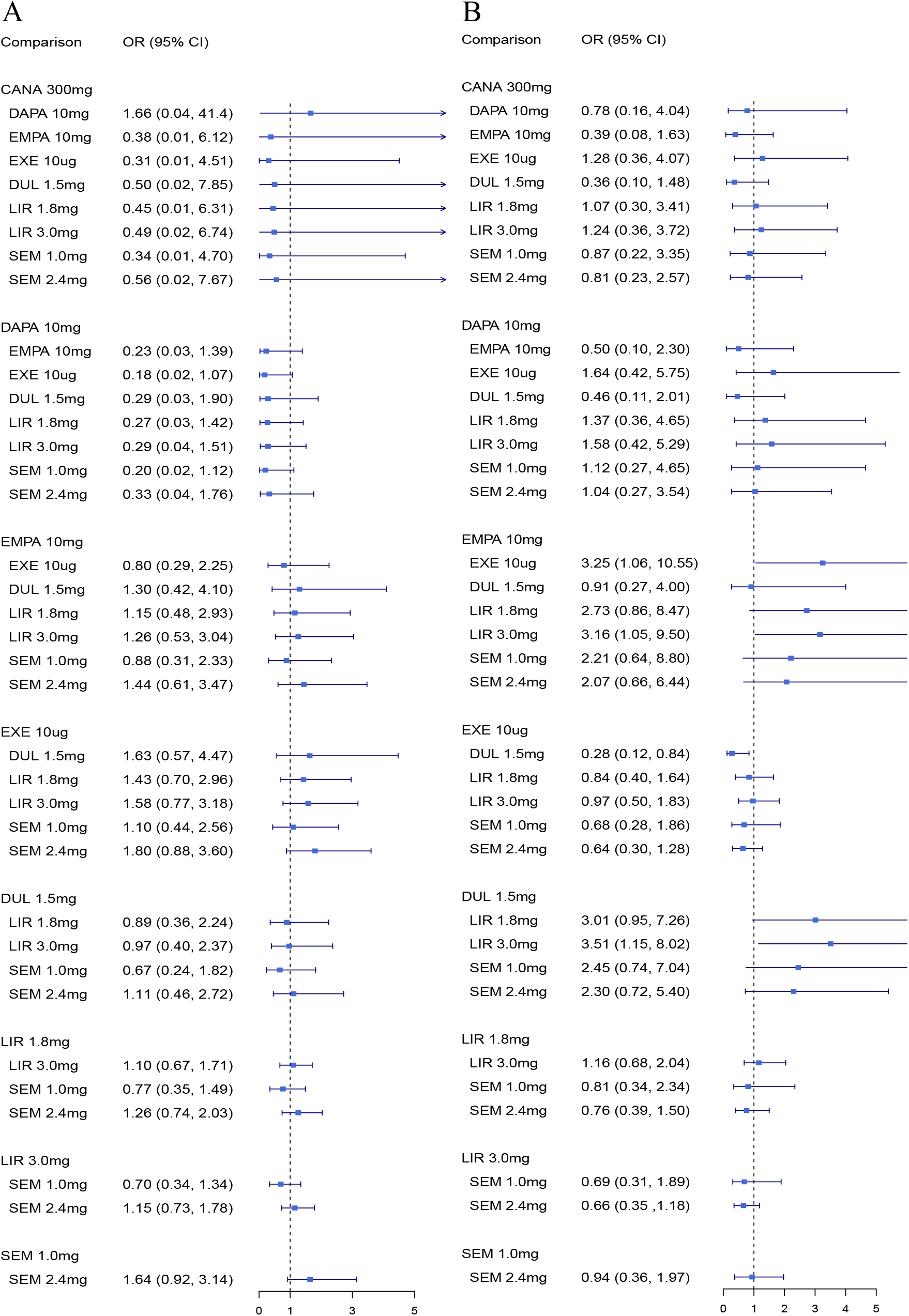
Network meta-analysis results among active treatment. (A) Serious adverse evnets; (B) Discontinuation due to adverse events. OR, odds ratio; 95% CI, 95% confidence interval; CANA, canagliflozin; DAPA, dapagliflozin; EMPA, empagliflozin; Exe, exenatide; DUL, dulaglutide; LIR, liraglutide; SEM, semaglutide.

#### Discontinuation due to adverse events

Pairwise meta-analysis results showed that compared with placebo, the majority of GLP-1RAs had higher odds of discontinuation due to adverse events, but not for dulaglutide 1.5mg (online Supplemental Figure 2). In network meta-analysis, compared with placebo, GLP-1RAs was associated with higher risk of discontinuation due to adverse events, while SGLT-2is had no significant difference. In addition, exenatide 10ug (OR: 2.96, 95% CI: 1.79 to 4.99), liraglutide 3.0mg (OR: 2.88, 95% CI: 1.90 to 4.24), semaglutide 2.4mg (OR: 1.88, 95% CI: 1.12 to 2.99) was ranked top three in the risk of discontinuation due to adverse events, while no differences were found in dulaglutide 1.5mg, semaglutide 1.0mg, canagliflozin 300mg, dapagliflozin 10mg and empagliflozin 10mg (Figure 6, online Supplemental Figure 3). The subgroup result of diabetic patients showed that only exenatide 10ug was associated with higher risk of discontinuation due to adverse events (online Supplemental Table 7). Most participants discontinued treatment due to adverse events mainly because of genital mycotic infection and urinary tract infection in the SGLT-2is group, and mainly because of gastrointestinal adverse events in the GLP-1RAs group.

### Sensitive analysis

Sensitivity analysis was performed to assess the body weight reduction that restricted to RCTs at low risk of bias. In sensitivity analysis, compared with placebo, most results were consistent with outcomes of main analysis. Compared with placebo, exenatide 10ug showed greater reduced body weight in the main analysis, while in the sensitive analysis exenatide 10ug had neutral effects in body weight reduction (online Supplemental Table 8).

## DISCUSSION

Both GLP-1RAs and SGLT-2is were antidiabetic agents; they also had potent effects on body weight reduction. Further, liraglutide 3.0mg and semaglutide 2.4mg was approved for obese or overweight individuals with at least one weight-related comorbidity by FDA.^[4]^ Thus, we conducted a network meta-analysis in overweight or obesity patients with or without diabetes to provide evidences of efficacy and safety difference in GLP-1RAs (including exenatide 10ug, liraglutide 1.8mg, liraglutide 3.0mg, semaglutide 1.0mg, semaglutide 2.4mg and dulaglutide 1.5mg) and SGLT-2is (including canagliflozin 300mg, dapagliflozin 10mg and empagliflozin 10mg).

Results of our network meta-analyses demonstrated that GLP-1RAs as a class played the roles in losing body weight, controlling glycemic levels and reducing blood pressure, while it increased the risk of adverse events. SGLT-2is as a class that showed benefits in reduction of body weight, HbA1c and FPG. Based on intraclass comparisons, our findings provided moderate certainty evidences that semaglutide 2.4mg was among the most effective interventions to reduce body weight, HbA1c, FPG, SBP and DBP. With respect to serious adverse events (SAE), semaglutide 2.4mg increased the risk of SAE with moderate certainty evidence. Liraglutide 1.8mg, liraglutide 3.0mg and semaglutide 2.4mg increased the risk of discontinuation due to adverse events with moderate certainty evidences.

Obesity is associated with a state of chronic inflammation that induces insulin resistance, which is considered as the first step in the development of type 2 diabetes. Weight loss above 5 kg is associated with improved diabetes comorbidities and can help 34%-70% of participants achieved diabetes remission.^[17 18]^ In our network meta-analysis, semaglutide 2.4mg and semaglutide 1.0mg were associated with weight reduction more than 5kg, while the result of semaglutide 1.0mg was with low certainty evidence. These results were consistent with the study conducted by Shi *et al*, which suggested that semaglutide showed greater weight loss than other weight-lowering drugs.^[19]^ All kinds of GLP-1RAs reduce the body weight by lowing energy intake, increasing the satiety and slowing the release of food from the stomach.^[20 21]^ But semaglutide had greater reduction in caloric intake than other GLP-1RAs and it was also associated with reduction in food cravings, which was not found in other GLP-1RAs.^[22 23]^

In our network meta-analysis, most interventions included were associated with HbA1c reduction, except for empagliflozin 10mg. Our findings provided moderate certainty evidences that semaglutide 2.4mg and dulaglutide 1.5mg were among the most effective interventions in the HbA1c reduction. In addition, a study of Hussein *et al* compared the efficacy and tolerability of GLP-1RAs and SGLT-2is, which indicated that dulaglutide and semaglutide showed greater reduction in HbA1c compared with other treatments.^[24]^ Semaglutide 2.4mg and dulaglutide 1.5mg demonstrated greater glucose-lowering effects suggesting that they provided high pharmacological levels of exogenous GLP-1 RA, which had great effects on β-cell function improvement.^[25 26]^ The analysis also found that all treatments showed greater extent in FPG reduction and semaglutide 2.4mg was among the most effective intervention compared with placebo with moderate certainty evidence.

The effects of GLP-1RAs and SGLT-2is in overweight or obese patients with or without diabetes on blood pressure were analyzed. Results demonstrated that semaglutide 2.4mg, exenatide 10ug, semaglutide 1.0mg, liraglutide 1.8mg and liraglutide 3.0mg were conferred systolic blood pressure decrease ranging from - 4.89mmHg to -2.68mmHg; only semaglutide 2.4mg was associated with diastolic blood pressure decrease of 1.61mmHg. A decrease of at least 5 mmHg in systolic blood pressure or 3 mmHg in diastolic blood pressure was defined as clinically meaningful reduction, which can reduce the risk of major cardiovascular events.^[27]^ Therefore, in our study, none of interventions met this threshold. A study conducted by Palmer *et al*. evaluated efficacy of glucose-lowering medications on blood pressure in type 2 diabetes.^[28]^ And all included interventions in this study did not meet this blood pressure threshold either.

Recently, some RCTs demonstrated that GLP-1RAs and SGLT-2is were associated with reduction of all-cause mortality, cardiovascular and renal events in diabetic patients complicated with/at high risk of atherosclerotic cardiovascular disease (ASCVD), which usually represented late stages of the disease. It is well known that obesity is strongly and continuously associated with ASCVD and renal disease in the adults.^[29]^ Decreasing body weight can reduce cardiometabolic risk factors and improve health-related quality of life in overweight or obese patients.^[30 31]^ Despite the reducing of body weight was intermediate outcome, it can also provide particularly useful treatment decisions for patients at low cardiovascular risk. It is important for clinicians to select appropriate drugs for patients with early stage of disease in clinical practice. Therefore, the participants included in our study were overweight/obese patients with or without diabetes mellitus and the efficacy and safety of GLP-1RAs and SGLT-2is were analyzed.

Since the different side effects in GLP-1RAs and SGLT-2is, serious adverse events and discontinuation due to adverse events were considered to assess the safety of these drugs. Results of our network analysis indicated that GLP-1RAs was associated with high risk of serious adverse events and discontinuation due to adverse events, SGLT-2is had better tolerability. Semaglutide 2.4mg conferred the highest risk of serious adverse events. In addition, exenatide 10ug, liraglutide 3.0mg and semaglutide 2.4mg were ranked top three in the risk of discontinuation due to adverse events. The higher risk of serious adverse events and discontinuation due to adverse events might be due to the high dose of the agents.

There are several strengths in our network meta-analysis. We utilized a comprehensive literature search, stringent inclusion and exclusion criteria to identify eligible studies, used RoB 2.0 and GRADE method to assess the risk of bias and evaluate confidence in effect estimates. Further, we included RCTs with different dosages of liraglutide and semaglutide, and compared the distinction among them. Moreover, not only was the difference between GLP-1RAs and SGLT-2is analyzed, but also intraclass differences among individuals of GLP-1RAs and SGLT-2is were illustrated. Certain limitations should be acknowledged. First, due to the small number of studies available in direct comparisons between active agents, the network meta-analysis was mostly based on indirect comparisons. Second, one of the inherent limitations of all meta-analyses is that the presence of a certain degree of clinical heterogeneity in terms of various factors-i.e., patient baseline characteristics, cointerventions, background therapy and outcome assessment. Third, with the outcome of achieving at least 5% weight loss, our analysis found that semaglutide 2.4mg had similar effects compared with semaglutide 1.0mg. This finding was different from the result of RCT study STEP 2^[32]^ due to limited “head-to-head” data in our analysis. Another limitation is that in some comparison, small numbers of RCTs were included in the analysis.

In clinical practice, individualized choice of optimal treatments such as efficacy, safety, medication adherence, treatment costs and medication accessibility should be considered. GLP-1RAs and SGLT-2is were more expensive than other anti-diabetic agents. Future trials are needed to evaluate the cost-effective and medication adherence between GLP-1RAs and SGLT-2is in overweight or obese patients with or without diabetes.

## CONCLUSION

In conclusion, both GLP-1RAs and SGLT-2is showed great reduction in body weight, HbA1c and fasting plasma glucose in overweight or obese patients with or without diabetes. Across individual agents, semaglutide 2.4mg showed greatest effects on losing body weight, controlling glycemic levels and reducing blood pressure, while it was associated with the highest risk of adverse events. SGLT-2is had modest effects on body weight and glucose level, but they had better tolerability. This network meta-analysis provided evidence of efficacy and safety differences of GLP-1RAs and SGLT-2is in overweight or obese adults with or without diabetes to guide the clinical practice.

## Data Availability

Data are available in a public, open access repository. All data relevant to the study are included in the article or uploaded as supplemental information.

## Acknowledgements

We would like to thank Natural Science Foundation of Fujian Province, China for funding this work.

## Author contributions

Two authors (HM and YH L) formulated the search strategy, selected studies for inclusion, extracted the data for outcomes, analyzed the data with R and drafted the manuscript. HM was responsible for the revision of the manuscript. LZ D was responsible for selected studies for inclusion and extracted the data for outcomes. CS L was responsible for the solution of any discrepancies. YH and SY L were responsible for the oversight of study.

## Patient consent for publication

Not required.

## Ethical approval and informed consent

Since this is a systematic review, ethical approval and informed consent are not required.

## Provenance and peer review

Not commissioned; externally peer reviewed.

## Data availability statement

No additional data are available.

## Open access

This is an open access article distributed in accordance with the Creative Commons Attribution Non Commercial (CC BY-NC4.0) license, which permits others to distribute, remix, adapt, build upon this work non-commercially, and license their derivative works on different terms, provided the original work is properly cited, appropriate credit is given, any changes made indicated, and the use is non-commercial. See: http://creativecommons.org/licenses/by-nc/4.0/.

