## supplement for "Efficacy and safety of GLP-1 receptor agonists versus SGLT-2 inhibitors in overweight/obese patients with or without diabetes mellitus: a systematic review and network meta-analysis"

### Supplement Table 1. Full search strategy.

**Pubmed**

| # | Searches | Results |
| --- | --- | --- |
| 1 | (Sodium-Glucose Transporter 2 Inhibitors[MeSH Terms]) OR (Canagliflozin[MeSH Terms]) | **3,617** |
| 2 | (((((((((((((((((((((((((((((((Sodium-Glucose Transporter 2 Inhibitors[Title/Abstract])) OR (Sodium Glucose Transporter 2 Inhibitors[Title/Abstract])) OR (Sodium-Glucose Transporter 2 Inhibitor[Title/Abstract])) OR (Sodium Glucose Transporter 2 Inhibitor[Title/Abstract])) OR (SGLT-2 Inhibitors[Title/Abstract])) OR (SGLT 2 Inhibitors[Title/Abstract])) OR (Gliflozins[Title/Abstract])) OR (SGLT2 Inhibitors[Title/Abstract])) OR (Gliflozin[Title/Abstract])) OR (SGLT-2 Inhibitor[Title/Abstract])) OR (Inhibitor, SGLT-2[Title/Abstract])) OR (SGLT 2 Inhibitor[Title/Abstract])) OR (SGLT2 Inhibitor[Title/Abstract])) OR (Inhibitor, SGLT2[Title/Abstract])) OR (Canagliflozin[Title/Abstract])) OR (Invokana[Title/Abstract])) OR (Canagliflozin Hemihydrate[Title/Abstract])) OR (Canagliflozin, Anhydrous[Title/Abstract])) OR (empagliflozin[Title/Abstract])) OR (Jardiance[Title/Abstract])) OR (dapagliflozin[Title/Abstract])) OR (Forxiga[Title/Abstract])) OR (ipragliflozin[Title/Abstract])) OR (Suglat[Title/Abstract])) OR (luseogliflozin[Title/Abstract])) OR (Lusefi[Title/Abstract])) OR (tofogliflozin[Title/Abstract])) OR (tofogliflozin hydrate[Title/Abstract])) OR (Apleway[Title/Abstract])) OR (Deberza[Title/Abstract])) OR (sergliflozin[Title/Abstract]) OR (remogliflozin etabonate[Title/Abstract]) OR (ertugliflozin[Title/Abstract]) OR (Steglatro[Title/Abstract) OR (bexagliflozin[Title/Abstract]) OR (henagliflozin[Title/Abstract]) | **6,258** |
| 3 | #1 OR #2 | **6,965** |
| 4 | (((Glucagon-Like Peptide 1[MeSH Terms]) OR (Liraglutide[MeSH Terms])) OR (exenatide[MeSH Terms])) | **11,169** |
| 5 | (((((((((((((((((((((((((Glucagon-Like Peptide 1[Title/Abstract]) OR (Glucagon Like Peptide 1[Title/Abstract])) OR (GLP-1[Title/Abstract])) OR (GLP 1[Title/Abstract])) OR (Glucagon-Like Peptide-1[Title/Abstract])) OR (Liraglutide[Title/Abstract])) OR (Victoza[Title/Abstract])) OR (Saxenda[Title/Abstract])) OR (exenatide[Title/Abstract])) OR (Bydureon[Title/Abstract])) OR (Exendin-4[Title/Abstract])) OR (Ex4 Peptide[Title/Abstract])) OR (Peptide, Ex4[Title/Abstract])) OR (Exendin 4[Title/Abstract])) OR (Byetta[Title/Abstract])) OR (dulaglutide[Title/Abstract])) OR (Trulicity[Title/Abstract])) OR (lixisenatide[Title/Abstract])) OR (Adlyxin[Title/Abstract])) OR (Lyxumia[Title/Abstract])) OR (albiglutide[Title/Abstract])) OR (Eperzan[Title/Abstract])) OR (Tanzeum[Title/Abstract])) OR (semaglutide[Title/Abstract])) OR (Ozempic[Title/Abstract])) | **19,128** |
| 6 | #4 OR #5 | **20,176** |
| 7 | #3 OR #6 | **26,507** |
| 8 | (((obesity[MeSH Terms]) OR (overweight[MeSH Terms])) OR (weight loss[MeSH Terms])) | **265,706** |
| 9 | ((((((((((((((((((obes*[Title/Abstract])) OR (body mass ind*[Title/Abstract])) OR (adipos*[Title/Abstract])) OR (overweight[Title/Abstract])) OR (overload syndrom*[Title/Abstract])) OR (overeat*[Title/Abstract])) OR (over eat*[Title/Abstract])) OR (overfeed*[Title/Abstract])) OR (over feed*[Title/Abstract])) OR (overfed[Title/Abstract])) OR (over fed[Title/Abstract])) OR (weight cycling[Title/Abstract])) OR (skinfold thickness[Title/Abstract])) OR (antiobesity[Title/Abstract])) OR (anti-obesity[Title/Abstract])) OR (obesitas[Title/Abstract])) OR (bodyweight[Title/Abstract])) OR (body weight[Title/Abstract]) | **742,566** |
| 10 | #8 OR #9 | **796,505** |
| 11 | randomized controlled trial[Publication Type] OR randomized[Title/Abstract] OR placebo[Title/Abstract] | **914,606** |
| 12 | #7 AND #10 AND #11 | **2,174** |

**ISI Web of Science**

| **#** | Searches | Results |
| --- | --- | --- |
| 1 | TS=(Sodium-Glucose Transporter 2 Inhibitor* OR Sodium Glucose Transporter 2 Inhibitor* OR Sodium-Glucose Transporter 2 Inhibitor OR SGLT-2 Inhibitor* OR SGLT 2 Inhibitor* OR Gliflozins OR Gliflozin OR Inhibitor, SGLT-2 OR SGLT2 Inhibitor OR Inhibitor, SGLT2 OR Canagliflozin OR Invokana OR Canagliflozin Hemihydrate OR Canagliflozin, Anhydrous OR empagliflozin OR Jardiance OR dapagliflozin OR Forxiga OR ipragliflozin OR Suglat OR luseogliflozin OR Lusefi OR tofogliflozin OR tofogliflozin hydrate OR Apleway OR Deberza OR sergliflozin OR remogliflozin etabonate OR ertugliflozin OR Steglatro OR sotagliflozin OR bexagliflozin OR henagliflozin) | **10,231** |
| 2 | TS=(Glucagon-Like Peptide 1 OR Glucagon Like Peptide 1 OR GLP-1 OR GLP 1 OR Glucagon-Like Peptide-1 OR Liraglutide OR Victoza OR Saxenda OR exenatide OR Bydureon OR Exendin-4 OR Ex4 Peptide OR Peptide, Ex4 OR Exendin 4 OR Byetta OR dulaglutide OR Trulicity OR lixisenatide OR Adlyxin OR Lyxumia OR albiglutide OR Eperzan OR Tanzeum OR semaglutide OR Ozempic) | **31,903** |
| 3 | #1 OR #2 | **40,697** |
| 4 | TS=(obes* OR body mass ind* OR adipos* OR overweight OR overload syndrom* OR overeat* OR over eat* OR overfeed* OR over feed* OR overfed OR over fed OR weight cycling OR skinfold thickness OR antiobesity OR anti-obesity OR obesitas OR weight loss OR bodyweight OR body weight) | **1,351,371** |
| 5 | TS=(random* controlled trial OR random* OR placebo) | **2,185,652** |
| 6 | #3 AND #4 AND #5 | **3,224** |

**Embase**

| # | Searches | Results |
| --- | --- | --- |
| 1 | 'sodium glucose cotransporter 2 inhibitor'/exp | **15,483** |
| 2 | 'sodium-glucose transporter 2 inhibitors':ab,ti OR 'sodium-glucose transporter 2 inhibitor':ab,ti OR 'sglt-2 inhibitors':ab,ti OR 'gliflozins':ab,ti OR 'sglt2 inhibitors':ab,ti OR 'gliflozin':ab,ti OR 'sglt-2 inhibitor':ab,ti OR 'inhibitor, sglt-2':ab,ti OR 'sglt2 inhibitor':ab,ti OR 'inhibitor, sglt2':ab,ti OR 'canagliflozin':ab,ti OR 'invokana':ab,ti OR 'canagliflozin hemihydrate':ab,ti OR 'empagliflozin':ab,ti OR 'jardiance':ab,ti OR 'dapagliflozin':ab,ti OR 'forxiga':ab,ti OR 'ipragliflozin':ab,ti OR 'suglat':ab,ti OR 'luseogliflozin':ab,ti OR 'lusefi':ab,ti OR 'tofogliflozin':ab,ti OR 'apleway':ab,ti OR 'deberza':ab,ti OR 'sergliflozin':ab,ti OR 'remogliflozin etabonate':ab,ti OR 'ertugliflozin':ab,ti OR 'steglatro':ab,ti OR 'bexagliflozin':ab,ti OR 'henagliflozin':ab,ti | **10,637** |
| 3 | #1 OR #2 | **16,124** |
| 4 | 'glucagon like peptide 1'/exp OR 'liraglutide'/exp OR 'exendin 4'/exp | **32,980** |
| 5 | 'glucagon like peptide 1':ab,ti OR 'glp-1':ab,ti OR 'liraglutide':ab,ti OR 'victoza':ab,ti OR 'saxenda':ab,ti OR 'exenatide':ab,ti OR 'bydureon':ab,ti OR 'ex4 peptide':ab,ti OR 'exendin 4':ab,ti OR 'byetta':ab,ti OR 'dulaglutide':ab,ti OR 'trulicity':ab,ti OR 'lixisenatide':ab,ti OR 'adlyxin':ab,ti OR 'lyxumia':ab,ti OR 'albiglutide':ab,ti OR 'eperzan':ab,ti OR 'tanzeum':ab,ti OR 'semaglutide':ab,ti OR 'ozempic':ab,ti OR 'taspoglutide':ab,ti | **30,532** |
| 6 | #4 OR #5 | **39,519** |
| 7 | #3 OR #6 | **52,089** |
| 8 | 'obesity'/exp OR 'body weight loss'/exp | **721,604** |
| 9 | 'obes*':ab,ti OR 'body mass ind*':ab,ti OR 'adipos*':ab,ti OR 'overweight':ab,ti OR 'overload syndrom*':ab,ti OR 'overeat*':ab,ti OR 'over eat*':ab,ti OR 'overfeed*':ab,ti OR 'over feed*':ab,ti OR 'overfed':ab,ti OR 'over fed':ab,ti OR 'weight cycling':ab,ti OR 'skinfold thickness':ab,ti OR 'antiobesity':ab,ti OR 'anti- obesity':ab,ti OR 'obesitas':ab,ti OR 'bodyweight':ab,ti OR 'body weight':ab,ti | **1,039,980** |
| 10 | #8 OR #9 | **1,291,067** |
| 11 | 'random':ab,ti OR 'placebo':ab,ti OR 'double-blind':ab,ti | **753,077** |
| 12 | #7 AND # 10 AND #11 | **3,091** |

Cochrane CENTRAL

| # | Searches | Results |
| --- | --- | --- |
| 1 | MeSH descriptor: [Sodium-Glucose Transporter 2 Inhibitors] | **382** |
| 2 | MeSH descriptor: [Canagliflozin] | **226** |
| 3 | (sodium-glucose transporter 2 inhibitors):ti,ab,kw OR (SGLT2 Inhibitors):ab,ti OR (SGLT-2 Inhibitors):ti,ab,kw OR (SGLT 2 Inhibitors):ti,ab,kw OR (gliflozins):ti,ab,kw OR (invokana):ti,ab,kw OR (canagliflozin hemihydrate):ti,ab,kw OR (Canagliflozin, Anhydrous):ti,ab,kw OR (empagliflozin):ti,ab,kw OR (jardiance):ti,ab,kw OR (dapagliflozin):ti,ab,kw OR (forxiga):ti,ab,kw OR (ipragliflozin):ti,ab,kw OR (suglat):ti,ab,kw OR (luseogliflozin):ti,ab,kw OR (lusefi):ti,ab,kw OR (tofogliflozin):ti,ab,kw OR (apleway):ti,ab,kw OR (deberza):ti,ab,kw OR (tofogliflozin anhydrous):ti,ab,kw OR (tofogliflozin hydrate):ti,ab,kw OR (sergliflozin):ti,ab,kw OR (remogliflozin etabonate):ti,ab,kw OR (ertugliflozin):ti,ab,kw OR (steglatro):ti,ab,kw OR (bexagliflozin):ti,ab,kw OR (henagliflozin):ti,ab,kw | **3,430** |
| 4 | #1 OR #2 OR #3 | **3,779** |
| 5 | MeSH descriptor: [Glucagon-Like Peptide 1] | **1,855** |
| 6 | MeSH descriptor: [Liraglutide] | **742** |
| 7 | MeSH descriptor: [Exenatide] | **566** |
| 8 | (glucagon like peptide 1):ti,ab,kw OR (glp-1):ti,ab,kw OR (liraglutide):ti,ab,kw OR (victoza):ti,ab,kw OR (saxenda):ti,ab,kw OR (exenatide):ti,ab,kw OR (bydureon):ti,ab,kw OR (ex4 peptide):ti,ab,kw OR (exendin 4):ti,ab,kw OR (byetta):ti,ab,kw OR (dulaglutide):ti,ab,kw OR (trulicity):ti,ab,kw OR (lixisenatide):ti,ab,kw OR (adlyxin):ti,ab,kw OR (lyxumia):ti,ab,kw OR (albiglutide):ti,ab,kw OR (eperzan):ti,ab,kw OR (tanzeum):ti,ab,kw OR (semaglutide):ti,ab,kw OR (ozempic):ti,ab,kw OR (taspoglutide):ti,ab,kw | **7,048** |
| 9 | #5 OR #6 OR #7 OR #8 | **7,479** |
| 10 | #4 OR #9 | **8,356** |
| 11 | MeSH descriptor: [Obesity] | **14,930** |
| 12 | MeSH descriptor: [Overweight] | **17,729** |
| 13 | MeSH descriptor: [Weight Loss] | **6,783** |
| 14 | (obes*):ti,ab,kw OR (body mass ind*):ti,ab,kw OR (adipos*):ti,ab,kw OR (overweight):ti,ab,kw OR (overload syndrom*):ti,ab,kw OR (overeat*):ti,ab,kw OR (over eat*):ti,ab,kw OR (overfeed*):ti,ab,kw OR (over feed*):ti,ab,kw OR (overfed):ti,ab,kw OR (over fed):ti,ab,kw OR (weight cycling):ti,ab,kw OR (skinfold thickness):ti,ab,kw OR (antiobesity):ti,ab,kw OR (anti- obesity):ti,ab,kw OR (obesitas):ti,ab,kw OR (bodyweight):ti,ab,kw OR (body weight):ti,ab,kw | **137,435** |
| 15 | #11 OR #12 OR #13 OR #14 | **139,690** |
| 16 | (randomized controlled trial):ti,ab,kw OR (randomized):ti,ab,kw OR (placebo):ti,ab,kw | **1,082,302** |
| 17 | #10 AND # 15 AND #16 | **4,323** |

### Supplement Table 2. Characteristics of studies and patients’ baseline characteristics.

| Study and year  (trial registration number) | Duration,weeks | Drug | Randomised patients, No | Age, mean (SD), years | Weight, mean (SD), kg | BMI, mean  (SD), kg/m2 | Diabetes duration, mean(SD),years | HbA1c, mean  (SD), % | FPG, mean  (SD), years | diet | exercise | background therapy |
| --- | --- | --- | --- | --- | --- | --- | --- | --- | --- | --- | --- | --- |
| Bays2014^1^  (NCT00650806) | 12 | Canagliflozin 300mg | 96 | 43.5 (11.0) | 100.2  (18.0) | 36.9 (5.3) | 0 | NA | NA | 600-kcal/d deficit | mild to moderate activity | hypertensive medication and/or lipid-lowering medication |
|  |  | Placebo | 89 | 45.1 (11.9) | 102.2  (19.9) | 36.6 (5.5) | 0 | NA | NA |  |  |  |
| Bolinder2012^2^  (NCT00855166) | 24 | Dapagliflozin 10mg | 89 | 60.6 (8.2) | 92.1  (14.1) | 32.1 (3.9) | 6.0  (4.5) | 7.19 (0.44) | 8.2 (1.4) | diet counseling | exercise counseling | metformin |
|  |  | Placebo | 91 | 60.8 (6.9) | 90.9  (13.7) | 31.7 (3.9) | 5.5  (5.3) | 7.16 (0.53) | 8.3 (1.4) |  |  |  |
| Eriksson2018^3^  (NCT02279407) | 12 | Dapagliflozin 10mg | 19 | 65.0 (6.5) | 90.2  (8.7) | 30.5 (2.8) | 6.7  (6.0) | 7.38 (0.56) | 8.99 (1.85) | NA | NA | metformin and/or SUs |
|  |  | Placebo | 19 | 65.6 (6.1) | 93.0  (12.2) | 30.3 (3.1) | 6.5  (4.2) | 7.44 (0.80) | 9.40 (1.65) |  |  |  |
| González-  Ortiz2017^4^ | 12 | Dapagliflozin 10mg | 13 | 46.5 (5.2) | 68.0  (4.6) | 27.3 (2.0) | 0 | NA | 5.23 (0.54) | maintain their individualized prestudy diet | maintain their individualized prestudy exercise | NA |
|  |  | Placebo | 13 | 45.0 (6.8) | 73.0  (8.2) | 27.3 (1.6) | 0 | NA | 5.03 (0.60) |  |  |  |
| Hollander2017^5^ (NCT02243202) | 26 | Canagliflozin 300mg | 84 | 45.2 (11.0) | 103.3  (19.1) | 37.3 (4.7) | 0 | 5.6 (0.4) | NA | 600-kcal/d deficit | ≥150 min/wk | hypertensive medication and/or lipid-lowering medication |
|  |  | Placebo | 82 | 44.8 (11.1) | 104.3  (18.2) | 38.0 (5.2) | 0 | 5.6 (0.3) | NA |  |  |  |

| Iacobellis2020^6^ (NCT02235298) | 24 | Dapagliflozin 10mg | 50 | 52  (9) | 104  (28) | 36.6 (7.8) | NA | 6.8 (0.9) | 7.2  (1.3) | maintain their individualized prestudy diet | maintain their individualized prestudy exercise | metformin |
| --- | --- | --- | --- | --- | --- | --- | --- | --- | --- | --- | --- | --- |
|  |  | Placebo | 50 | 51 (11) | 96.9  (23) | 34.7 (6) | NA | 6.7 (0.8) | 7.1  (1.5) |  |  |  |
| Neeland2020^7^ (NCT02833415) | 12 | Empagliflozin 10mg | 18 | 50.5 (9.6) | 101.6  (14.8) | 37.0 (4.4) | 0 | 5.6  (0.4) | 5.3  (0.4) | NA | NA | NA |
|  |  | Placebo | 17 | 54.0 (8.1) | 103.0  (14.1) | 35.0 (5.2) | 0 | 5.9  (0.4) | 5.3  (0.4) |  |  |  |
| Newman2018^8^ (NCT02371187) | 12 | Dapagliflozin 10mg | 15 | 28 (12) | 87.8  (17.3) | 31.3 (5.3) | 0 | NA | 4.4  (0.4) | NA | exercise counseling | NA |
|  |  | Placebo | 15 | 24 (10) | 91.8  (17.0) | 31.2 (4.3) | 0 | NA | 4.2  (0.3) |  |  |  |
| Rosenstock2012^9^ (NCT00642278) | 12 | Canagliflozin 300mg | 64 | 55.2 (7.1) | 86.0  (19.7) | 31.8 (5.2) | 5.8  (4.6) | 7.73 (0.89) | 8.7  (1.9) | NA | NA | metformin |
|  |  | Placebo | 65 | 53.3 (7.8) | 85.9  (19.5) | 30.6 (4.6) | 6.4  (5.0) | 7.75 (0.83) | 9.1  (2.1) |  |  |  |
| Rosenstock2014^10^ (NCT01306214) | 52 | Empagliflozin 10mg | 190 | 58.0 (9.4) | 95.9  (17.3) | 35.0 (4.0) | NA | 8.29 (0.72) | 8.34  (2.70) | diet counseling | exercise counseling | metformin and/or insulin |
|  |  | Placebo | 189 | 56.7 (10.1) | 95.5  (17.5) | 34.7 (4.3) | NA | 8.33 (0.72) | 8.41  (2.54) |  |  |  |
| Ryan2020^11^ (NCT03180489) | 12 | Dapagliflozin 10mg | 25 | 38  (11) | 96.5  (15.6) | 33.5 (3.9) | 0 | NA | 4.23  (0.43) | diet counseling | maintain their individualized prestudy exercise | NA |
|  |  | Placebo | 25 | 35  (11) | 93.8  (20.4) | 33.2 (5.6) | 0 | NA | 4.15  (0.38) |  |  |  |
| Van Ruiten2021^12^ (NCT03361098) | 16 | Dapagliflozin 10mg | 16 | 64.1 (8.4) | NA | 31.7 (3.3) | 8  (7.5) | 7.76 (0.74) | NA | NA | NA | metformin and/or SUs |
|  |  | Exenatide  10μg | 16 | 65.0 (5.8) | NA | 32.7 (5.1) | 10  (11.7) | 8.1 (1.03) | NA |  |  |  |
|  |  | Placebo | 17 | 61.5 (7.2) | NA | 31.5 (5.7) | 10.4  (7.5) | 8.07 (1.03) | NA |  |  |  |
| Apovian2010^13^ | 24 | Exenatide  10μg | 97 | 54.5 (10.0) | 94.9  (16.5) | 33.6 (3.7) | 5.7  (5.5) | 7.7  (0.9) | 8.9  (2.4) | 600-kcal/d deficit | ≥150 min/wk | metformin or SUs |
|  |  | Placebo | 99 | 55.1 (9.0) | 96.2  (15.6) | 33.9 (4.3) | 5.3  (5.1) | 7.5  (0.8) | 8.4  (2.1) |  |  |  |
| Armstrong2016^14^ (NCT01237119) | 48 | liraglutide  1.8mg | 26 | 50  (11) | 101  (18) | 34.2 (4.7) | NA | 5.9  (0.7) | 6.0  (1.7) | diet counseling | exercise counseling | metformin and/or SUs |
|  |  | Placebo | 26 | 52  (12) | 108  (18) | 37.7 (6.2) | NA | 6.0  (0.9) | 6.1  (1.5) |  |  |  |
| Astrup2009^15^ (NCT00422058) | 20 | Liraglutide  1.8mg | 90 | 45.5 (10.9) | 98.0  (12.5) | 35.0 (2.6) | 0 | 5.60 (0.40) | 5.29  (0.56) | 500-kcal/d deficit | maintain or increase their individualized prestudy exercise | hypertensive medication and/or cholesterol-lowering medication |
|  |  | liraglutide  3.0mg | 93 | 45.9 (10.7) | 97.6  (13.7) | 34.8 (2.8) | 0 | 5.57  (0.40) | 5.36  (0.61) |  |  |  |
|  |  | Placebo | 98 | 45.9 (10.3) | 97.3  (12.3) | 34.9 (2.8) | 0 | 5.60 (0.38) | 5.42   (0.81) |  |  |  |
| Basolo2018^16^ (NCT00856609) | 24 | Exenatide  10μg | 40 | 35.1 (8.4) | 105.5  (19.2) | 38.0 (6.6) | 0 | NA | 5.38  (0.47) | diet counseling | exercise counseling | NA |
|  |  | Placebo | 39 | 33.7 (9.0) | 108.8  (20.7) | 38.2 (6.8) | 0 | NA | 5.49  (0.49) |  |  |  |
| Blackman2016^17^ (NCT01557166) | 32 | Liraglutide  3.0mg | 180 | 48.6 (9.9) | 116.5  (23.0) | 38.9 (6.4) | 0 | 5.7  (0.4) | 5.4  (0.6) | 500-kcal/d deficit | ≥150 min/wk | NA |
|  |  | Placebo | 179 | 48.4 (9.5) | 118.7  (25.4) | 39.4 (7.4) | 0 | 5.6  (0.4) | 5.4  (0.9) |  |  |  |
| Buse2004^18^ | 30 | Exenatide  10μg | 129 | 56  (11) | 95  (18) | 33  (6) | 6.6  (6.6) | 8.6  (1.2) | 9.9  (2.8) | NA | NA | SUs |
|  |  | Placebo | 123 | 55  (11) | 99  (18) | 34  (5) | 5.7  (4.7) | 8.7  (1.2) | 10.8  (3.2) |  |  |  |
| Davies2015^19^ (NCT01620489) | 26 | Liraglutide  1.8mg | 140 | 68.0 (8.3) | 93.63  (17.41) | 33.4 (5.4) | 15.9  (8.9) | 8.08 (0.79) | 9.48  (3.27) | NA | NA | anti-diabetic |
|  |  | Placebo | 139 | 66.3 (8.0) | 95.63  (17.65) | 34.5 (5.4) | 14.2  (7.5) | 8.00 (0.85) | 9.27  (2.84) |  |  |  |

| Davies2016^20^ (NCT01272232) | 56 | Liraglutide  1.8mg | 211 | 54.9 (10.7) | 105.8  (21.0) | 37.0 (6.9) | 7.4  (5.16) | 8.0  (0.8) | 8.9  (2.0) | 500-kcal/d deficit | ≥150 min/wk | oral anti-diabetic |
| --- | --- | --- | --- | --- | --- | --- | --- | --- | --- | --- | --- | --- |
|  |  | Liraglutide  3.0mg | 423 | 55.0 (10.8) | 105.7  (21.9) | 37.1 (6.5) | 7.5  (5.65) | 7.9  (0.8) | 8.8  (1.9) |  |  |  |
|  |  | Placebo | 212 | 54.7 (9.8) | 106.5  (21.3) | 37.4 (7.1) | 6.7  (5.07) | 7.9  (0.8) | 8.6  (1.8) |  |  |  |
| Davies2017^21^  (NCT01923181) | 26 | Semaglutide 1mg | 70 | 56.8  (11.8) | 88.8  (15.4) | 30.7  (4.0) | 5.6  (5.0) | 7.8  (0.7) | 9.6  (2.47) | diet counseling | exercise counseling | metformin |
|  |  | Placebo | 71 | 58.9  (10.3) | 93.8  (18.1) | 32.6  (4.5) | 6.7  (5.1) | 8.0  (0.8) | 9.54  (2.69) |  |  |  |
| Davies2021^22^  (NCT03552757) | 68 | Semaglutide 2.4mg | 404 | 55  (11) | 99.9  (22.5) | 35.9 (6.4) | 8.2  (6.2) | 8.1  (0.8) | 8.5  (2.3) | 500-kcal/d deficit | ≥150 min/wk | oral anti-diabetic |
|  |  | Semaglutide 1mg | 403 | 56  (10) | 99.0  (21.1) | 35.3  (5.9) | 7.7  (5.9) | 8.1  (0.8) | 8.6  (2.3) |  |  |  |
|  |  | Placebo | 403 | 55  (11) | 100.5  (20.9) | 35.9 (6.5) | 8.2  (6.2) | 8.1  (0.8) | 8.8  (2.3) |  |  |  |
| Defronzo2005^23^ | 30 | Exenatide  10μg | 113 | 52  (11) | 101  (20) | 34  (6) | 4.9  (4.7) | 8.2  (1.0) | 9.3  (2.5) | diet counseling | NA | metformin |
|  |  | Placebo | 113 | 54  (9) | 100  (19) | 34  (6) | 6.6  (6.1) | 8.2  (1.0) | 9.4  (2.2) |  |  |  |
| Dejgaard2016^24^ (NCT01612468) | 24 | Liraglutide  1.8mg | 50 | 47  (13) | 93.4  (14.2) | 30.3 (3.5) | 20  (12) | 8.7  (0.7) | 13.0  (4.74) | NA | NA | insulin |
|  |  | Placebo | 50 | 49  (12) | 94.0  (12.5) | 29.8 (3.1) | 25  (12) | 8.7  (0.7) | 14.4  (3.82) |  |  |  |
| Derosa2012^25^ | 52 | Exenatide  10μg | 86 | 57.3 (7.7) | 89.0  (9.7) | 31.9 (1.7) | 7.6  (2.8) months | 8.1  (0.8) | 7.8  (0.9) | 600-kcal/d deficit | walk briskly or bicycle 20-30min/time,2-3times/week | metformin |
|  |  | Placebo | 85 | 56.7 (7.3) | 90.5  (10.3) | 31.7 (1.5) | 7.8  (3.1) months | 7.9  (0.6) | 7.6  (0.8) |  |  |  |

| Frias2017^26^ (NCT02205528) | 12 | Liraglutide  1.8mg | 35 | 55.2 (7.87) | 90.9  (17.98) | 32.35 (4.31) | 7.8  (5.24) | 8.36 (0.96) | 9.28  (2.02) | maintain their individualized prestudy diet | maintain their individualized prestudy exercise | metformin |
| --- | --- | --- | --- | --- | --- | --- | --- | --- | --- | --- | --- | --- |
|  |  | Placebo | 36 | 54.6 (7.91) | 89.9  (21.05) | 33.29 (5.02) | 7.6  (5.71) | 8.22 (0.87) | 9.29  (2.27) |  |  |  |
| Frias2019^27^ | 20 | Dulaglutide  1.5mg | 81 | 57.7 (9.8) | 87.9  (17.0) | 32.2 (4.8) | NA | 8.0  (0.8) | 9.5  (2.5) | NA | NA | metformin |
|  |  | Placebo | 82 | 56.5 (8.9) | 94.5  (22.9) | 33.2 (6.0) | NA | 8.1  (0.8) | 9.1  (2.2) |  |  |  |
| Friedrichsen2020^28^ (NCT03842202) | 20 | Semaglutide 2.4mg | 36 | 40.7 (12.2) | 106.2  (16.2) | 34.2 (3.0) | 0 | NA | NA | ad libitum meal | NA | NA |
|  |  | Placebo | 36 | 45.0 (9.5) | 104.9  (14.0) | 34.6 (3.1) | 0 | NA | NA |  |  |  |
| Ghanim2020^29^ (NCT01753362) | 26 | Liraglutide  1.8mg | 42 | 47  (2) | 94.2  (3.1) | 33.3 (1.2) | NA | 7.96 (0.19) | 8.56  (0.44) | NA | NA | insulin |
|  |  | Placebo | 42 | 45  (3) | 83.3  (3.4) | 29.5 (1.3) | NA | 7.79 (0.18) | 8.22  (0.39) |  |  |  |
| Gill2010^30^ (NCT00516074) | 12 | Exenatide  10μg | 28 | 57  (11) | 91.6  (15.2) | 29.5 (3.4) | 7  (4) | 7.5  (0.9) | NA | NA | NA | metformin and/or TZD or anti-hypertensive medications |
|  |  | Placebo | 26 | 54  (10) | 85.9  (12.2) | 30.1 (3.9) | 6  (4) | 7.1  (0.7) | NA |  |  |  |
| Grunberger2012^31^ | 12 | Dulaglutide  1.5mg | 29 | 57.5 (7.9) | 85.8  (18.6) | 31.0 (4.3) | 4.6  (4.1) | 7.3  (0.4) | NA | NA | NA | drug naïve or metformin |
|  |  | Placebo | 32 | 55.0 (9.3) | 90.9  (18.9) | 32.1 (5.2) | 3.9  (4.7) | 7.4  (0.6) | NA |  |  |  |
| Guo2020^32^ (ChiCTR2000035091) | 26 | Liraglutide  1.8mg | 32 | 53.1 (6.3) | 84.3  (10.8) | 29.2 (4.2) | NA | 7.5  (1.3) | 7.1  (1.2) | diet counseling | exercise counseling | metformin |
|  |  | Placebo | 32 | 52.6 (3.9) | 82.2  (12.4) | 28.6 (3.7) | NA | 7.4  (1.0) | 7.3  (1.6) |  |  |  |

| Halawi2017^33^ (NCT02647944) | 16 | Liraglutide  3.0mg | 19 | 40.95  (11.08) | 103.7  (90.0 to  112.2)^1^ | 37.2 (33.6 to 41.0)^1^ | 0 | NA | NA | NA | NA | drug naïve |
| --- | --- | --- | --- | --- | --- | --- | --- | --- | --- | --- | --- | --- |
|  |  | Placebo | 21 | 37.81  (12.14) | 99.1  (90.4 to  111.0)^1^ | 34.6 (33.4 to 38.9)^1^ | 0 | NA | NA |  |  |  |
| Joubert2020^34^ (NCT01140893) | 26 | Exenatide  10μg | 28 | 61  (7) | 95.0  (16.7) | 35.5 (4.2) | 19.3  (7.9) | 8.8  (0.8) | NA | diet counseling | exercise counseling | insulin |
|  |  | Placebo | 18 | 58  (8) | 99.3  (17.7) | 35.1 (4.4) | 18.4  (7.7) | 8.9  (0.9) | NA |  |  |  |
| Kendall2005^35^ | 30 | Exenatide  10μg | 241 | 55  (10) | 98  (21) | 34  (6) | 8.7  (6.4) | 8.5  (1.1) | 9.9  (2.4) | diet counseling | NA | metformin and SUs |
|  |  | Placebo | 247 | 56  (10) | 99  (19) | 34  (5) | 9.4  (6.2) | 8.5  (1.0) | 10.0  (2.7) |  |  |  |
| Kim2013^36^ (NCT01784965) | 14 | Liraglutide  1.8mg | 35 | 58  (7) | 88.4  (11.2) | 31.9 (2.7) | 0 | NA | 5.8  (0.4) | 500-kcal/d deficit | maintain their individualized prestudy exercise | NA |
|  |  | Placebo | 33 | 58  (8) | 88.7  (11.2) | 31.9 (3.5) | 0 | NA | 5.9  (0.5) |  |  |  |
| Lau2021^37^  (NCT03856047) | 26 | Liraglutide  3.0mg | 99 | 51.5  (9.3) | 107.8  (24.1) | 38.4  (7.4) | 0 | 5.6  (0.4) | 5.7  (0.8) | 500-kcal/d deficit | ≥150 min/wk | NA |
|  |  | Placebo | 101 | 51.4  (11.9) | 106.2  (21.6) | 37.4  (5.7) | 0 | 5.6  (0.4) | 5.6  (0.6) |  |  |  |
| Lind2015^38^  (EudraCT2012-001941-42) | 24 | Liraglutide  1.8mg | 64 | 63.7 (8.2) | 98.9  (14.0) | 33.7 (4.3) | 17.3  (7.6) | 9.0  (1.0) | 9.9  (3.1) | diet counseling | exercise counseling | insulin |
|  |  | Placebo | 60 | 63.5 (7.7) | 100.0  (14.8) | 33.5 (4.0) | 17.0  (8.1) | 9.0  (1.1) | 9.4  (2.5) |  |  |  |
| Liutkus2010^39^ | 26 | Exenatide  10μg | 111 | 55  (8) | 94.5  (17.8) | 34  (6) | 6.3  (4.2) | 8.2  (0.9) | 9.2  (2.8) | diet counseling | NA | metformin and/or TZD |
|  |  | Placebo | 54 | 54  (9) | 92.6  (18.0) | 33  (5) | 6.4  (4.6) | 8.3  (0.9) | 9.0  (2.0) |  |  |  |

| Matikainen2018^40^ (NCT02765399) | 16 | Liraglutide  1.8mg | 15 | 62  (2) | 98.6  (11.0) | 31.8 (3.4) | 7.8  (4.7) | 7.0  (1.0) | 8.3  (2.4) | 927 kcal/d | maintain their individualized prestudy exercise | metformin |
| --- | --- | --- | --- | --- | --- | --- | --- | --- | --- | --- | --- | --- |
|  |  | Placebo | 7 | 63  (2) | 92.0  (7.4) | 33.0 (4.1) | 5.9  (6.5) | 6.3  (0.3) | 6.5  (0.8) |  |  |  |
| Mensberg2016^41^ (NCT01455441) | 16 | Liraglutide  1.8mg | 17 | 56.5  (9) | 101.0  (14.5) | 32.5 (3.7) | 6.0  (5.2) | 8.2  (1.4) | 9.9  (2.7) | maintain their individualized prestudy diet | maintain their individualized prestudy exercise | metformin |
|  |  | Placebo | 16 | 55.6 (12) | 96.8  (17.4) | 32.4 (5.2) | 3.7  (3.3) | 8.0  (1.2) | 10.2  (3.1) |  |  |  |
| Moretto2008^42^ (NCT00381342) | 24 | Exenatide  10μg | 78 | 55  (10) | 86  (16) | 31  (5) | 2  (3) | 7.8  (1.0) | 8.56  (2.22) | maintain their individualized prestudy diet | maintain their individualized prestudy exercise | anti-diabetic drug-naive |
|  |  | Placebo | 78 | 53  (9) | 86  (16) | 32  (5) | 1  (2) | 7.8  (0.9) | 8.89  (2.56) |  |  |  |
| Nahra2021^43^ (NCT03235050) | 54 | Liraglutide  1.8mg | 110 | 55.5 (9.8) | 102.1  (22.7) | 35.4 (6.1) | 7.6  (6.1) | 8.1  (1.0) | 10.27 (3.10) | NA | NA | NA |
|  |  | Placebo | 112 | 57.3 (9.5) | 98.1  (19.8) | 34.2 (5.1) | 7.6  (5.0) | 8.2  (1.1) | 10.18 (2.64) |  |  |  |
| Nauck2014^44^ (NCT00734474) | 26 | Dulaglutide 1.5mg | 304 | 54  (10) | 87  (17) | 31  (5) | 7  (6) | 8.1  (1.1) | 9.75  (3.27) | NA | NA | metformin |
|  |  | Placebo | 177 | 55  (9) | 87  (17) | 31  (4) | 7  (5) | 8.1  (1.1) | 9.86  (3.15) |  |  |  |
| Neeland2021^45^ (NCT03038620) | 40 | Liraglutide  3.0mg | 73 | 49.6 (9.8) | 101.0  (17.9) | 37.2 (6.0) | 0 | NA | NA | 500-kcal/d deficit | ≥150 min/wk | treated or untreated weight-related comorbid |
|  |  | Placebo | 55 | 50.9 (8.8) | 102.3  (17.9) | 38.1 (6.1) | 0 | NA | NA |  |  |  |
| Nexøe-Larsen2018^46^ (NCT02717858) | 12 | Liraglutide  3.0mg | 26 | 47.6 (10.4) | 98.2  (17.0) | 32.5 (3.6) | 0 | NA | NA | 500-kcal/d deficit | ≥150 min/wk | NA |
|  |  | Placebo | 26 | 47.5 (9.7) | 99.8  (14.7) | 32.6 (3.3) | 0 | NA | NA |  |  |  |
| O'Neil2018^47^ (NCT02453711) | 52 | Liraglutide  3.0mg | 103 | 49  (11) | 108.7  (21.9) | 38.6 (6.6) | 0 | 5.5  (0.4) | 5.5  (0.73) | 500-kcal/d deficit | ≥150 min/wk | unsuccessful weight-loss attempt |
|  |  | Placebo | 136 | 46  (13) | 114.2  (25.4) | 40.1 (7.2) | 0 | 5.5  (0.4) | 5.5  (0.63) |  |  |  |
| Pi-Sunyer2015^48^ (NCT01272219) | 56 | Liraglutide  3.0mg | 2487 | 45.2 (12.1) | 106.2  (21.2) | 38.3 (6.4) | 0 | 5.6  (0.4) | 5.32  (0.59) | 500-kcal/d deficit | ≥150 min/wk | treated or untreated weight-related comorbidity |
|  |  | Placebo | 1244 | 45.0 (12.0) | 106.2  (21.7) | 38.3 (6.3) | 0 | 5.6  (0.4) | 5.31  (0.54) |  |  |  |
| Pratley2019^49^ (NCT02863419) | 52 | Liraglutide  1.8mg | 284 | 56  (10) | 95.5  (21.9) | 33.4 (6.7) | 7.3 (5.3) | 8.0  (0.7) | 9.30  (2.22) | NA | NA | metformin and/or SGLT-2i |
|  |  | Placebo | 142 | 57  (10) | 93.2  (20.0) | 32.9 (6.1) | 7.8 (5.5) | 7.9  (0.7) | 9.25  (2.27) |  |  |  |
| Rosenstock2010^50^ (NCT00500370) | 24 | Exenatide  10μg | 73 | 47.01  (10.9) | 109.48  (23.07) | 39.64  (7.01) | 0 | 5.61  (0.34) | 5.37  (0.60) | diet counseling | exercise counseling | anti-diabetic |
|  |  | Placebo | 79 | 45.15  (12.7) | 107.64  (23.20) | 39.39  (7.02) | 0 | 5.58  (0.36) | 5.37  (0.62) |  |  |  |
| Rubino2021^51^ (NCT03548987) | 68 | Semaglutide 2.4mg | 535 | 47  (12) | 96.5  (22.5) | 34.5 (6.9) | 0 | 5.4  (0.3) | 4.83  (0.43) | 500-kcal/d deficit | ≥150 min/wk | treated or untreated weight-related comorbidity |
|  |  | Placebo | 268 | 46  (12) | 95.4  (22.7) | 34.1 (7.1) | 0 | 5.4  (0.3) | 4.83  (0.42) |  |  |  |
| Rubino2022^52^  (NCT04074161) | 68 | Liraglutide  1.8mg | 127 | 49  (13) | 103.7  (22.5) | 37.2  (6.4) | 0 | 5.5  (0.3) | 5.3  (0.47) | 500-kcal/d deficit | ≥150 min/wk | unsuccessful dietary weight-loss attempt |
|  |  | Semaglutide 2.4mg | 126 | 48  (14) | 102.5  (25.3) | 37.0  (7.4) | 0 | 5.5  (0.3) | 5.3  (0.57) |  |  |  |
|  |  | Placebo | 85 | 51  (12) | 108.8  (23.1) | 38.8  (6.5) | 0 | 5.6  (0.4) | 5.4  (0.68) |  |  |  |
| Schiavon2021^53^ (NCT02973321) | 26 | Liraglutide  1.8mg | 67 | 56.1  (11.4) | 103.9  (36.37) | 34.23  (5.51) | NA | 8.11  (0.86) | NA | 600-kcal/d deficit | NA | metformin |
|  |  | Placebo | 33 | 56.2  (9.3) | 89.7  (11.19) | 32.07  (4.41) | NA | 8.04  (0.86) | NA |  |  |  |
| Smits2016^54^ (NCT01744236) | 12 | Liraglutide  1.8mg | 17 | 60.8 (7.42) | 103.2  (13.19) | 32.8 (4.12) | 7.9  (4.95) | 7.4 (0.82) | 8.3  (1.24) | NA | NA | metformin and/or SUs |
|  |  | Placebo | 17 | 65.8 (5.77) | 95.8  (9.90) | 30.6 (2.89) | 8.2  (4.95) | 7.5 (0.82) | 8.9  (2.06) |  |  |  |

| Umpierrez2011^55^ (NCT00630825) | 16 | Dulaglutide  2.0mg | 65 | 54  (11) | 98.6  (18.4) | 34.2 (4.1) | 8.6  (6.9) | 8.43 (1.0) | NA | NA | NA | oral anti-diabetic |
| --- | --- | --- | --- | --- | --- | --- | --- | --- | --- | --- | --- | --- |
|  |  | Placebo | 66 | 56  (12) | 94.7  (15) | 33.9 (4.3) | 7.5  (5.4) | 8.05 (0.8) | NA |  |  |  |
| VanEyk2019^56^ (NCT01761318) | 26 | Liraglutide  1.8mg | 22 | 55  (11) | 81.9  (11.0) | 30.4 (3.8) | 19  (10) | 8.1  (0.9) | NA | NA | NA | anti-diabetic |
|  |  | Placebo | 25 | 55  (9) | 77.8  (12.4) | 28.6 (4.0) | 17  (10) | 8.6  (1.1) | NA |  |  |  |
| Wadden2018^57^ (NCT02911818) | 52 | Liraglutide  3.0mg | 50 | 45.2 (12.3) | 107.8  (17.9) | 38.5 (5.4) | 0 | 5.6  (0.3) | 4.91  (0.45) | 1500-1800 kcal/d | ≥225 min/wk | NA |
|  |  | Placebo | 50 | 48.0 (11.9) | 111.7  (19.4) | 38.8 (5.0) | 0 | 5.7  (0.4) | 4.91  (0.47) |  |  |  |
| Wadden2020^58^ (NCT02963935) | 56 | Liraglutide  3.0mg | 142 | 45.4 (11.6) | 108.5  (22.1) | 39.3 (6.8) | 0 | 5.5  (0.4) | 5.4  (0.5) | 1200-1800 kcal/d | ≥250 min/wk | drug naïve |
|  |  | Placebo | 140 | 49.0 (11.2) | 106.7  (22.0) | 38.7 (7.2) | 0 | 5.5  (0.4) | 5.4  (0.6) |  |  |  |
| Wadden2021^59^ (NCT03611582) | 68 | Semaglutide 2.4mg | 407 | 46  (13) | 106.9  (22.8) | 38.1 (6.7) | 0 | 5.7  (0.3) | 5.22  (0.52) | 1200-1800 kcal/d | ≥200 min/wk | drug naïve |
|  |  | Placebo | 204 | 46  (13) | 103.7  (22.9) | 37.8 (6.9) | 0 | 5.8  (0.3) | 5.22  (0.54) |  |  |  |
| Wilding2021^60^ (NCT03548935) | 68 | Semaglutide 2.4mg | 1306 | 46  (13) | 105.4  (22.1) | 37.8 (6.7) | 0 | 5.7  (0.3) | NA | 500-kcal/d deficit | ≥150 min/wk | teated or untreated weight-related comorbidity |
|  |  | Placebo | 655 | 47  (12) | 105.2  (21.5) | 38.0 (6.5) | 0 | 5.7  (0.3) | NA |  |  |  |
| Zinman2007^61^ (NCT00099320) | 16 | Exenatide  10μg | 121 | 55.6 (10.8) | 97.5  (18.8) | 34.0 (5.1) | 7.3 (4.9) | 7.9  (0.9) | 9.1  (2.6) | NA | NA | metformin and/or TZD |
|  |  | Placebo | 112 | 56.6 (10.2) | 96.9  (19.0) | 34.0 (5.0) | 8.2 (5.8) | 7.9  (0.8) | 8.8  (1.9) |  |  |  |

### Supplement Table 3. The risk of bias of individual studies.

| Study | Bias domain | | | | | |
| --- | --- | --- | --- | --- | --- | --- |
|  | Bias arising from the  randomization process | Bias due to deviations from intended interventions | Bias due to missing  outcome data | Bias in measurement  of the outcome | Bias in selection of the reported result | Overall bias |
| Bays2014 | Low risk | Low risk | Low risk | Low risk | Low risk | Low risk |
| Bolinder2012 | Low risk | Low risk | Low risk | Low risk | Low risk | Low risk |
| Eriksson2018 | Low risk | Low risk | Low risk | Low risk | Low risk | Low risk |
| González-Ortiz2017 | Low risk | Low risk | Some concerns | Low risk | Low risk | Some concerns |
| Hollander2017 | Low risk | Low risk | Low risk | Low risk | Low risk | Low risk |
| Iacobellis2020 | Low risk | Low risk | Low risk | Low risk | Low risk | Low risk |
| Neeland2020 | Low risk | Low risk | Low risk | Low risk | Low risk | Low risk |
| Newman2018 | Low risk | Low risk | Some concerns | Low risk | Low risk | Some concerns |
| Rosenstock2012 | Low risk | Low risk | Low risk | Low risk | Low risk | Low risk |
| Rosenstock2014 | Low risk | Low risk | Low irsk | Low risk | Low risk | Low risk |
| Ryan2020 | Low risk | Low risk | Some concerns | Low risk | Low risk | Some concerns |
| Van Ruiten2021 | Low risk | Low risk | Low risk | Low risk | Low risk | Low risk |
| Apovian2010 | Low risk | Low risk | Low risk | Low risk | Low risk | Low risk |
| Armstrong2016 | Low risk | Low risk | Low risk | Low risk | Low risk | Low risk |
| Astrup2009 | Low risk | Low risk | Low risk | Low risk | Low risk | Low risk |
| Basolo2018 | Low risk | Low risk | Low risk | Low risk | Low risk | Low risk |
| Blackman2016 | Low risk | Low risk | Low risk | Low risk | Low risk | Low risk |
| Buse2004 | Low risk | Low risk | Low risk | Low risk | Low risk | Low risk |
| Davies2015 | Low risk | Low risk | Low risk | Low risk | Low risk | Low risk |
| Davies2016 | Low risk | Low risk | Low risk | Low risk | Low risk | Low risk |
| Davies2017 | Low risk | Low risk | Low risk | Low risk | Low risk | Low risk |
| Davies2021 | Low risk | Low risk | Low risk | Low risk | Low risk | Low risk |
| Defronzo2005 | Low risk | Low risk | Low risk | Low risk | Low risk | Low risk |
| Dejgaard2016 | Low risk | Low risk | Low risk | Low risk | Low risk | Low risk |
| Derosa2012 | Low risk | Low risk | Low risk | Low risk | Low risk | Low risk |
| Frias2017 | Some concerns | Low risk | Low risk | Low risk | Low risk | Some concerns |
| Frias2019 | Low risk | Low risk | Low risk | Low risk | Low risk | Low risk |
| Friedrichsen2020 | Low risk | Low risk | Low risk | Low risk | Low risk | Low risk |
| Ghanim2020 | Low risk | Low risk | Low risk | Low risk | Low risk | Low risk |
| Gill2010 | Some concerns | Low risk | Low risk | Low risk | Low risk | Some concerns |
| Grunberger2012 | Low risk | Low risk | Low risk | Low risk | Low risk | Low risk |
| Guo2020 | Low risk | Some concerns | Low risk | Low risk | Low risk | Some concerns |
| Halawi2017 | Low risk | Low risk | Low risk | Low risk | Low risk | Low risk |
| Joubert2020 | Some concerns | Some concerns | Low risk | Low risk | Low risk | Some concerns |
| Kendall2005 | Low risk | Low risk | Low risk | Low risk | Low risk | Low risk |
| Kim2013 | Low risk | Low risk | Some concerns | Low risk | Low risk | Some concerns |
| Lau2021 | Low risk | Low risk | Low risk | Low risk | Low risk | Low risk |
| Lind2015 | Low risk | Low risk | Low risk | Low risk | Low risk | Low risk |
| Liutkus2010 | Low risk | Low risk | Low risk | Low risk | Low risk | Low risk |
| Matikainen2018 | Low risk | Some concerns | Some concerns | Low risk | Low risk | High risk |
| Mensberg2016 | Low risk | Low risk | Some concerns | Low risk | Low risk | Some concerns |
| Moretto2008 | Low risk | Low risk | Low risk | Low risk | Low risk | Low risk |
| Nahra2021 | Low risk | Low risk | Low risk | Low risk | Low risk | Low risk |
| Nauck2014 | Low risk | Low risk | Low risk | Low risk | Low risk | Low risk |
| Neeland2021 | Low risk | Low risk | Low risk | Low risk | Low risk | Low risk |
| Nexøe-Larsen2018 | Low risk | Low risk | Low risk | Low risk | Low risk | Low risk |
| O'Neil2018 | Low risk | Low risk | Low risk | Low risk | Low risk | Low risk |
| Pi-Sunyer2015 | Low risk | Low risk | Low risk | Low risk | Low risk | Low risk |
| Pratley2019 | Low risk | Low risk | Low risk | Low risk | Low risk | Low risk |
| Rosenstock2010 | Low risk | Low risk | Low risk | Low risk | Low risk | Low risk |
| Rubino2021 | Low risk | Low risk | Low risk | Low risk | Low risk | Low risk |
| Rubino2022 | Low risk | Low risk | Low risk | Low risk | Low risk | Low risk |
| Schiavon2021 | Some cocerns | High risk | Low risk | Low risk | Low risk | High risk |
| Smits2016 | Low risk | Low risk | Some concerns | Low risk | Low risk | Some concerns |
| Umpierrez2011 | Low risk | Low risk | Some concerns | Low risk | Low risk | Some concerns |
| Van Eyk2019 | Low risk | Low risk | Low risk | Low risk | Low risk | Low risk |
| Wadden2018 | Some concerns | Low risk | Low risk | Low risk | Low risk | Some concerns |
| Wadden2020 | Low risk | Low risk | Low risk | Low risk | Low risk | Low risk |
| Thomas2021 | Low risk | Low risk | Low risk | Low risk | Low risk | Low risk |
| Wilding2021 | Low risk | Low risk | Low risk | Low risk | Low risk | Low risk |
| Zinman2007 | Low risk | Low risk | Low risk | Low risk | Low risk | Low risk |

### Supplement Table 4. GRADE Quality of evidence.

Supplement Table 4A. Body weight.

| **Intervention** | | **Body weight** |
| --- | --- | --- |
| **Compared with Placbo** | | |
| Canagliflozin 300mg | | Low |
| Dapagliflozin 10mg | | Moderate |
| Empagliflozin 10mg | | Low |
| Exenatide 10ug | | Moderate |
| Dulaglutide 1.5mg | | Low |
| Liraglutide 1.8mg | | Moderate |
| Liraglutide 3.0mg | | Moderate |
| Semaglutide 1.0mg | | Low |
| Semaglutide 2.4mg | | Moderate |
| **Compared with Canagliflozin 300mg** | | |
| Dapagliflozin 10mg | | Low |
| Empagliflozin 10mg | | Low |
| Exenatide 10ug | | Low |
| Dulaglutide 1.5mg | | Low |
| Liraglutide 1.8mg | | Low |
| Liraglutide 3.0mg | | Moderate |
| Semaglutide 1.0mg | | Low |
| Semaglutide 2.4mg | | Moderate |
| **Compared with Dapagliflozin 10mg** | | |
| Empagliflozin 10mg | | Low |
| Exenatide 10ug | | Low |
| Dulaglutide 1.5mg | | Low |
| Liraglutide 1.8mg | | Low |
| Liraglutide 3.0mg | | Low |
| Semaglutide 1.0mg | | Low |
| Semaglutide 2.4mg | | Moderate |
| **Compared with Empagliflozin 10mg** | | |
| Exenatide 10ug | | Low |
| Dulaglutide 1.5mg | | Low |
| Liraglutide 1.8mg | | Low |
| Liraglutide 3.0mg | | Low |
| Semaglutide 1.0mg | | Low |
| Semaglutide 2.4mg | | Moderate |
| **Compared with Exenatide 10ug** | | |
| Dulaglutide 1.5mg | | Low |
| Liraglutide 1.8mg | | Low |
| Liraglutide 3.0mg | | Moderate |
| Semaglutide 1.0mg | | Moderate |
| Semaglutide 2.4mg | | Moderate |
| **Compared with Dulaglutide 1.5mg** | | |
| Liraglutide 1.8mg | Low | |
| Liraglutide 3.0mg | Low | |
| Semaglutide 1.0mg | Moderate | |
| Semaglutide 2.4mg | Moderate | |
| **Compared with Liraglutide 1.8mg** | | |
| Liraglutide 3.0mg | High | |
| Semaglutide 1.0mg | Moderate | |
| Semaglutide 2.4mg | Moderate | |
| **Compared with Liraglutide 3.0mg** | | |
| Semaglutide 1.0mg | Low | |
| Semaglutide 2.4mg | Moderate | |
| **Compared with Semaglutide 1.0mg** | | |
| Semaglutide 2.4mg | Moderate | |

Supplement Table 4B. Achieving at least 5% weight loss.

| **Intervention** | **Achieving at least 5% weight loss** |
| --- | --- |
| **Compared with Placbo** | |
| Canagliflozin 300mg | Low |
| Exenatide 10ug | Moderate |
| Liraglutide 1.8mg | Low |
| Liraglutide 3.0mg | Moderate |
| Semaglutide 1.0mg | Low |
| Semaglutide 2.4mg | Moderate |
| **Compared with Canagliflozin 300mg** | |
| Exenatide 10ug | Moderate |
| Liraglutide 1.8mg | Low |
| Liraglutide 3.0mg | Moderate |
| Semaglutide 1.0mg | Moderate |
| Semaglutide 2.4mg | Low |
| **Compared with Exenatide 10ug** | |
| Liraglutide 1.8mg | Low |
| Liraglutide 3.0mg | Moderate |
| Semaglutide 1.0mg | Low |
| Semaglutide 2.4mg | Moderate |
| **Compared with Liraglutide 1.8mg** | |
| Liraglutide 3.0mg | Moderate |
| Semaglutide 1.0mg | Low |
| Semaglutide 2.4mg | Moderate |
| **Compared with Liraglutide 3.0mg** | |
| Semaglutide 1.0mg | Low |
| Semaglutide 2.4mg | Moderate |
| **Compared with Semaglutide 1.0mg** | |
| Semaglutide 2.4mg | Moderate |

Supplement Table 4C. HbA1c.

| **Intervention** | | **HbA1c** |
| --- | --- | --- |
| **Compared with Placbo** | | |
| Canagliflozin 300mg | | Low |
| Dapagliflozin 10mg | | Moderate |
| Empagliflozin 10mg | | Low |
| Exenatide 10ug | | Low |
| Dulaglutide 1.5mg | | Moderate |
| Liraglutide 1.8mg | | Moderate |
| Liraglutide 3.0mg | | Moderate |
| Semaglutide 1.0mg | | Low |
| Semaglutide 2.4mg | | Moderate |
| **Compared with Canagliflozin 300mg** | | |
| Dapagliflozin 10mg | | Low |
| Empagliflozin 10mg | | Low |
| Exenatide 10ug | | Low |
| Dulaglutide 1.5mg | | Low |
| Liraglutide 1.8mg | | Low |
| Liraglutide 3.0mg | | Low |
| Semaglutide 1.0mg | | Low |
| Semaglutide 2.4mg | | Moderate |
| **Compared with Dapagliflozin 10mg** | | |
| Empagliflozin 10mg | | Low |
| Exenatide 10ug | | Moderate |
| Dulaglutide 1.5mg | | Low |
| Liraglutide 1.8mg | | Low |
| Liraglutide 3.0mg | | Low |
| Semaglutide 1.0mg | | Low |
| Semaglutide 2.4mg | | Moderate |
| **Compared with Empagliflozin 10mg** | | |
| Exenatide 10ug | | Low |
| Dulaglutide 1.5mg | | Low |
| Liraglutide 1.8mg | | Low |
| Liraglutide 3.0mg | | Low |
| Semaglutide 1.0mg | | Low |
| Semaglutide 2.4mg | | Low |
| **Compared with Exenatide 10ug** | | |
| Dulaglutide 1.5mg | | Low |
| Liraglutide 1.8mg | | Low |
| Liraglutide 3.0mg | | Moderate |
| Semaglutide 1.0mg | | Moderate |
| Semaglutide 2.4mg | | Low |
| **Compared with Dulaglutide 1.5mg** | | |
| Liraglutide 1.8mg | Moderate | |
| Liraglutide 3.0mg | Moderate | |
| Semaglutide 1.0mg | Low | |
| Semaglutide 2.4mg | Low | |
| **Compared with Liraglutide 1.8mg** | | |
| Liraglutide 3.0mg | Moderate | |
| Semaglutide 1.0mg | Low | |
| Semaglutide 2.4mg | Low | |
| **Compared with Liraglutide 3.0mg** | | |
| Semaglutide 1.0mg | Low | |
| Semaglutide 2.4mg | Low | |
| **Compared with Semaglutide 1.0mg** | | |
| Semaglutide 2.4mg | Low | |

Supplement Table 4D. Fasting plasma glucose (FPG).

| **Intervention** | | **FPG** |
| --- | --- | --- |
| **Compared with Placbo** | | |
| Canagliflozin 300mg | | Low |
| Dapagliflozin 10mg | | Moderate |
| Empagliflozin 10mg | | Low |
| Exenatide 10ug | | Low |
| Dulaglutide 1.5mg | | Moderate |
| Liraglutide 1.8mg | | Moderate |
| Liraglutide 3.0mg | | Moderate |
| Semaglutide 1.0mg | | Low |
| Semaglutide 2.4mg | | Moderate |
| **Compared with Canagliflozin 300mg** | | |
| Dapagliflozin 10mg | | Low |
| Empagliflozin 10mg | | Low |
| Exenatide 10ug | | Low |
| Dulaglutide 1.5mg | | Low |
| Liraglutide 1.8mg | | Low |
| Liraglutide 3.0mg | | Low |
| Semaglutide 1.0mg | | Low |
| Semaglutide 2.4mg | | Moderate |
| **Compared with Dapagliflozin 10mg** | | |
| Empagliflozin 10mg | | Low |
| Exenatide 10ug | | Moderate |
| Dulaglutide 1.5mg | | Low |
| Liraglutide 1.8mg | | Low |
| Liraglutide 3.0mg | | Low |
| Semaglutide 1.0mg | | Low |
| Semaglutide 2.4mg | | Moderate |
| **Compared with Empagliflozin 10mg** | | |
| Exenatide 10ug | | Low |
| Dulaglutide 1.5mg | | Low |
| Liraglutide 1.8mg | | Low |
| Liraglutide 3.0mg | | Low |
| Semaglutide 1.0mg | | Low |
| Semaglutide 2.4mg | | Low |
| **Compared with Exenatide 10ug** | | |
| Dulaglutide 1.5mg | | Low |
| Liraglutide 1.8mg | | Low |
| Liraglutide 3.0mg | | Moderate |
| Semaglutide 1.0mg | | Moderate |
| Semaglutide 2.4mg | | Low |
| **Compared with Dulaglutide 1.5mg** | | |
| Liraglutide 1.8mg | Moderate | |
| Liraglutide 3.0mg | Moderate | |
| Semaglutide 1.0mg | Low | |
| Semaglutide 2.4mg | Low | |
| **Compared with Liraglutide 1.8mg** | | |
| Liraglutide 3.0mg | Moderate | |
| Semaglutide 1.0mg | Low | |
| Semaglutide 2.4mg | Low | |
| **Compared with Liraglutide 3.0mg** | | |
| Semaglutide 1.0mg | Low | |
| Semaglutide 2.4mg | Low | |
| **Compared with Semaglutide 1.0mg** | | |
| Semaglutide 2.4mg | Low | |

Supplement Table 4E. Systolic blood pressure (SBP).

| **Intervention** | | **SBP** |
| --- | --- | --- |
| **Compared with Placbo** | | |
| Canagliflozin 300mg | | Low |
| Dapagliflozin 10mg | | Moderate |
| Empagliflozin 10mg | | Low |
| Exenatide 10ug | | Moderate |
| Dulaglutide 1.5mg | | Low |
| Liraglutide 1.8mg | | Moderate |
| Liraglutide 3.0mg | | Moderate |
| Semaglutide 1.0mg | | Low |
| Semaglutide 2.4mg | | Moderate |
| **Compared with Canagliflozin 300mg** | | |
| Dapagliflozin 10mg | | Low |
| Empagliflozin 10mg | | Low |
| Exenatide 10ug | | Low |
| Dulaglutide 1.5mg | | Low |
| Liraglutide 1.8mg | | Low |
| Liraglutide 3.0mg | | Low |
| Semaglutide 1.0mg | | Low |
| Semaglutide 2.4mg | | Low |
| **Compared with Dapagliflozin 10mg** | | |
| Empagliflozin 10mg | | Low |
| Exenatide 10ug | | Low |
| Dulaglutide 1.5mg | | Low |
| Liraglutide 1.8mg | | Low |
| Liraglutide 3.0mg | | Low |
| Semaglutide 1.0mg | | Low |
| Semaglutide 2.4mg | | Low |
| **Compared with Empagliflozin 10mg** | | |
| Exenatide 10ug | | Low |
| Dulaglutide 1.5mg | | Low |
| Liraglutide 1.8mg | | Low |
| Liraglutide 3.0mg | | Low |
| Semaglutide 1.0mg | | Low |
| Semaglutide 2.4mg | | Low |
| **Compared with Exenatide 10ug** | | |
| Dulaglutide 1.5mg | | Low |
| Liraglutide 1.8mg | | Moderate |
| Liraglutide 3.0mg | | Moderate |
| Semaglutide 1.0mg | | Moderate |
| Semaglutide 2.4mg | | Moderate |
| **Compared with Dulaglutide 1.5mg** | | |
| Liraglutide 1.8mg | Moderate | |
| Liraglutide 3.0mg | Moderate | |
| Semaglutide 1.0mg | Low | |
| Semaglutide 2.4mg | Moderate | |
| **Compared with Liraglutide 1.8mg** | | |
| Liraglutide 3.0mg | Moderate | |
| Semaglutide 1.0mg | Low | |
| Semaglutide 2.4mg | Moderate | |
| **Compared with Liraglutide 3.0mg** | | |
| Semaglutide 1.0mg | Low | |
| Semaglutide 2.4mg | Moderate | |
| **Compared with Semaglutide 1.0mg** | | |
| Semaglutide 2.4mg | Moderate | |

Supplement Table 4F. Diastolic blood pressure (DBP).

| **Intervention** | | **DBP** |
| --- | --- | --- |
| **Compared with Placbo** | | |
| Canagliflozin 300mg | | Low |
| Dapagliflozin 10mg | | Moderate |
| Exenatide 10ug | | Low |
| Dulaglutide 1.5mg | | Moderate |
| Liraglutide 1.8mg | | Low |
| Liraglutide 3.0mg | | Moderate |
| Semaglutide 1.0mg | | Moderate |
| Semaglutide 2.4mg | | Low |
| **Compared with Canagliflozin 300mg** | | |
| Dapagliflozin 10mg | | Low |
| Exenatide 10ug | | Low |
| Dulaglutide 1.5mg | | Low |
| Liraglutide 1.8mg | | Low |
| Liraglutide 3.0mg | | Low |
| Semaglutide 1.0mg | | Low |
| Semaglutide 2.4mg | | Low |
| **Compared with Dapagliflozin 10mg** | | |
| Exenatide 10ug | | Low |
| Dulaglutide 1.5mg | | Low |
| Liraglutide 1.8mg | | Low |
| Liraglutide 3.0mg | | Low |
| Semaglutide 1.0mg | | Low |
| Semaglutide 2.4mg | | Low |
| **Compared with Exenatide 10ug** | | |
| Dulaglutide 1.5mg | | Low |
| Liraglutide 1.8mg | | Moderate |
| Liraglutide 3.0mg | | Moderate |
| Semaglutide 1.0mg | | Moderate |
| Semaglutide 2.4mg | | Moderate |
| **Compared with Dulaglutide 1.5mg** | | |
| Liraglutide 1.8mg | Moderate | |
| Liraglutide 3.0mg | Moderate | |
| Semaglutide 1.0mg | Low | |
| Semaglutide 2.4mg | Moderate | |
| **Compared with Liraglutide 1.8mg** | | |
| Liraglutide 3.0mg | Moderate | |
| Semaglutide 1.0mg | Low | |
| Semaglutide 2.4mg | Moderate | |
| **Compared with Liraglutide 3.0mg** | | |
| Semaglutide 1.0mg | Low | |
| Semaglutide 2.4mg | Moderate | |
| **Compared with Semaglutide 1.0mg** | | |
| Semaglutide 2.4mg | Moderate | |

Supplement Table 4G. Serious adverse events (SAE).

| **Intervention** | | **SAE** |
| --- | --- | --- |
| **Compared with Placbo** | | |
| Canagliflozin 300mg | | Low |
| Dapagliflozin 10mg | | Moderate |
| Empagliflozin 10mg | | Low |
| Exenatide 10ug | | Moderate |
| Dulaglutide 1.5mg | | Low |
| Liraglutide 1.8mg | | Moderate |
| Liraglutide 3.0mg | | Moderate |
| Semaglutide 1.0mg | | Low |
| Semaglutide 2.4mg | | Moderate |
| **Compared with Canagliflozin 300mg** | | |
| Dapagliflozin 10mg | | Low |
| Empagliflozin 10mg | | Low |
| Exenatide 10ug | | Low |
| Dulaglutide 1.5mg | | Low |
| Liraglutide 1.8mg | | Low |
| Liraglutide 3.0mg | | Low |
| Semaglutide 1.0mg | | Low |
| Semaglutide 2.4mg | | Low |
| **Compared with Dapagliflozin 10mg** | | |
| Empagliflozin 10mg | | Low |
| Exenatide 10ug | | Low |
| Dulaglutide 1.5mg | | Low |
| Liraglutide 1.8mg | | Low |
| Liraglutide 3.0mg | | Low |
| Semaglutide 1.0mg | | Low |
| Semaglutide 2.4mg | | Low |
| **Compared with Empagliflozin 10mg** | | |
| Exenatide 10ug | | Low |
| Dulaglutide 1.5mg | | Low |
| Liraglutide 1.8mg | | Low |
| Liraglutide 3.0mg | | Low |
| Semaglutide 1.0mg | | Low |
| Semaglutide 2.4mg | | Low |
| **Compared with Exenatide 10ug** | | |
| Dulaglutide 1.5mg | | Low |
| Liraglutide 1.8mg | | Moderate |
| Liraglutide 3.0mg | | Moderate |
| Semaglutide 1.0mg | | Moderate |
| Semaglutide 2.4mg | | Moderate |
| **Compared with Dulaglutide 1.5mg** | | |
| Liraglutide 1.8mg | Moderate | |
| Liraglutide 3.0mg | Moderate | |
| Semaglutide 1.0mg | Low | |
| Semaglutide 2.4mg | Moderate | |
| **Compared with Liraglutide 1.8mg** | | |
| Liraglutide 3.0mg | Moderate | |
| Semaglutide 1.0mg | Moderate | |
| Semaglutide 2.4mg | Moderate | |
| **Compared with Liraglutide 3.0mg** | | |
| Semaglutide 1.0mg | Moderate | |
| Semaglutide 2.4mg | Moderate | |
| **Compared with Semaglutide 1.0mg** | | |
| Semaglutide 2.4mg | Moderate | |

Supplement Table 4H. Discontinue due to adverse events.

| **Intervention** | **Discontinue due to adverse events** |
| --- | --- |
| **Compared with Placbo** | |
| Canagliflozin 300mg | Low |
| Dapagliflozin 10mg | Moderate |
| Empagliflozin 10mg | Low |
| Exenatide 10ug | Moderate |
| Dulaglutide 1.5mg | Low |
| Liraglutide 1.8mg | Moderate |
| Liraglutide 3.0mg | Moderate |
| Semaglutide 1.0mg | Low |
| Semaglutide 2.4mg | Moderate |
| **Compared with Canagliflozin 300mg** | |
| Dapagliflozin 10mg | Low |
| Empagliflozin 10mg | Low |
| Exenatide 10ug | Low |
| Dulaglutide 1.5mg | Low |
| Liraglutide 1.8mg | Low |
| Liraglutide 3.0mg | Low |
| Semaglutide 1.0mg | Low |
| Semaglutide 2.4mg | Low |
| **Compared with Dapagliflozin 10mg** | |
| Empagliflozin 10mg | Low |
| Exenatide 10ug | Low |
| Dulaglutide 1.5mg | Low |
| Liraglutide 1.8mg | Low |
| Liraglutide 3.0mg | Low |
| Semaglutide 1.0mg | Low |
| Semaglutide 2.4mg | Low |
| **Compared with Empagliflozin 10mg** | |
| Exenatide 10ug | Low |
| Dulaglutide 1.5mg | Low |
| Liraglutide 1.8mg | Low |
| Liraglutide 3.0mg | Low |
| Semaglutide 1.0mg | Low |
| Semaglutide 2.4mg | Low |
| **Compared with Exenatide 10ug** | |
| Dulaglutide 1.5mg | Low |
| Liraglutide 1.8mg | Moderate |
| Liraglutide 3.0mg | Moderate |
| Semaglutide 1.0mg | Moderate |
| Semaglutide 2.4mg | Moderate |
| **Compared with Dulaglutide 1.5mg** | |
| Liraglutide 1.8mg | Moderate |
| Liraglutide 3.0mg | Moderate |
| Semaglutide 1.0mg | Low |
| Semaglutide 2.4mg | Moderate |
| **Compared with Liraglutide 1.8mg** | |
| Liraglutide 3.0mg | Moderate |
| Semaglutide 1.0mg | Moderate |
| Semaglutide 2.4mg | Moderate |
| **Compared with Liraglutide 3.0mg** | |
| Semaglutide 1.0mg | Moderate |
| Semaglutide 2.4mg | Moderate |
| **Compared with Semaglutide 1.0mg** | |
| Semaglutide 2.4mg | Moderate |

### Supplement Table 5. Classification of 9 interventions, based on network meta-analysis of interventions

**Supplement Table 5A. Body weight.**

Not convincingly different than placebo (category 0)

Canagliflozin 300mg

Empagliflozin 10mg

Exenatide 10ug

Dulaglutide 1.5mg

Dapagliflozin 10mg

Intermediate—more effective than placebo (category 1)

Liraglutide 1.8mg

Liraglutide 3.0mg

More effective than at least one category 1 intervention (category 2)

Semaglutide 1.0mg

Semaglutide 2.4mg

| Certainty of the evidence, and  classification of intervention | Intervention | Intervention *v* placebo  (mean difference (95% credible interval)) | Surface under the  cumulative ranking curve |
| --- | --- | --- | --- |
| **High certainty (moderate to high certainty evidence)** | | | |
| Category 2:  among the most effective | Semaglutide 2.4mg (M) | -11.51 (-12.83, -10.21) | 0.99 |
| Category 1:  inferior to the most effective,  or superior to the least effective | Liraglutide 3.0mg (M) | -4.65 (-5.60, -3.69) | 0.79 |
|  | Liraglutide 1.8mg (M) | -3.24 (-4.43, -2.04) | 0.61 |
| Category 0:  among the least effective | Dapagliflozin 10mg (M) | -2.23 (-3.92, -0.54) | 0.43 |
|  | Exenatide 10ug  (M) | -1.27 (-2.28, -0.31) | 0.24 |
| **Low certainty (low to very low certainty evidence)** | | | |
| Category 1:  might be inferior to the most  effective or superior than the least effective | Semaglutide 1.0mg (L) | -5.67 (-7.84, -3.52) | 0.86 |
| Category 0:  might be among  the least effective | Empagliflozin 10mg (L) | -2.48 (-5.52, 0.49) | 0.43 |
|  | Canagliflozin 300mg (L) | -1.59 (-3.55, 0.37) | 0.31 |
|  | Dulaglutide 1.5mg (L) | -1.39 (-2.99, -0.2) | 0.27 |

**Supplement Table 5B. Reaching ≥ 5% weight loss**

Not convincingly different than placebo (category 0)

Canagliflozin 300mg

Exenatide 10ug

Intermediate—more effective than placebo (category 1)

Liraglutide 1.8mg

Liraglutide 3.0mg

More effective than at least one category 1 intervention (category 2)

Semaglutide 1.0mg

Semaglutide 2.4mg

| Certainty of the evidence, and  classification of intervention | Intervention | Intervention *v* placebo  (odds ratio (95% credible interval)) | Surface under the  cumulative ranking curve |
| --- | --- | --- | --- |
| **High certainty (moderate to high certainty evidence)** | | | |
| Category 2:  among the most effective | Semaglutide 2.4mg (M) | 10.88 (6.69, 18.39) | 0.97 |
| Category 1:  inferior to the most effective,  or superior to the least effective | Liraglutide 3.0mg (M) | 4.9 (3.37, 7.13) | 0.64 |
| Category 0:  among the least effective | Exenatide 10ug  (M) | 2.11 (0.88, 5.22) | 0.25 |
| **Low certainty (low to very low certainty evidence)** | | | |
| Category 2:  might be among the most effective | Semaglutide 1.0mg (L) | 7.02 (3.25, 16.50) | 0.79 |
| Category 1:  might be inferior to the most  effective or superior than the least effective | Liraglutide 1.8mg (L) | 3.81 (2.31, 6.7) | 0.49 |
| Category 0:  might be among  the least effective | Canagliflozin 300mg  (L) | 2.7 (1.19, 6.4) | 0.34 |

**Supplement Table 5C. HbA1c**

Not convincingly different than placebo (category 0)

Canagliflozin 300mg

Empagliflozin 10mg

Dapagliflozin 10mg

Intermediate—more effective than placebo (category 1)

Exenatide 10ug

Liraglutide 1.8mg

Liraglutide 3.0mg

More effective than at least one category 1 intervention (category 2)

Dulaglutide 1.5mg

Semaglutide 1.0mg

Semaglutide 2.4mg

| Certainty of the evidence, and  classification of intervention | Intervention | Intervention *v* placebo  (mean difference (95% credible interval)) | Surface under the  cumulative ranking curve |
| --- | --- | --- | --- |
| **High certainty (moderate to high certainty evidence)** | | | |
| Category 2:  among the most effective | Semaglutide 2.4mg (M) | -1.49 (-2.07, -0.92) | 0.93 |
|  | Dulaglutide 1.5mg (M) | -1.11 (-1.43, -0.77) | 0.75 |
| Category 1:  inferior to the most effective,  or superior to the least effective | Liraglutide 3.0mg  (M) | -0.98 (-1.51, -0.44) | 0.64 |
|  | Liraglutide 1.8mg  (M) | -0.75 (-0.93, -0.56) | 0.46 |
| Category 0:  among the least effective | Dapagliflozin 10mg (M) | -0.48 (-0.85, -0.10) | 0.25 |
| **Low certainty (low to very low certainty evidence)** | | | |
| Category 2:  might be among the most effective | Semaglutide 1.0mg (L) | -1.39 (-1.83, -0.95) | 0.89 |
| Category 1:  might be inferior to the most  effective or superior than the least effective | Exenatide 10ug  (L) | -0.81 (-1.02, -0.59) | 0.53 |
| Category 0:  might be among  the least effective | Empagliflozin 10mg (L) | -0.46 (-1.09, 0.16) | 0.28 |
|  | Canagliflozin 300mg (L) | -0.50 (-0.94, -0.05) | 0.27 |

**Supplement Table 5D. FPG**

Not convincingly different than placebo (category 0)

Empagliflozin 10mg

Intermediate—more effective than placebo (category 1)

Canagliflozin 300mg

Dapagliflozin 10mg

Liraglutide 1.8mg

Exenatide 10ug

Liraglutide 3.0mg

Dulaglutide 1.5mg

More effective than at least one category 1 intervention (category 2)

Semaglutide 1.0mg

Semaglutide 2.4mg

| Certainty of the evidence, and  classification of intervention | Intervention | Intervention *v* placebo  (mean difference (95% credible interval)) | Surface under the  cumulative ranking curve |
| --- | --- | --- | --- |
| **High certainty (moderate to high certainty evidence)** | | | |
| Category 2:  among the most effective | Semaglutide 2.4mg (M) | -2.15 (-2.83, -1.59) | 0.92 |
| Category 1:  inferior to the most effective,  or superior to the least effective | Liraglutide 3.0mg  (M) | -1.86 (-2.45, -1.26) | 0.75 |
|  | Dulaglutide 1.5mg (M) | -1.79 (-2.21, -1.33) | 0.71 |
|  | Liraglutide 1.8mg  (M) | -1.39 (-1.65, -1.12) | 0.45 |
|  | Dapagliflozin 10mg (M) | -1.10 (-1.68, -0.53) | 0.28 |
| **Low certainty (low to very low certainty evidence)** | | | |
| Category 2:  might be among the most effective | Semaglutide 1.0mg (L) | -2.01 (-2.67, -1.54) | 0.84 |
| Category 1:  might be inferior to the most  effective or superior than the least effective | Canagliflozin 300mg (L) | -1.50 (-2.29, -0.70) | 0.53 |
|  | Exenatide 10ug  (L) | -1.23 (-1.53, -0.94) | 0.34 |
| Category 0:  might be among  the least effective | Empagliflozin 10mg (L) | -0.81 (-1.58, -0.03) | 0.18 |

**Supplement Table 5E. SBP**

Not convincingly different than placebo (category 0)

Empagliflozin 10mg

Dulaglutide 1.5mg

Canagliflozin 300mg

Dapagliflozin 10mg

Intermediate—more effective than placebo (category 1)

Liraglutide 1.8mg

Exenatide 10ug

Liraglutide 3.0mg

Semaglutide 1.0mg

Semaglutide 2.4mg

| Certainty of the evidence, and  classification of intervention | Intervention | Intervention *v* placebo  (mean difference (95% credible interval)) | Surface under the  cumulative ranking curve |
| --- | --- | --- | --- |
| **High certainty (moderate to high certainty evidence)** | | | |
| Category 1:  inferior to the most effective,  or superior to the least effective | Semaglutide 2.4mg (M) | -4.89 (-6.04, -3.71) | 0.96 |
|  | Exenatide 10ug  (M) | -3.86 (-6.29, -1.47) | 0.78 |
|  | Liraglutide 1.8mg (M) | -3.08 (-4.30, -1.88) | 0.66 |
|  | Liraglutide 3.0mg (M) | -2.68 (-3.64, -1.69) | 0.56 |
| Category 0:  among the least effective | Dapagliflozin 10mg (M) | -1.19 (-4.51, 4.45) | 0.32 |
| **Low certainty (low to very low certainty evidence)** | | | |
| Category 1:  might be inferior to the most  effective or superior than the least effective | Semaglutide 1.0mg (L) | -3.25 (-5.46, -1.04) | 0.68 |
| Category 0:  might be among  the least effective | Dulaglutide 1.5mg (L) | -1.57 (-3.84, 0.82) | 0.36 |
|  | Canagliflozin 300mg (L) | -1.14 (-4.03, 1.80) | 0.29 |
|  | Empagliflozin 10mg (L) | -0.99 (-4.68, 4.89) | 0.28 |

**Supplement Table 5F. DBP**

Not convincingly different than placebo (category 0)

Dulaglutide 1.5mg

Canagliflozin 300mg

Dapagliflozin 10mg

Liraglutide 1.8mg

Exenatide 10ug

Liraglutide 3.0mg

Semaglutide 1.0mg

Intermediate—more effective than placebo (category 1)

Semaglutide 2.4mg

| Certainty of the evidence, and  classification of intervention | Intervention | Intervention *v* placebo  (mean difference (95% credible interval)) | Surface under the  cumulative ranking curve |
| --- | --- | --- | --- |
| **High certainty (moderate to high certainty evidence)** | | | |
| Category 1:  inferior to the most effective,  or superior to the least effective | Semaglutide 2.4mg (M) | -1.61 (-2.39, -0.85) | 0.93 |
| Category 0:  among the least effective | Exenatide 10ug  (M) | -1.27 (-2.73, 0.25) | 0.81 |
|  | Liraglutide 3.0mg (M) | -0.51 (-1.14, 0.18) | 0.58 |
|  | Dapagliflozin 10mg (M) | -0.28 (-1.74, 1.36) | 0.46 |
|  | Liraglutide 1.8mg (M) | -0.25 (-1.08, 0.62) | 0.44 |
| **Low certainty (low to very low certainty evidence)** | | | |
| Category 0:  might be among  the least effective | Dulaglutide 1.5mg (L) | -1.57 (-3.84, 0.82) | 0.46 |
|  | Semaglutide 1.0mg (L) | 0.14 (-1.17, 1.58) | 0.27 |
|  | Canagliflozin 300mg (L) | -1.14 (-4.03, 1.80) | 0.25 |

**Supplement Table 5G. Serious adverse events.**

Not convincingly different than placebo (category 0)

Empagliflozin 10mg

Dulaglutide 1.5mg

Canagliflozin 300mg

Dapagliflozin 10mg

Liraglutide 1.8mg

Exenatide 10ug

Liraglutide 3.0mg

Semaglutide 1.0mg

Intermediate—more effective than placebo (category 1)

Semaglutide 2.4mg

| Certainty of the evidence, and  classification of intervention | Intervention | Intervention *v* placebo  (mean difference (95% credible interval)) | Surface under the  cumulative ranking curve |
| --- | --- | --- | --- |
| **High certainty (moderate to high certainty evidence)** | | | |
| Category 1:  inferior to the most effective,  or superior to the least effective | Semaglutide 2.4mg (M) | 1.42 (1.01, 1.97) | 0.31 |
| Category 0:  among the least effective | Dapagliflozin 10mg (M) | 4.07 (0.89, 43.04) | 0.09 |
|  | Liraglutide 3.0mg  (M) | 1.25 (0.89, 1.72) | 0.43 |
|  | Liraglutide 1.8mg  (M) | 1.31 (0.78, 1.69) | 0.53 |
|  | Exenatide 10ug  (M) | 0.80 (0.44, 2.25) | 0.80 |
| **Low certainty (low to very low certainty evidence)** | | | |
| Category 0:  might be among  the least effective | Canagliflozin 300mg (L) | 2.47 (0.22, 76.81) | 0.30 |
|  | Dulaglutide 1.5mg (L) | 1.29 (0.58, 3.05) | 0.44 |
|  | Empagliflozin 10mg (L) | 1.01 (0.44, 2.25) | 0.64 |
|  | Semaglutide 1.0mg (L) | 0.86 (0.45, 1.54) | 0.77 |

**Supplement Table 5H. Discontinue due to adverse events**

Not convincingly different than placebo (category 0)

Empagliflozin 10mg

Dulaglutide 1.5mg

Canagliflozin 300mg

Dapagliflozin 10mg

Semaglutide 1.0mg

Intermediate—more effective than placebo (category 1)

Exenatide 10ug

Liraglutide 1.8mg

Liraglutide 3.0mg

Semaglutide 2.4mg

| Certainty of the evidence, and  classification of intervention | Intervention | Intervention *v* placebo  (mean difference (95% credible interval)) | Surface under the  cumulative ranking curve |
| --- | --- | --- | --- |
| **High certainty (moderate to high certainty evidence)** | | | |
| Category 1:  inferior to the most effective,  or superior to the least effective | Exenatide 10ug  (M) | 2.96 (1.79, 4.99) | 0.19 |
|  | Liraglutide 3.0mg  (M) | 2.88 (1.90, 4.24) | 0.19 |
|  | Liraglutide 1.8mg  (M) | 2.48 (1.49, 3.92) | 0.32 |
|  | Semaglutide 2.4mg (M) | 1.88 (1.12, 2.99) | 0.50 |
| Category 0:  among the least effective | Dapagliflozin 10mg (M) | 1.81 (0.58, 6.19) | 0.48 |
| **Low certainty (low to very low certainty evidence)** | | | |
| Category 0:  might be among  the least effective | Canagliflozin 300mg (L) | 2.30 (0.82, 7.41) | 0.36 |
|  | Semaglutide 1.0mg (L) | 2.00 (0.96, 4.87) | 0.44 |
|  | Empagliflozin 300mg (L) | 0.92 (0.32, 2.52) | 0.82 |
|  | Dulaglutide 1.5mg  (L) | 0.82 (0.39, 2.20) | 0.87 |

### Supplement Table 6. Network meta-analysis of GLP-1RAs and SGLT-2is.

Supplement Table 6A. Body wieght (kg).

| Placebo | -2.27 (-4.46, -0.08) | -4.23 (-5.27, -3.17) |
| --- | --- | --- |
| 2.27 (0.08, 4.46) | SGLT-2is | -1.96 (-4.34, 0.42) |
| 4.23 (3.17, 5.27) | 1.96 (-0.42, 4.34) | GLP-1RAs |

Supplement Table 6B. Achieving at least 5% weight loss (%).

| Placebo | 3.78 (1.56, 9.6) | 6.32 (4.46, 9.15) |
| --- | --- | --- |
| 0.26 (0.1, 0.64) | SGLT-2is | 1.67 (0.62, 4.31) |
| 0.16 (0.11, 0.22) | 0.6 (0.23, 1.61) | GLP-1RAs |

Supplement Table 6C. HbA1c (%).

| Placebo | -0.49 (-0.78, -0.2) | -0.88 (-1.01, -0.74) |
| --- | --- | --- |
| 0.49 (0.2, 0.78) | SGLT-2is | -0.39 (-0.7, -0.08) |
| 0.88 (0.74, 1.01) | 0.39 (0.08, 0.7) | GLP-1RAs |

Supplement Table 6D. Fasting plasma glucose (mmol/L).

| Placebo | -1.13 (-1.62, -0.64) | -1.51 (-1.72, -1.3) |
| --- | --- | --- |
| 1.13 (0.64, 1.62) | SGLT-2is | -0.38 (-0.92, 0.16) |
| 1.51 (1.3, 1.72) | 0.38 (-0.16, 0.92) | GLP-1RAs |

Supplement Table 6E. Systolic blood pressure (mmHg).

| Placebo | -1.11 (-3.14, 0.92) | -3.41 (-4.17, -2.64) |
| --- | --- | --- |
| 1.11 (-0.92, 3.14) | SGLT-2is | -2.3 (-4.45, -0.09) |
| 3.41 (2.64, 4.17) | 2.3 (0.09, 4.45) | GLP-1RAs |

Supplement Table 6F. Diastolic blood pressure (mmHg).

| Placebo | 0.37 (-1.07, 1.83) | -0.83 (-1.27, -0.29) |
| --- | --- | --- |
| -0.37 (-1.83, 1.07) | SGLT-2is | -1.2 (-2.7, 0.34) |
| 0.83 (0.29, 1.27) | 1.2 (-0.34, 2.7) | GLP-1RAs |

Supplement Table 6G. Serious adverse events.

| Placebo | 1.34 (0.71, 2.85) | 1.27 (1.04, 1.55) |
| --- | --- | --- |
| 0.75 (0.35, 1.41) | SGLT-2is | 0.95 (0.43, 1.86) |
| 0.78 (0.64, 0.96) | 1.06 (0.54, 2.32) | GLP-1RAs |

Supplement Table 6H. Discontinue due to adverse events.

| Placebo | 1.57 (0.78, 3.33) | 2.46 (1.85, 3.31) |
| --- | --- | --- |
| 0.64 (0.3, 1.29) | SGLT-2is | 1.57 (0.7, 3.34) |
| 0.41 (0.3, 0.54) | 0.64 (0.3, 1.43) | GLP-1RAs |

### Supplement Table 7. Subgroup analysis.

Supplement Table 7A. Body weight (with diabetes).

| Placebo | -2.1  (-3.69, -0.52) | -2.27  (-3.18, -1.37) | -2.38  (-4.05, -0.71) | -1.57  (-2.25, -1.02) | -1.49  (-2.3, -0.59) | -3.03  (-3.66, -2.43) | -4.46  (-5.95, -2.99) | -4.08  (-5.42, -2.8) | -6.52  (-8.17, -4.92) |
| --- | --- | --- | --- | --- | --- | --- | --- | --- | --- |
| 2.1  (0.52, 3.69) | Canagliflozin  300mg | -0.16  (-2, 1.65) | -0.27  (-2.61, 2.04) | 0.53  (-1.21, 2.17) | 0.62  (-1.15, 2.44) | -0.93  (-2.67, 0.77) | -2.36  (-4.53, -0.2) | -1.98  (-4.07, 0.02) | -4.41  (-6.67, -2.16) |
| 2.27  (1.37, 3.18) | 0.16  (-1.65, 2) | Dapagliflozin  10mg | -0.11  (-1.98, 1.81) | 0.7  (-0.39, 1.68) | 0.78  (-0.41, 2.08) | -0.75  (-1.86, 0.33) | -2.19  (-3.9, -0.45) | -1.81  (-3.4, -0.24) | -4.25  (-6.1, -2.39) |
| 2.38  (0.71, 4.05) | 0.27  (-2.04, 2.61) | 0.11  (-1.81, 1.98) | Empagliflozin  10mg | 0.81  (-1.04, 2.54) | 0.9  (-0.97, 2.83) | -0.65  (-2.44, 1.11) | -2.09  (-4.36, 0.16) | -1.71  (-3.86, 0.37) | -4.14  (-6.5, -1.85) |
| 1.57  (1.02, 2.25) | -0.53  (-2.17, 1.21) | -0.7  (-1.68, 0.39) | -0.81  (-2.54, 1.04) | Exenatide  10ug | 0.09  (-0.88, 1.23) | -1.45  (-2.27, -0.56) | -2.89  (-4.43, -1.26) | -2.51  (-3.91, -1.05) | -4.94  (-6.66, -3.17) |
| 1.49  (0.59, 2.3) | -0.62  (-2.44, 1.15) | -0.78  (-2.08, 0.41) | -0.9  (-2.83, 0.97) | -0.09  (-1.23, 0.88) | Dulaglutide  1.5mg | -1.54  (-2.63, -0.53) | -2.98  (-4.72, -1.31) | -2.6  (-4.23, -1.09) | -5.02  (-6.92, -3.24) |
| 3.03  (2.43, 3.66) | 0.93  (-0.77, 2.67) | 0.75  (-0.33, 1.86) | 0.65  (-1.11, 2.44) | 1.45  (0.56, 2.27) | 1.54  (0.53, 2.63) | Liraglutide  1.8mg | -1.43  (-2.88, 0.06) | -1.05  (-2.5, 0.36) | -3.49  (-5.23, -1.78) |
| 4.46  (2.99, 5.95) | 2.36  (0.2, 4.53) | 2.19  (0.45, 3.9) | 2.09  (-0.16, 4.36) | 2.89  (1.26, 4.43) | 2.98  (1.31, 4.72) | 1.43  (-0.06, 2.88) | Liraglutide  3.0mg | 0.38  (-1.61, 2.38) | -2.04  (-4.29, 0.12) |
| 4.08  (2.8, 5.42) | 1.98  (-0.02, 4.07) | 1.81  (0.24, 3.4) | 1.71  (-0.37, 3.86) | 2.51  (1.05, 3.91) | 2.6  (1.09, 4.23) | 1.05  (-0.36, 2.5) | -0.38  (-2.38, 1.61) | Semaglutide  1.0mg | -2.43  (-4.08, -0.78) |
| 6.52  (4.92, 8.17) | 4.41  (2.16, 6.67) | 4.25  (2.39, 6.1) | 4.14  (1.85, 6.5) | 4.94  (3.17, 6.66) | 5.02  (3.24, 6.92) | 3.49  (1.78, 5.23) | 2.04  (-0.12, 4.29) | 2.43  (0.78, 4.08) | Semaglutide  2.4mg |

Supplement Table 7A. Body weight (without diabetes).

| Placebo | -1.32  (-3.88, 1.28) | -1.83  (-5.21, 1.34) | -3.65  (-14.07, 6.65) | 1.13  (-1.49, 3.75) | -3.2  (-5.61, -0.78) | -4.74  (-5.89, -3.61) | -12.72  (-14.38, -11.11) |
| --- | --- | --- | --- | --- | --- | --- | --- |
| 1.32  (-1.28, 3.88) | Canagliflozin  300mg | -0.53  (-4.82, 3.59) | -2.27  (-13.04, 8.39) | 2.46  (-1.25, 6.07) | -1.9  (-5.42, 1.65) | -3.42  (-6.28, -0.61) | -11.4  (-14.47, -8.3) |
| 1.83  (-1.34, 5.21) | 0.53  (-3.59, 4.82) | Dapagliflozin  10mg | -1.8  (-12.71, 9.14) | 2.98  (-1.11, 7.25) | -1.37  (-5.35, 2.84) | -2.91  (-6.31, 0.68) | -10.88  (-14.46, -7.06) |
| 3.65  (-6.65, 14.07) | 2.27  (-8.39, 13.04) | 1.8  (-9.14, 12.71) | Empagliflozin  10mg | 4.79  (-5.81, 15.47) | 0.49  (-10.1, 11.02) | -1.07  (-11.49, 9.34) | -9.07  (-19.47, 1.43) |
| -1.13  (-3.75, 1.49) | -2.46  (-6.07, 1.25) | -2.98  (-7.25, 1.11) | -4.79  (-15.47, 5.81) | Exenatide  10ug | -4.33  (-7.84, -0.79) | -5.86  (-8.74, -3.04) | -13.84  (-16.92, -10.78) |
| 3.2  (0.78, 5.61) | 1.9  (-1.65, 5.42) | 1.37  (-2.84, 5.35) | -0.49  (-11.02, 10.1) | 4.33  (0.79, 7.84) | Liraglutide  1.8mg | -1.54  (-4.1, 0.95) | -9.52  (-12.42, -6.6) |
| 4.74  (3.61, 5.89) | 3.42  (0.61, 6.28) | 2.91  (-0.68, 6.31) | 1.07  (-9.34, 11.49) | 5.86  (3.04, 8.74) | 1.54  (-0.95, 4.1) | Liraglutide  3.0mg | -7.97  (-9.88, -6.04) |
| 12.72  (11.11, 14.38) | 11.4  (8.3, 14.47) | 10.88  (7.06, 14.46) | 9.07  (-1.43, 19.47) | 13.84  (10.78, 16.92) | 9.52  (6.6, 12.42) | 7.97  (6.04, 9.88) | Semaglutide  2.4mg |

Supplement Table 7B. Reaching ≥ 5% weight loss (with diabetes).

| Placebo | 10.59  (0.67, 187.38) | 1.92  (0.14, 24.26) | 4.29  (1.23, 18.88) | 6.95  (0.74, 74) | 6.66  (1.17, 47.45) | 7.89  (0.69, 93.14) |
| --- | --- | --- | --- | --- | --- | --- |
| 0.09  (0.01, 1.49) | Canagliflozin  300mg | 0.18  (0, 7.39) | 0.41  (0.02, 9.8) | 0.65  (0.02, 24.85) | 0.63  (0.02, 19.74) | 0.74  (0.02, 29.24) |
| 0.52  (0.04, 7.25) | 5.52  (0.14, 273.2) | Exenatide  10ug | 2.21  (0.13, 50.87) | 3.63  (0.13, 124.66) | 3.51  (0.16, 104.75) | 4.19  (0.13, 157.43) |
| 0.23  (0.05, 0.82) | 2.46  (0.1, 54.43) | 0.45  (0.02, 7.42) | Liraglutide  1.8mg | 1.63  (0.16, 14.88) | 1.56  (0.15, 15.31) | 1.86  (0.1, 26.23) |
| 0.14  (0.01, 1.36) | 1.53  (0.04, 58.1) | 0.28  (0.01, 7.64) | 0.61  (0.07, 6.39) | Liraglutide  3.0mg | 0.96  (0.05, 19.75) | 1.13  (0.04, 32.6) |
| 0.15  (0.02, 0.86) | 1.58  (0.05, 45.65) | 0.28  (0.01, 6.23) | 0.64  (0.07, 6.8) | 1.04  (0.05, 19.1) | Semaglutide  1.0mg | 1.2  (0.1, 12.86) |
| 0.13  (0.01, 1.46) | 1.36  (0.03, 56.48) | 0.24  (0.01, 7.95) | 0.54  (0.04, 9.82) | 0.88  (0.03, 24.72) | 0.84  (0.08, 10.23) | Semaglutide  2.4mg |

Supplement Table 7B. Reaching ≥ 5% weight loss (without diabetes).

| Placebo | 1.62  (0.61, 4.38) | 2.36  (0.65, 8.68) | 3.97  (1.74, 11) | 4.7  (3.2, 7.09) | 11.95  (6.97, 21.57) |
| --- | --- | --- | --- | --- | --- |
| 0.62  (0.23, 1.64) | Canagliflozin  300mg | 1.46  (0.29, 7.38) | 2.45  (0.69, 10.11) | 2.9  (1.01, 8.42) | 7.4  (2.44, 23.54) |
| 0.42  (0.12, 1.54) | 0.68 (0.14, 3.48) | Exenatide  10ug | 1.69  (0.37, 8.62) | 2  (0.51, 7.79) | 5.07  (1.24, 21.19) |
| 0.25  (0.09, 0.58) | 0.41  (0.1, 1.46) | 0.59  (0.12, 2.69) | Liraglutide  1.8mg | 1.18  (0.43, 2.81) | 3.01  (0.96, 8.13) |
| 0.21  (0.14, 0.31) | 0.34  (0.12, 0.99) | 0.5  (0.13, 1.98) | 0.85  (0.36, 2.35) | Liraglutide  3.0mg | 2.54  (1.32, 4.96) |
| 0.08  (0.05, 0.14) | 0.14  (0.04, 0.41) | 0.2  (0.05, 0.81) | 0.33  (0.12, 1.04) | 0.39  (0.2, 0.76) | Semaglutide  2.4mg |

Supplement Table 7C. Systolic blood pressure (with diabetes).

| Placebo | -0.53  (-4.46, 3.63) | -0.88  (-5.63, 3.78) | -3.77  (-6.43, -0.82) | -1.32  (-3.97, 1.97) | -3.31  (-5.16, -1.66) | -2.48  (-5.96, 0.84) | -2.67  (-5.96, 0.42) | -3.51  (-7.31, -0.02) |
| --- | --- | --- | --- | --- | --- | --- | --- | --- |
| 0.53  (-3.63, 4.46) | Dapagliflozin  10mg | -0.33  (-6.67, 5.71) | -3.22  (-8.13, 1.59) | -0.78  (-5.55, 4.4) | -2.77  (-7.36, 1.42) | -1.92  (-7.34, 3.01) | -2.12  (-7.45, 2.92) | -2.96  (-8.6, 2.21) |
| 0.88  (-3.78, 5.63) | 0.33  (-5.71, 6.67) | Empagliflozin  10mg | -2.92  (-8.19, 2.76) | -0.44  (-5.68, 5.42) | -2.44  (-7.64, 2.51) | -1.61  (-7.46, 4.2) | -1.79  (-7.67, 3.89) | -2.62  (-8.71, 3.16) |
| 3.77  (0.82, 6.43) | 3.22  (-1.59, 8.13) | 2.92  (-2.76, 8.19) | Exenatide  10ug | 2.43  (-1.4, 6.68) | 0.45  (-3.13, 3.54) | 1.3  (-3.32, 5.44) | 1.07  (-3.44, 5.11) | 0.26  (-4.61, 4.52) |
| 1.32  (-1.97, 3.97) | 0.78  (-4.4, 5.55) | 0.44  (-5.42, 5.68) | -2.43  (-6.68, 1.4) | Dulaglutide  1.5mg | -1.97  (-5.94, 1.07) | -1.12  (-6.18, 2.87) | -1.35  (-6.1, 2.67) | -2.19  (-7.49, 2.04) |
| 3.31  (1.66, 5.16) | 2.77  (-1.42, 7.36) | 2.44  (-2.51, 7.64) | -0.45  (-3.54, 3.13) | 1.97  (-1.07, 5.94) | Liraglutide  1.8mg | 0.81  (-2.48, 4.29) | 0.62  (-2.93, 4.31) | -0.23  (-4.18, 3.84) |
| 2.48  (-0.84, 5.96) | 1.92  (-3.01, 7.34) | 1.61  (-4.2, 7.46) | -1.3  (-5.44, 3.32) | 1.12  (-2.87, 6.18) | -0.81  (-4.29, 2.48) | Liraglutide  3.0mg | -0.18  (-4.92, 4.38) | -1.03  (-6.04, 3.95) |
| 2.67  (-0.42, 5.96) | 2.12  (-2.92, 7.45) | 1.79  (-3.89, 7.67) | -1.07  (-5.11, 3.44) | 1.35  (-2.67, 6.1) | -0.62  (-4.31, 2.93) | 0.18  (-4.38, 4.92) | Semaglutide  1.0mg | -0.85  (-4.5, 2.77) |
| 3.51  (0.02, 7.31) | 2.96  (-2.21, 8.6) | 2.62  (-3.16, 8.71) | -0.26  (-4.52, 4.61) | 2.19  (-2.04, 7.49) | 0.23  (-3.84, 4.18) | 1.03  (-3.95, 6.04) | 0.85  (-2.77, 4.5) | Semaglutide  2.4mg |

Supplement Table 7C. Systolic blood pressure (without diabetes).

| Placebo | -1.1  (-4.09, 1.93) | -7.04  (-17.98, 4.13) | -1.56  (-9.92, 6.85) | -2.38  (-5.24, 0.31) | -2.71  (-3.84, -1.51) | -5.23  (-6.61, -3.79) |
| --- | --- | --- | --- | --- | --- | --- |
| 1.1  (-1.93, 4.09) | Canagliflozin  300mg | -5.96  (-17.26, 5.51) | -0.46  (-9.45, 8.38) | -1.28  (-5.43, 2.76) | -1.6  (-4.82, 1.62) | -4.1  (-7.46, -0.83) |
| 7.04  (-4.13, 17.98) | 5.96  (-5.51, 17.26) | Dapagliflozin  10mg | 5.41  (-8.14, 19.37) | 4.67  (-6.84, 16.01) | 4.35  (-6.82, 15.25) | 1.84  (-9.41, 12.8) |
| 1.56  (-6.85, 9.92) | 0.46  (-8.38, 9.45) | -5.41  (-19.37, 8.14) | Empagliflozin  10mg | -0.8  (-9.73, 8.08) | -1.15  (-9.69, 7.27) | -3.65  (-12.35, 4.84) |
| 2.38  (-0.31, 5.24) | 1.28  (-2.76, 5.43) | -4.67  (-16.01, 6.84) | 0.8  (-8.08, 9.73) | Liraglutide  1.8mg | -0.34  (-3.06, 2.58) | -2.86  (-5.86, 0.26) |
| 2.71  (1.51, 3.84) | 1.6  (-1.62, 4.82) | -4.35  (-15.25, 6.82) | 1.15  (-7.27, 9.69) | 0.34  (-2.58, 3.06) | Liraglutide  3.0mg | -2.52  (-4.32, -0.78) |
| 5.23  (3.79, 6.61) | 4.1  (0.83, 7.46) | -1.84  (-12.8, 9.41) | 3.65  (-4.84, 12.35) | 2.86  (-0.26, 5.86) | 2.52  (0.78, 4.32) | Semaglutide  2.4mg |

Supplement Table 7D. Diastolic blood pressure (with diabetes).

| Placebo | -0.34  (-2.1, 1.88) | -1.2  (-2.77, 0.51) | -0.21  (-1.77, 1.76) | -0.33  (-1.48, 0.82) | -0.26  (-2.29, 1.8) | 0.47  (-1.21, 2.39) | -1.06  (-2.68, 0.5) |
| --- | --- | --- | --- | --- | --- | --- | --- |
| 0.34  (-1.88, 2.1) | Dapagliflozin  10mg | -0.87  (-3.53, 1.59) | 0.13  (-2.44, 2.74) | -0.01  (-2.47, 2.09) | 0.09  (-3.08, 2.69) | 0.79  (-1.98, 3.31) | -0.72  (-3.57, 1.59) |
| 1.2  (-0.51, 2.77) | 0.87  (-1.59, 3.53) | Exenatide  10ug | 1.01  (-1.28, 3.47) | 0.88  (-1.19, 2.79) | 0.94  (-1.77, 3.51) | 1.68  (-0.73, 4.08) | 0.15  (-2.27, 2.32) |
| 0.21  (-1.76, 1.77) | -0.13  (-2.74, 2.44) | -1.01  (-3.47, 1.28) | Dulaglutide  1.5mg | -0.14  (-2.42, 1.79) | -0.05  (-3.02, 2.42) | 0.67  (-1.93, 3) | -0.85  (-3.53, 1.27) |
| 0.33  (-0.82, 1.48) | 0.01  (-2.09, 2.47) | -0.88  (-2.79, 1.19) | 0.14  (-1.79, 2.42) | Liraglutide  1.8mg | 0.07  (-1.96, 2.05) | 0.8  (-1.25, 3.01) | -0.73  (-2.72, 1.18) |
| 0.26  (-1.8, 2.29) | -0.09  (-2.69, 3.08) | -0.94  (-3.51, 1.77) | 0.05  (-2.42, 3.02) | -0.07  (-2.05, 1.96) | Liraglutide  3.0mg | 0.71  (-1.83, 3.63) | -0.79  (-3.44, 1.75) |
| -0.47  (-2.39, 1.21) | -0.79  (-3.31, 1.98) | -1.68  (-4.08, 0.73) | -0.67  (-3, 1.93) | -0.8  (-3.01, 1.25) | -0.71  (-3.63, 1.83) | Semaglutide  1.0mg | -1.51  (-3.66, 0.24) |
| 1.06  (-0.5, 2.68) | 0.72  (-1.59, 3.57) | -0.15  (-2.32, 2.27) | 0.85  (-1.27, 3.53) | 0.73  (-1.18, 2.72) | 0.79  (-1.75, 3.44) | 1.51  (-0.24, 3.66) | Semaglutide  2.4mg |

Supplement Table 7D. Diastolic blood pressure (without diabetes).

| Placebo | 0.33  (-1.53, 2.22) | 2.92  (-3.51, 9.78) | 0.34  (-1.51, 2.2) | -0.55  (-1.27, 0.24) | -1.95  (-2.96, -1.08) |
| --- | --- | --- | --- | --- | --- |
| -0.33  (-2.22, 1.53) | Canagliflozin  300mg | 2.61  (-4.08, 9.69) | 0  (-2.6, 2.67) | -0.88  (-2.91, 1.18) | -2.27  (-4.42, -0.19) |
| -2.92  (-9.78, 3.51) | -2.61  (-9.69, 4.08) | Dapagliflozin  10mg | -2.59  (-9.71, 4.2) | -3.45  (-10.39, 2.96) | -4.86  (-11.78, 1.6) |
| -0.34  (-2.2, 1.51) | 0  (-2.67, 2.6) | 2.59  (-4.2, 9.71) | Liraglutide  1.8mg | -0.88  (-2.75, 1) | -2.28  (-4.42, -0.27) |
| 0.55  (-0.24, 1.27) | 0.88  (-1.18, 2.91) | 3.45  (-2.96, 10.39) | 0.88  (-1, 2.75) | Liraglutide  3.0mg | -1.39  (-2.68, -0.32) |
| 1.95  (1.08, 2.96) | 2.27  (0.19, 4.42) | 4.86  (-1.6, 11.78) | 2.28  (0.27, 4.42) | 1.39  (0.32, 2.68) | Semaglutide  2.4mg |

Supplement Table 7E. Serious adverse events (with diabetes).

| Placebo | 1.21  (0.03, 51.45) | 4  (0.78, 32.64) | 0.98  (0.42, 2.35) | 0.81  (0.43, 1.58) | 1.29  (0.56, 3.09) | 0.91  (0.6, 1.44) | 1.18  (0.57, 2.44) | 0.74  (0.36, 1.4) | 1.04  (0.49, 2.17) |
| --- | --- | --- | --- | --- | --- | --- | --- | --- | --- |
| 0.82  (0.02, 32.87) | Canagliflozin  300mg | 3.51  (0.06, 197.61) | 0.81  (0.02, 34.14) | 0.67  (0.01, 27.23) | 1.1  (0.02, 44.98) | 0.76  (0.02, 31.57) | 0.97  (0.02, 41.44) | 0.6  (0.01, 24.94) | 0.84  (0.02, 36.19) |
| 0.25  (0.03, 1.28) | 0.29  (0.01, 16.7) | Dapagliflozin  10mg | 0.24  (0.03, 1.59) | 0.2  (0.02, 1.14) | 0.31  (0.04, 2.01) | 0.22  (0.03, 1.26) | 0.29  (0.03, 1.8) | 0.18  (0.02, 1.07) | 0.26  (0.03, 1.58) |
| 1.02  (0.43, 2.4) | 1.24  (0.03, 55.93) | 4.12  (0.63, 39.12) | Empagliflozin  10mg | 0.82  (0.28, 2.45) | 1.32  (0.39, 4.26) | 0.93  (0.34, 2.47) | 1.2  (0.39, 3.74) | 0.75  (0.24, 2.12) | 1.06  (0.34, 3.13) |
| 1.24  (0.63, 2.31) | 1.49  (0.04, 72.42) | 5  (0.88, 43.71) | 1.22  (0.41, 3.55) | Exenatide  10ug | 1.6  (0.54, 4.54) | 1.14  (0.51, 2.43) | 1.46  (0.54, 3.82) | 0.91  (0.33, 2.2) | 1.29  (0.45, 3.26) |
| 0.77  (0.32, 1.78) | 0.91  (0.02, 47.77) | 3.19  (0.5, 27.34) | 0.76  (0.23, 2.57) | 0.62  (0.22, 1.84) | Dulaglutide  1.5mg | 0.71  (0.27, 1.82) | 0.91  (0.29, 2.79) | 0.56  (0.19, 1.64) | 0.79  (0.26, 2.5) |
| 1.09  (0.69, 1.68) | 1.32  (0.03, 56.09) | 4.46  (0.8, 36.75) | 1.08  (0.4, 2.91) | 0.88  (0.41, 1.95) | 1.4  (0.55, 3.73) | Liraglutide  1.8mg | 1.29  (0.62, 2.69) | 0.81  (0.34, 1.74) | 1.14  (0.47, 2.63) |
| 0.85  (0.41, 1.76) | 1.03  (0.02, 46.65) | 3.47  (0.56, 32) | 0.84  (0.27, 2.55) | 0.69  (0.26, 1.86) | 1.1  (0.36, 3.46) | 0.78  (0.37, 1.61) | Liraglutide  3.0mg | 0.62  (0.22, 1.63) | 0.88  (0.31, 2.42) |
| 1.36  (0.71, 2.8) | 1.67  (0.04, 72.03) | 5.5  (0.93, 49.95) | 1.34  (0.47, 4.21) | 1.1  (0.45, 2.99) | 1.78  (0.61, 5.26) | 1.24  (0.58, 2.92) | 1.61  (0.61, 4.48) | Semaglutide  1.0mg | 1.41  (0.69, 3.07) |
| 0.96  (0.46, 2.03) | 1.19  (0.03, 53.92) | 3.89  (0.63, 34.87) | 0.95  (0.32, 2.97) | 0.78  (0.31, 2.2) | 1.26  (0.4, 3.87) | 0.88  (0.38, 2.12) | 1.13  (0.41, 3.27) | 0.71  (0.33, 1.44) | Semaglutide  2.4mg |

Supplement Table 7F. Discontinuation due to adverse events (with diabetes).

| Placebo | 1.02  (0.06, 17.59) | 2.55  (0.38, 23.34) | 1  (0.13, 7.56) | 2.83  (1.26, 6.5) | 1.17  (0.39, 4.82) | 1.88  (0.81, 4) | 2.49  (0.45, 13.24) | 3.04  (0.76, 15.62) | 3.01  (0.53, 19.21) |
| --- | --- | --- | --- | --- | --- | --- | --- | --- | --- |
| 0.98  (0.06, 16.71) | Canagliflozin  300mg | 2.5  (0.08, 92.17) | 0.97  (0.03, 33.63) | 2.8  (0.14, 51.67) | 1.15  (0.06, 28.83) | 1.84  (0.09, 35.51) | 2.39  (0.09, 63.6) | 3.02  (0.13, 80.16) | 3  (0.11, 88.59) |
| 0.39  (0.04, 2.67) | 0.4  (0.01, 12.43) | Dapagliflozin  10mg | 0.39  (0.02, 5.95) | 1.11  (0.11, 8.65) | 0.47  (0.04, 4.91) | 0.74  (0.06, 5.62) | 0.96  (0.06, 11.49) | 1.2  (0.09, 14.91) | 1.19  (0.07, 16.36) |
| 1  (0.13, 7.63) | 1.03  (0.03, 32.94) | 2.54  (0.17, 52.56) | Empagliflozin  10mg | 2.8  (0.32, 26.25) | 1.16  (0.13, 15.54) | 1.88  (0.2, 15.69) | 2.46  (0.17, 34.05) | 3.02  (0.27, 43.53) | 3.02  (0.22, 48.76) |
| 0.35  (0.15, 0.8) | 0.36  (0.02, 7.12) | 0.9  (0.12, 9.49) | 0.36  (0.04, 3.11) | Exenatide  10ug | 0.41  (0.11, 2.12) | 0.67  (0.2, 1.97) | 0.88  (0.13, 5.55) | 1.07  (0.22, 6.84) | 1.06  (0.15, 8.35) |
| 0.86  (0.21, 2.58) | 0.87  (0.03, 18.09) | 2.14  (0.2, 24.09) | 0.86  (0.06, 7.94) | 2.41  (0.47, 9.44) | Dulaglutide  1.5mg | 1.59  (0.29, 6.17) | 2.14  (0.21, 14.26) | 2.63  (0.35, 17.22) | 2.62  (0.27, 20.53) |
| 0.53  (0.25, 1.24) | 0.54  (0.03, 11.17) | 1.36  (0.18, 15.48) | 0.53  (0.06, 4.95) | 1.5  (0.51, 5.03) | 0.63  (0.16, 3.49) | Liraglutide  1.8mg | 1.31  (0.25, 7.27) | 1.62  (0.33, 10.99) | 1.62  (0.24, 12.89) |
| 0.4  (0.08, 2.22) | 0.42  (0.02, 11.56) | 1.04  (0.09, 17.86) | 0.41  (0.03, 5.72) | 1.13  (0.18, 7.6) | 0.47  (0.07, 4.82) | 0.76  (0.14, 3.97) | Liraglutide  3.0mg | 1.22  (0.15, 13.96) | 1.22  (0.11, 16.64) |
| 0.33  (0.06, 1.32) | 0.33  (0.01, 7.67) | 0.83  (0.07, 11.17) | 0.33  (0.02, 3.68) | 0.94  (0.15, 4.62) | 0.38  (0.06, 2.87) | 0.62  (0.09, 3.02) | 0.82  (0.07, 6.82) | Semaglutide  1.0mg | 1.01  (0.15, 5.62) |
| 0.33  (0.05, 1.89) | 0.33  (0.01, 9.4) | 0.84  (0.06, 14.56) | 0.33  (0.02, 4.62) | 0.94  (0.12, 6.56) | 0.38  (0.05, 3.69) | 0.62  (0.08, 4.09) | 0.82  (0.06, 8.97) | 0.99  (0.18, 6.66) | Semaglutide  2.4mg |

Supplement Table 7F. Discontinuation due to adverse events (without diabetes).

| Placebo | 3.03  (0.89, 13.75) | 1.4  (0.2, 10.33) | 0.72  (0.09, 5.16) | 4.36  (1.02, 26.58) | 4.78  (1.54, 17.04) | 2.98  (1.81, 4.67) | 1.69  (0.85, 2.89) |
| --- | --- | --- | --- | --- | --- | --- | --- |
| 0.33  (0.07, 1.12) | Canagliflozin  300mg | 0.45  (0.04, 4.88) | 0.23  (0.02, 2.36) | 1.44  (0.18, 12.19) | 1.55  (0.24, 8.64) | 0.98  (0.19, 3.6) | 0.55  (0.1, 2.09) |
| 0.72  (0.1, 5.07) | 2.2  (0.2, 25.24) | Dapagliflozin  10mg | 0.52  (0.03, 8.32) | 3.1  (0.27, 42.81) | 3.41  (0.33, 34.42) | 2.13  (0.27, 15.67) | 1.2  (0.14, 8.82) |
| 1.4  (0.19, 10.87) | 4.3  (0.42, 52.73) | 1.94  (0.12, 35.37) | Empagliflozin  10mg | 6.28 (0.52, 94.88) | 6.57  (0.7, 75.07) | 4.13  (0.53, 34.07) | 2.32  (0.29, 19.26) |
| 0.23  (0.04, 0.98) | 0.69  (0.08, 5.46) | 0.32  (0.02, 3.75) | 0.16  (0.01, 1.91) | Exenatide  10ug | 1.09  (0.14, 7.54) | 0.69  (0.1, 3.17) | 0.39  (0.05, 1.79) |
| 0.21  (0.06, 0.65) | 0.64  (0.12, 4.18) | 0.29  (0.03, 3.02) | 0.15  (0.01, 1.43) | 0.92  (0.13, 7.41) | Liraglutide  1.8mg | 0.62  (0.17, 1.97) | 0.36  (0.08, 1.2) |
| 0.34  (0.21, 0.55) | 1.02  (0.28, 5.21) | 0.47  (0.06, 3.71) | 0.24  (0.03, 1.89) | 1.46  (0.32, 9.55) | 1.61  (0.51, 5.86) | Liraglutide  3.0mg | 0.57  (0.26, 1.13) |
| 0.59  (0.35, 1.17) | 1.82  (0.48, 9.91) | 0.84  (0.11, 6.97) | 0.43  (0.05, 3.46) | 2.6  (0.56, 19.02) | 2.81  (0.83, 12.37) | 1.76  (0.89, 3.82) | Semaglutide  2.4mg |

### Supplement Table 8. Sensitive analysis.

Supplement Table 8A. Body weight.

| Placebo | -1.59  (-3.55, 0.37) | -2.23  (-3.92, -0.54) | -1.92  (-5.1, 1.31) | -1.03  (-2.18, 0.09) | -1.04  (-2.96, 0.9) | -3.24  (-4.43, -2.04) | -4.62  (-5.66, -3.59) | -5.65  (-7.95, -3.37) | -11.51  (-12.92, -10.12) |
| --- | --- | --- | --- | --- | --- | --- | --- | --- | --- |
| 1.59  (-0.37, 3.55) | Canagliflozin  300mg | -0.63  (-3.19, 1.93) | -0.33  (-4.07, 3.45) | 0.57  (-1.72, 2.78) | 0.56  (-2.16, 3.33) | -1.65  (-3.98, 0.63) | -3.03  (-5.23, -0.82) | -4.07  (-7.06, -1.05) | -9.92  (-12.34, -7.53) |
| 2.23  (0.54, 3.92) | 0.63  (-1.93, 3.19) | Dapagliflozin  10mg | 0.29  (-3.31, 3.94) | 1.19  (-0.74, 3.13) | 1.2  (-1.39, 3.77) | -1.01  (-3.08, 1.05) | -2.39  (-4.41, -0.42) | -3.44  (-6.22, -0.59) | -9.29  (-11.5, -7.11) |
| 1.92  (-1.31, 5.1) | 0.33  (-3.45, 4.07) | -0.29  (-3.94, 3.31) | Empagliflozin  10mg | 0.89  (-2.51, 4.27) | 0.89  (-2.89, 4.61) | -1.31  (-4.75, 2.06) | -2.69  (-6.06, 0.65) | -3.72  (-7.71, 0.17) | -9.58  (-13.1, -6.11) |
| 1.03  (-0.09, 2.18) | -0.57  (-2.78, 1.72) | -1.19  (-3.13, 0.74) | -0.89  (-4.27, 2.51) | Exenatide  10ug | -0.01  (-2.25, 2.28) | -2.2  (-3.88, -0.58) | -3.59  (-5.13, -2.06) | -4.63  (-7.18, -2.04) | -10.49  (-12.26, -8.65) |
| 1.04  (-0.9, 2.96) | -0.56  (-3.33, 2.16) | -1.20  (-3.77, 1.39) | -0.89  (-4.61, 2.89) | 0.01  (-2.28, 2.25) | Dulaglutide  1.5mg | -2.2  (-4.49, 0.06) | -3.58  (-5.79, -1.41) | -4.62  (-7.67, -1.61) | -10.48  (-12.89, -8.13) |
| 3.24  (2.04, 4.43) | 1.65  (-0.63, 3.98) | 1.01  (-1.05, 3.08) | 1.31  (-2.06, 4.75) | 2.2  (0.58, 3.88) | 2.2  (-0.06, 4.49) | Liraglutide  1.8mg | -1.39  (-2.83, 0.09) | -2.41  (-4.97, 0.2) | -8.27  (-10.08, -6.48) |
| 4.62  (3.59, 5.66) | 3.03  (0.82, 5.23) | 2.39  (0.42, 4.41) | 2.69  (-0.65, 6.06) | 3.59  (2.06, 5.13) | 3.58  (1.41, 5.79) | 1.39  (-0.09, 2.83) | Liraglutide  3.0mg | -1.04  (-3.51, 1.47) | -6.89  (-8.59, -5.23) |
| 5.65  (3.37, 7.95) | 4.07  (1.05, 7.06) | 3.44  (0.59, 6.22) | 3.72  (-0.17, 7.71) | 4.63  (2.04, 7.18) | 4.62  (1.61, 7.67) | 2.41  (-0.2, 4.97) | 1.04  (-1.47, 3.51) | Semaglutide  1.0mg | -5.85  (-8.3, -3.43) |
| 11.51  (10.12, 12.92) | 9.92  (7.53, 12.34) | 9.29  (7.11, 11.5) | 9.58  (6.11, 13.1) | 10.49  (8.65, 12.26) | 10.48  (8.13, 12.89) | 8.27  (6.48, 10.08) | 6.89  (5.23, 8.59) | 5.85  (3.43, 8.3) | Semaglutide  2.4mg |

| Placebo | 2.67  (1.26, 5.9) | 2.1  (0.94, 4.64) | 2.77  (1.65, 4.65) | 4.83  (3.44, 6.85) | 6.74  (3.4, 14.43) | 10.72  (7.04, 17.1) |
| --- | --- | --- | --- | --- | --- | --- |
| 0.37  (0.17, 0.79) | Canagliflozin  300mg | 0.78  (0.26, 2.33) | 1.03  (0.41, 2.56) | 1.81  (0.77, 4.15) | 2.53  (0.9, 7.42) | 4  (1.65, 9.76) |
| 0.48  (0.22, 1.07) | 1.28  (0.43, 3.87) | Exenatide  10ug | 1.32 (0.51, 3.43) | 2.31  (0.98, 5.58) | 3.21  (1.15, 9.8) | 5.14  (2.11, 13.02) |
| 0.36  (0.21, 0.61) | 0.97  (0.39, 2.46) | 0.76  (0.29, 1.97) | Liraglutide  1.8mg | 1.74  (1, 3.08) | 2.43  (1.05, 6.13) | 3.87  (2, 7.7) |
| 0.21  (0.15, 0.29) | 0.55  (0.24, 1.3) | 0.43  (0.18, 1.03) | 0.57  (0.32, 1) | Liraglutide  3.0mg | 1.4  (0.66, 3.18) | 2.22  (1.31, 3.86) |
| 0.15  (0.07, 0.29) | 0.39  (0.13, 1.12) | 0.31  (0.1, 0.87) | 0.41  (0.16, 0.95) | 0.72  (0.31, 1.52) | Semaglutide  1.0mg | 1.59  (0.74, 3.28) |
| 0.09  (0.06, 0.14) | 0.25  (0.1, 0.61) | 0.19  (0.08, 0.47) | 0.26  (0.13, 0.5) | 0.45  (0.26, 0.77) | 0.63  (0.3, 1.36) | Semaglutide  2.4mg |

Supplement Table 8B. Achieving at least 5% weight loss.

### Supplement Figure 1. Comparison-adjusted funnel plot for outcome.

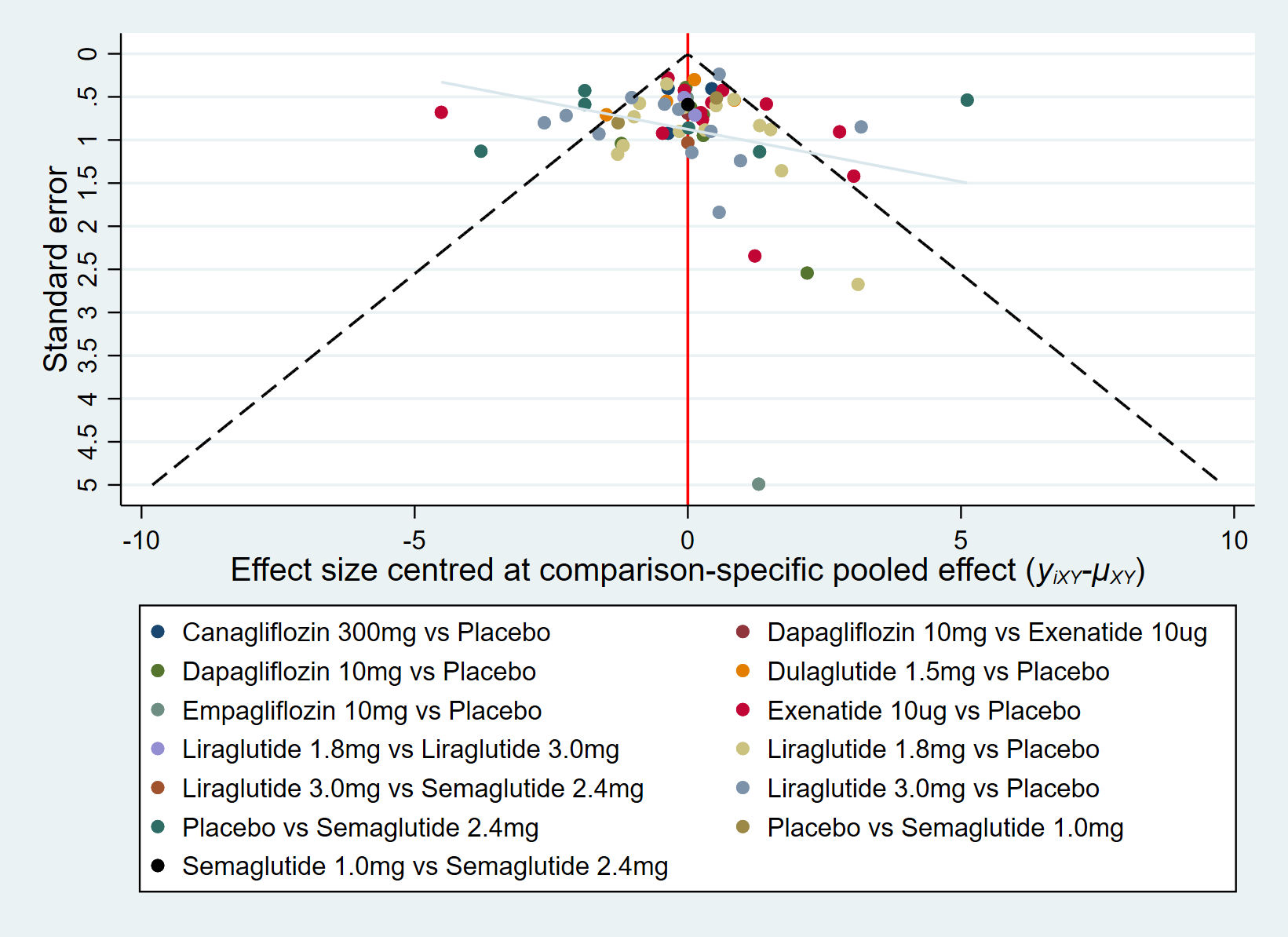

Supplement Figure 1A. Body weight.

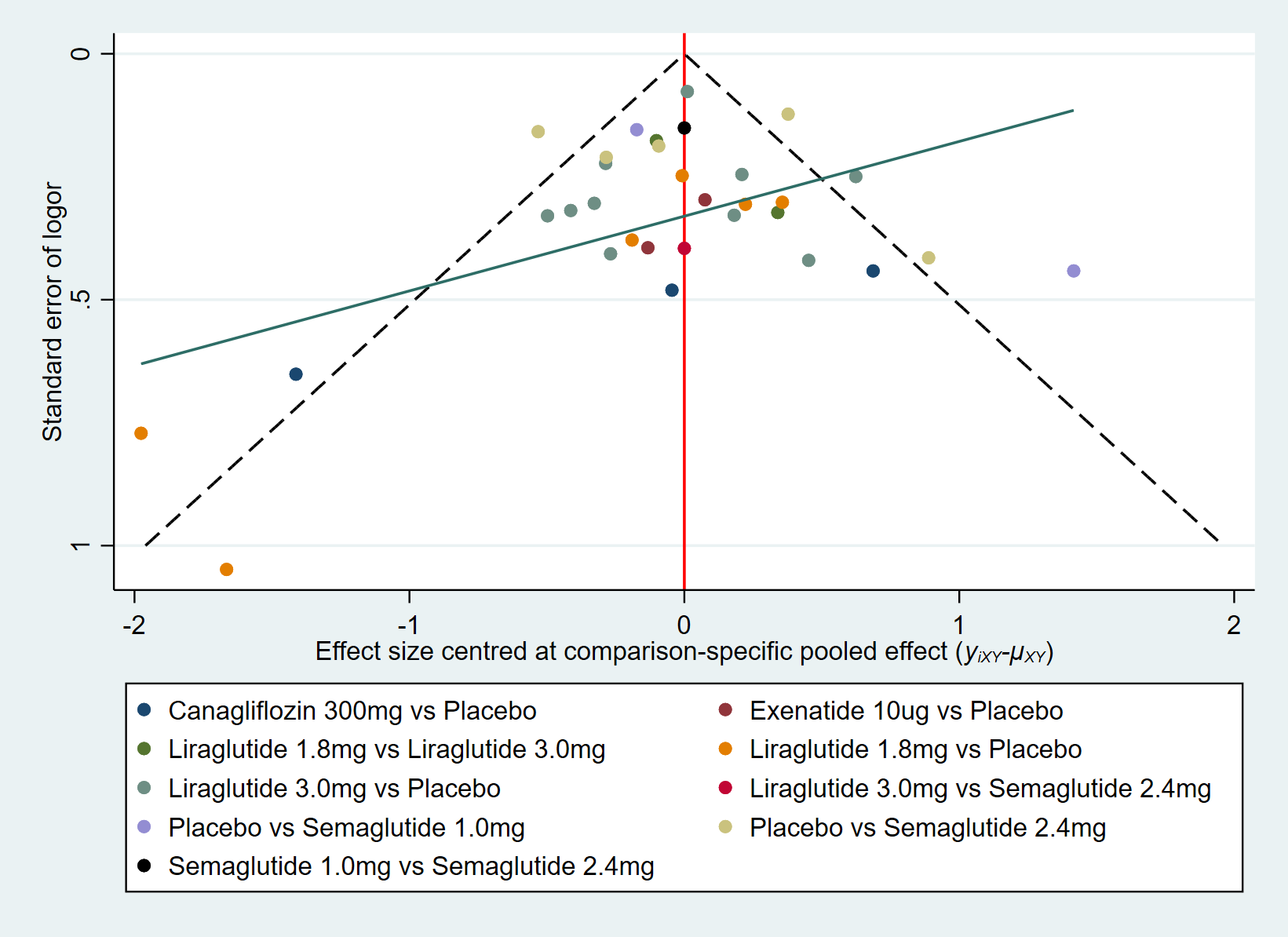

Supplement Figure 1B. Achieving at least 5% weight loss.

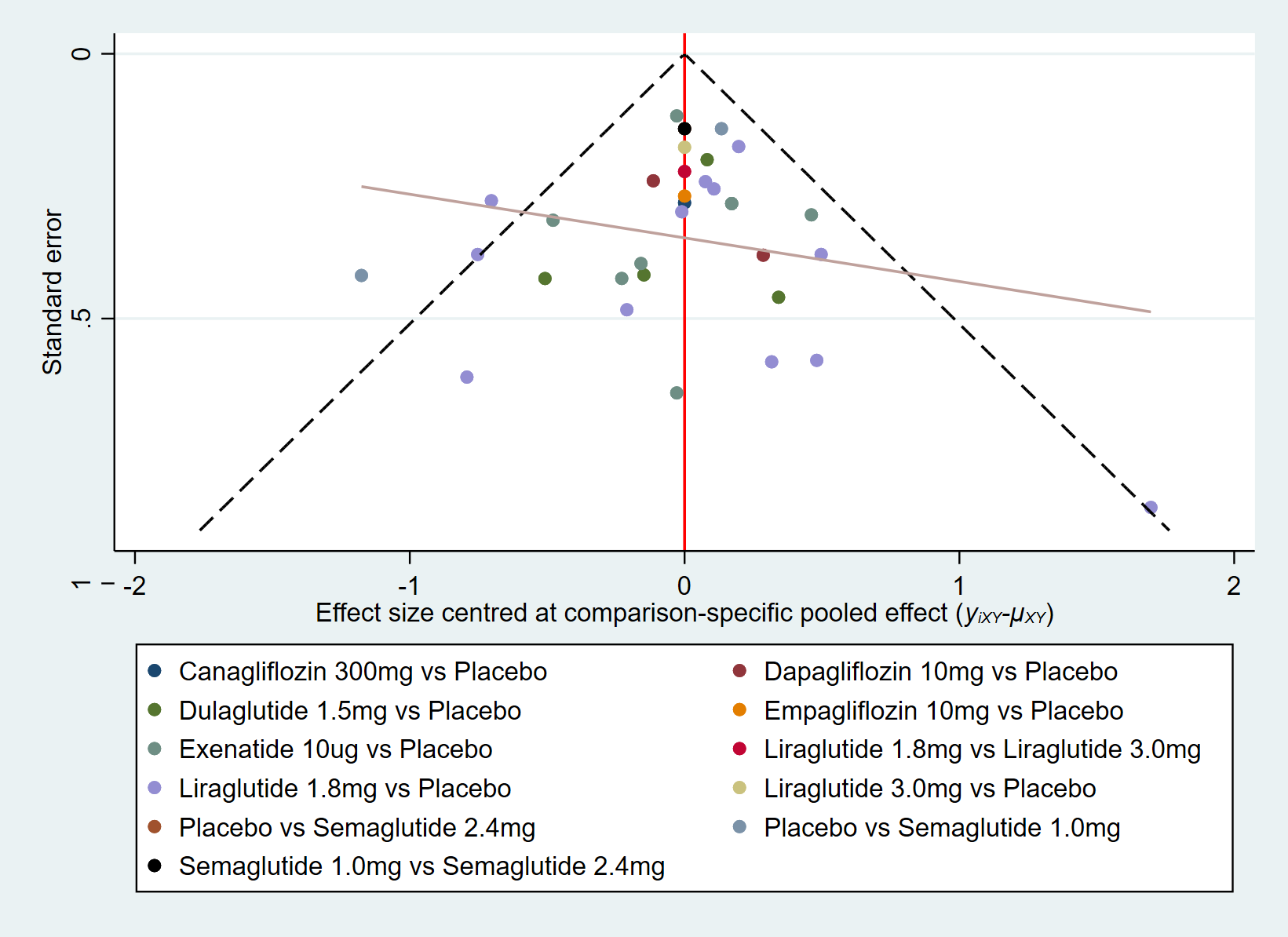

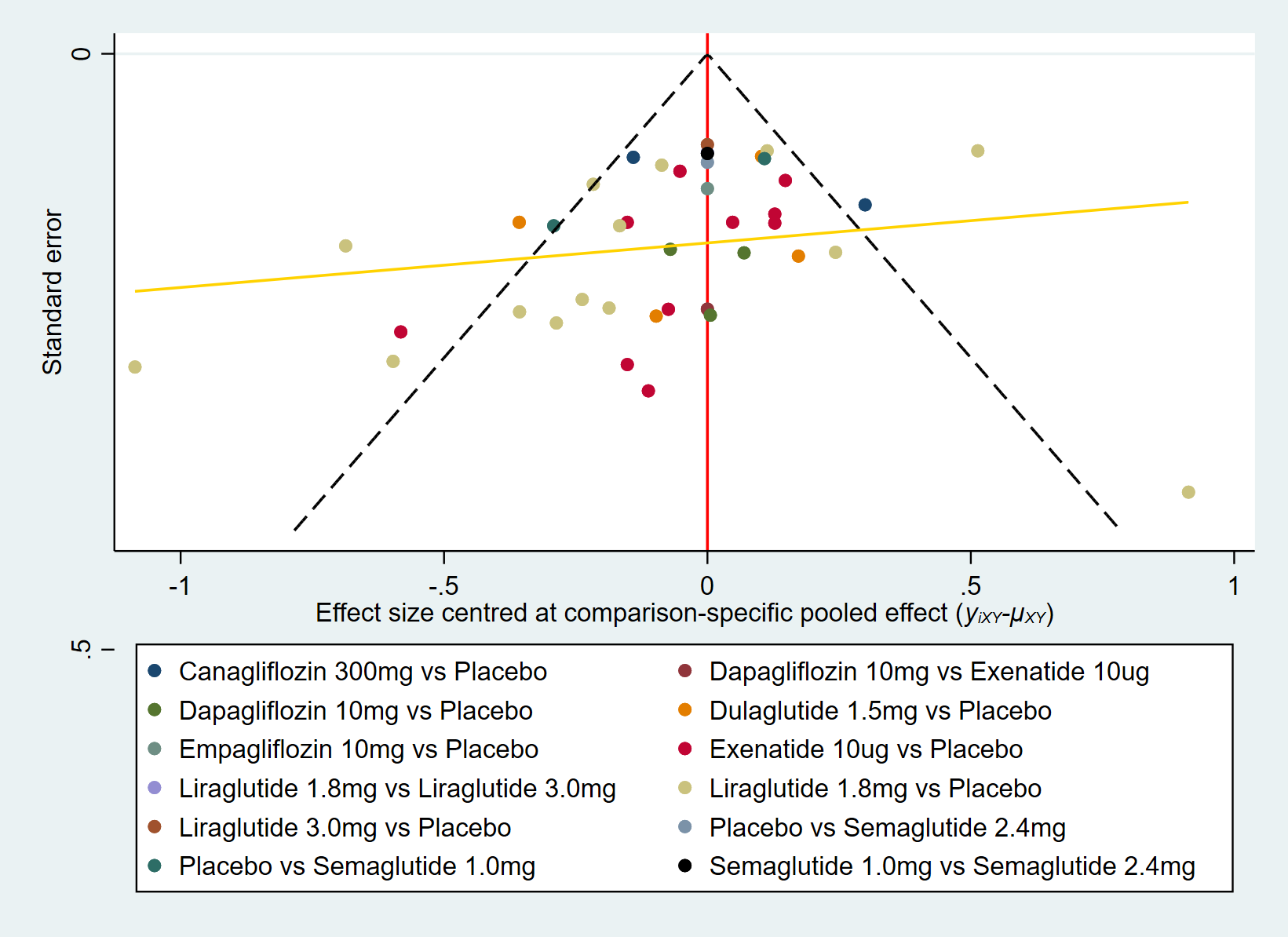

Supplement Figure 1D. Fasting plasma glucose (FPG).

Supplement Figure 1C. HbA1c.

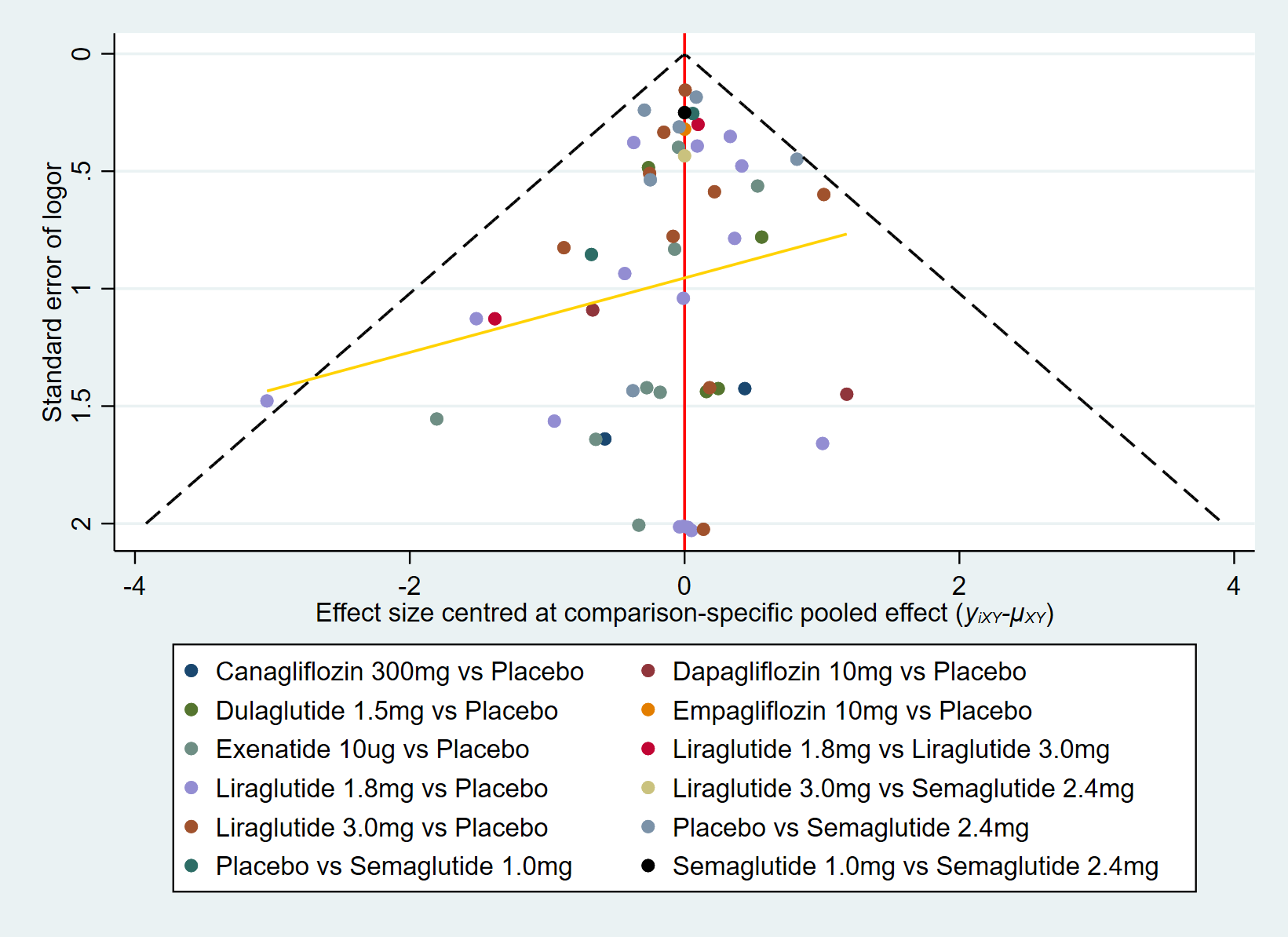

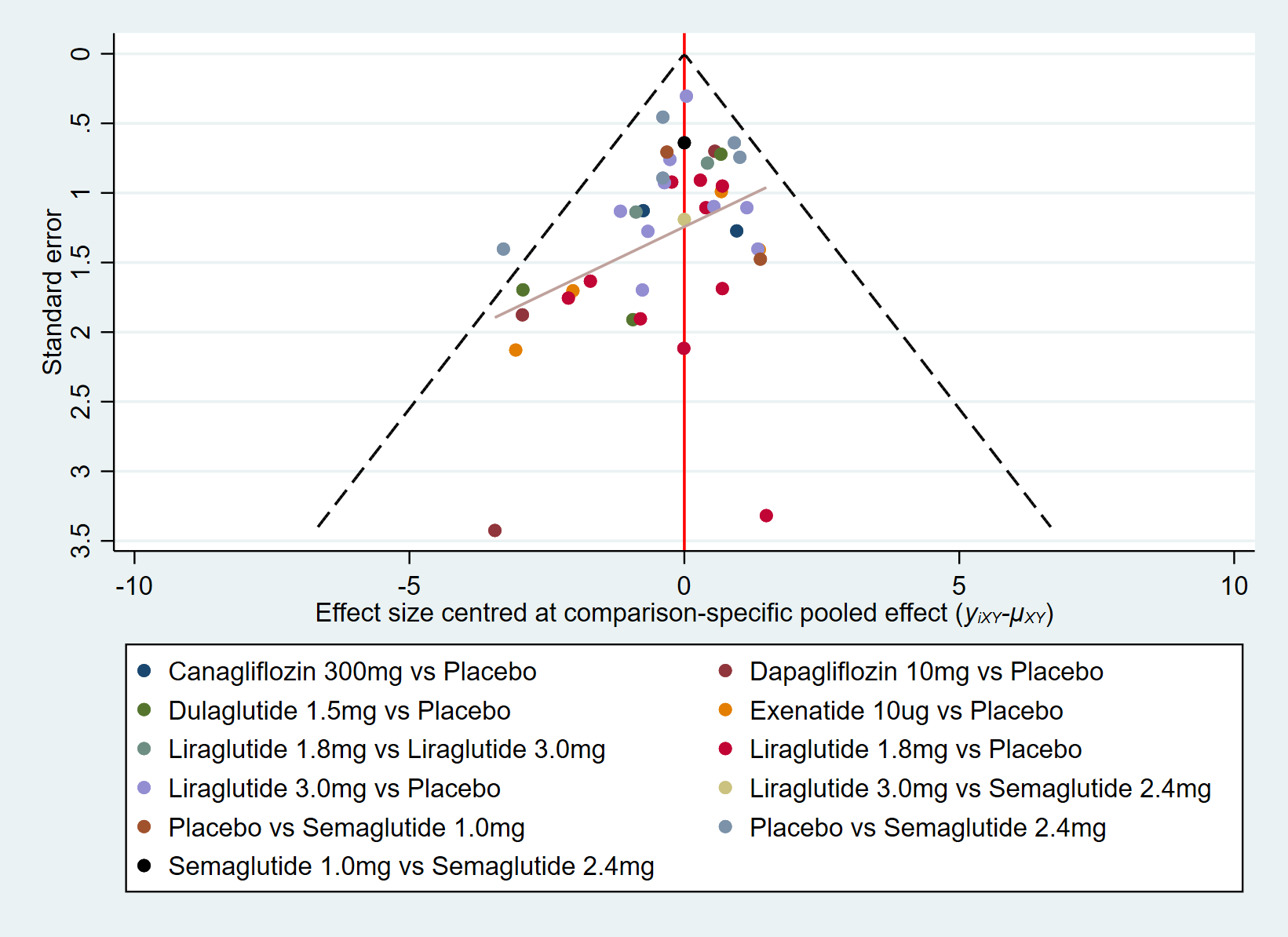

Supplement Figure 1E. Systolic blood pressure (SBP).

Supplement Figure 1F. Diastolic blood pressure (DBP).

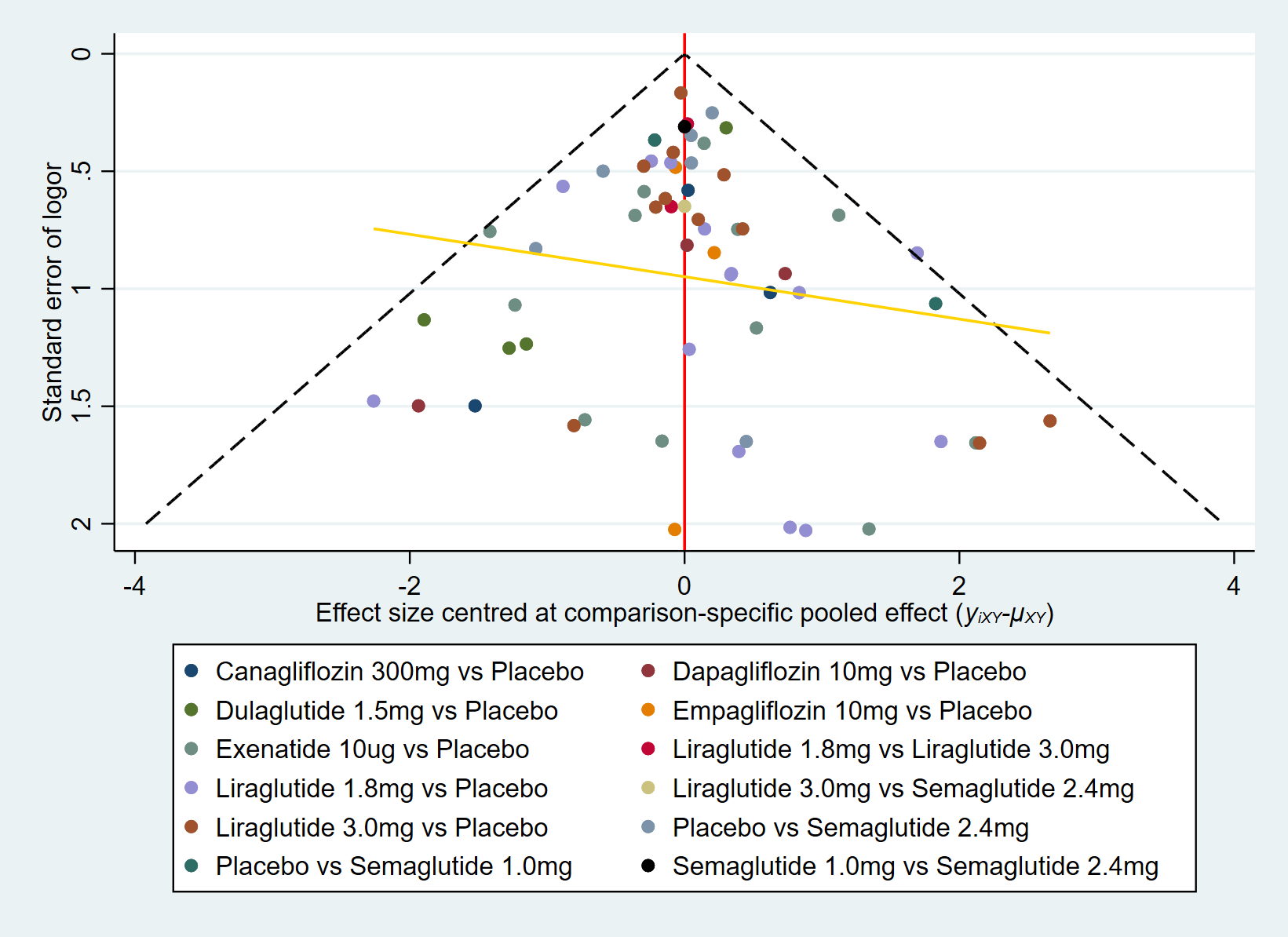
Supplement Figure 1G. Serious adverse events (SAE).

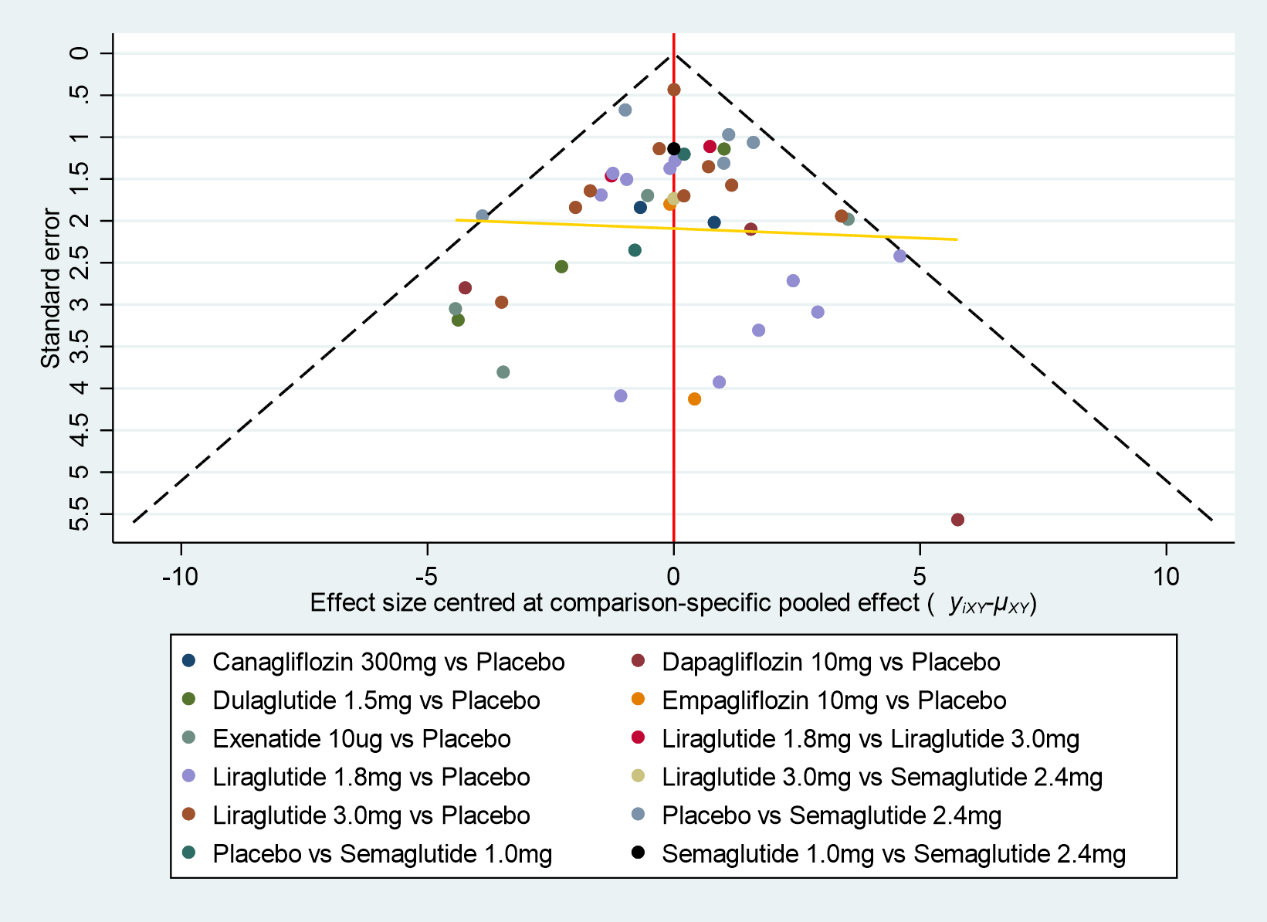

Supplement Figure 1H. Discontinue due to adverse events.

### Supplement Figure 2. Direct meta-analysis of different interventions.

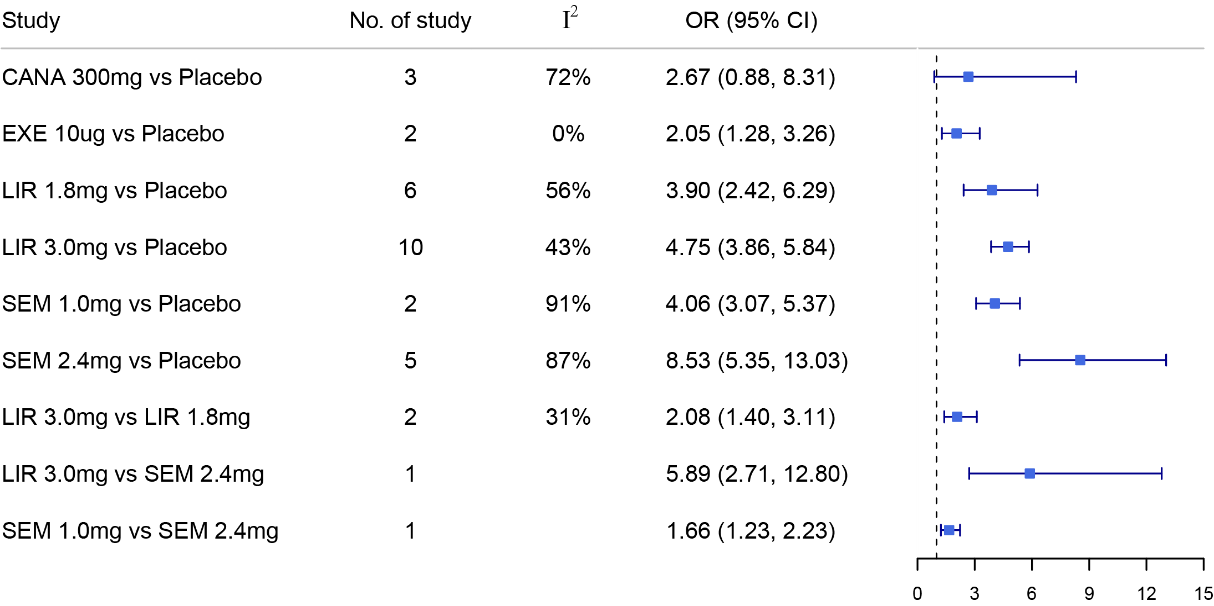

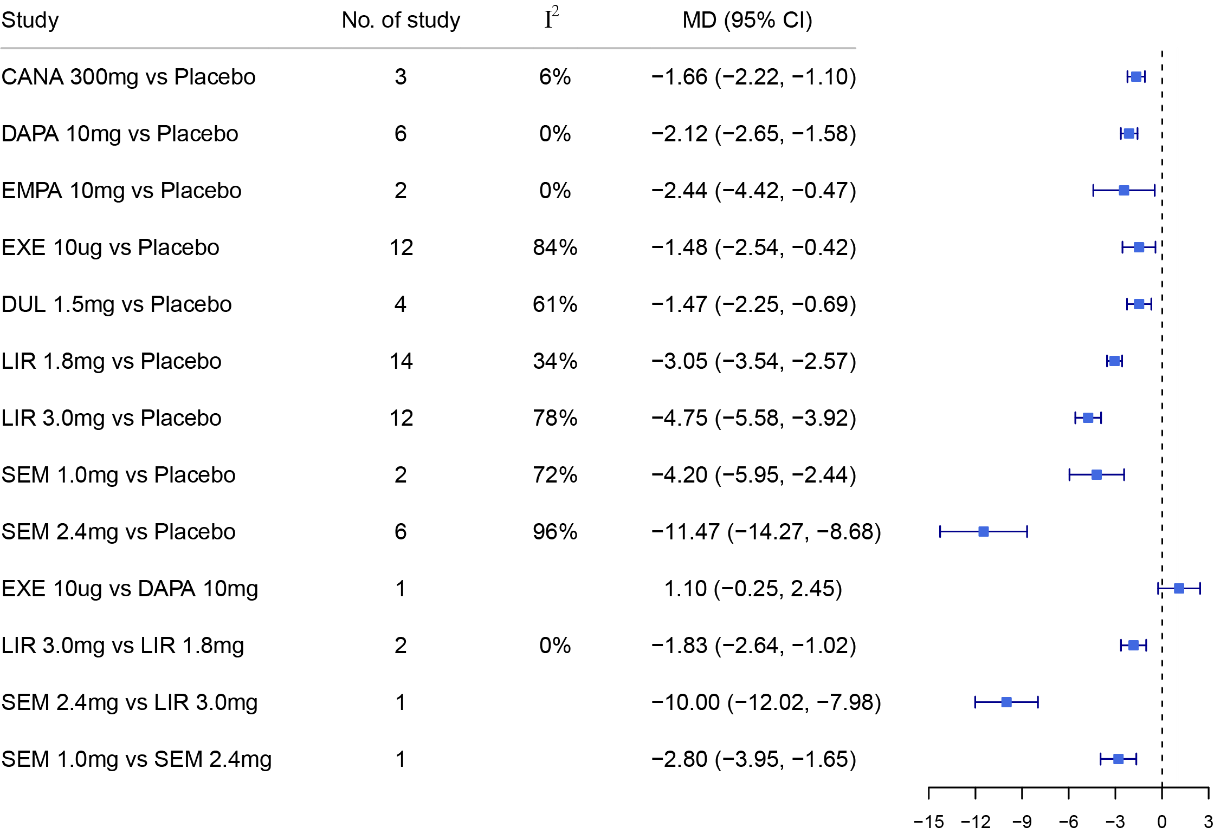
Supplement Figure 2A. Body weight (kg).

Supplement Figure 2B. ≥5% Weight loss (%).

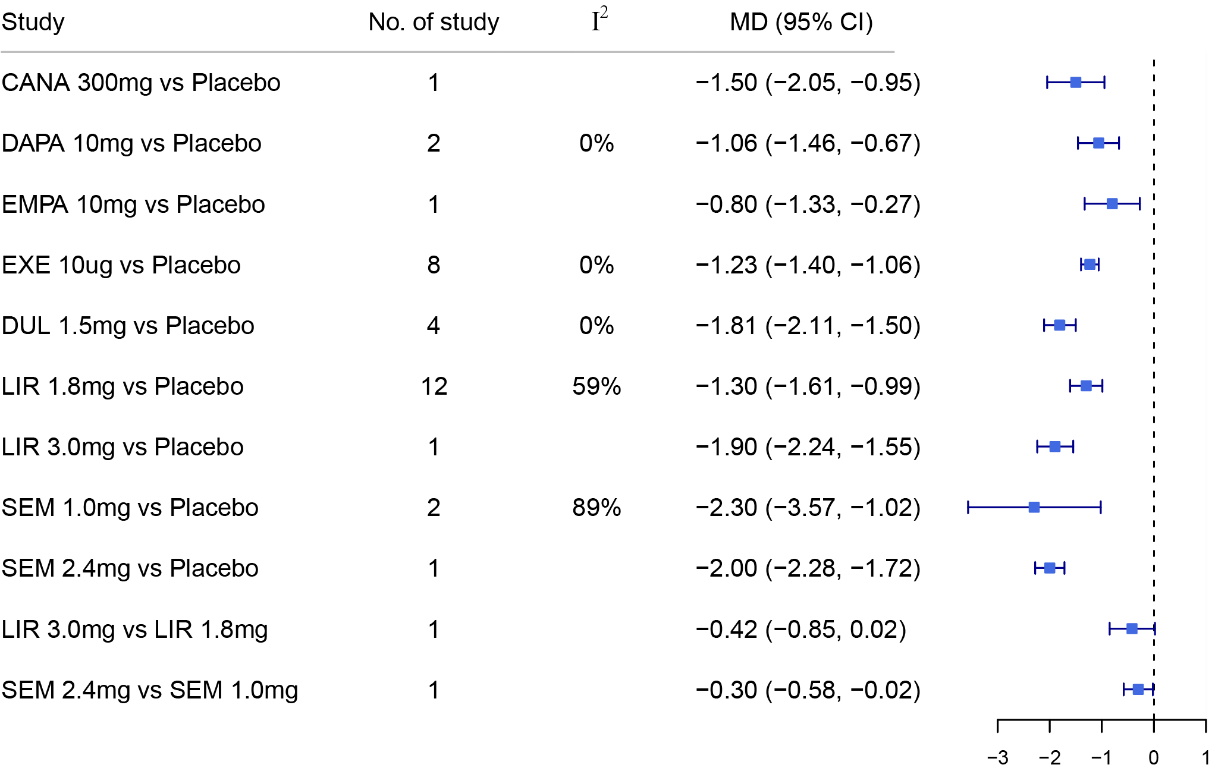

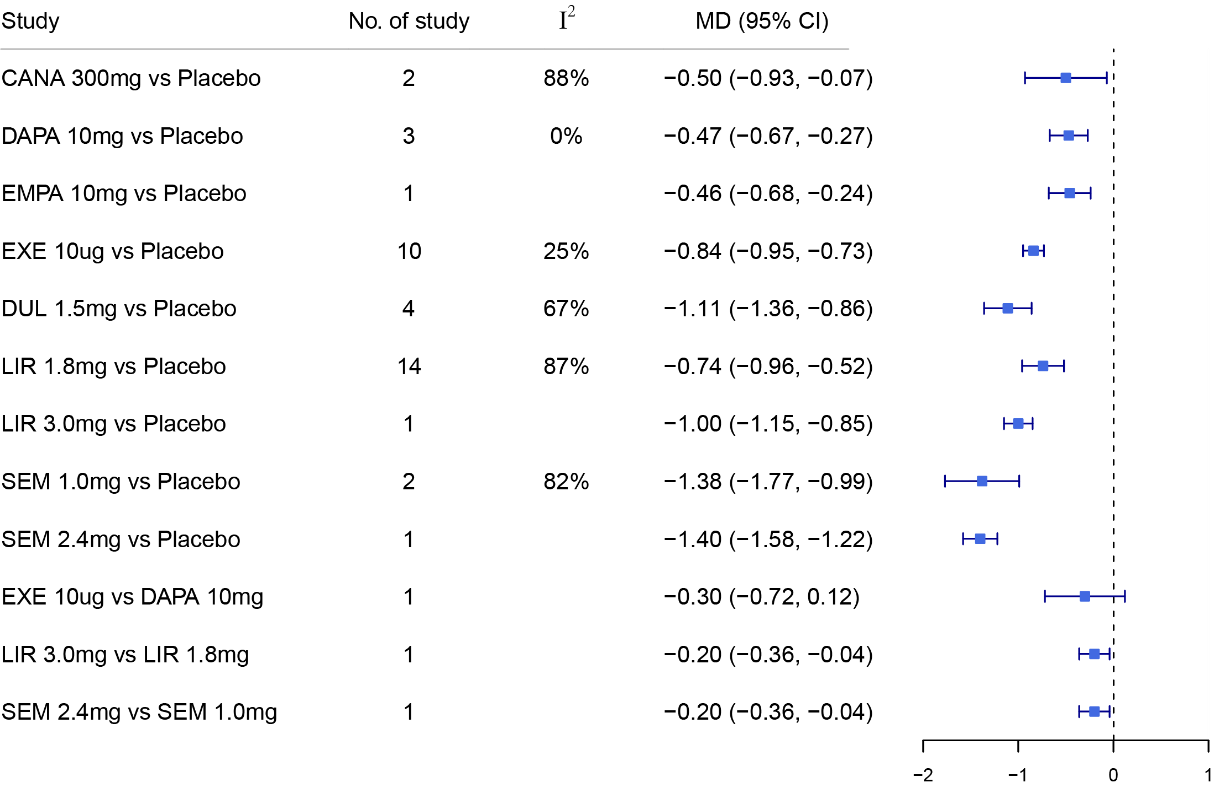

Supplement Figure 2D. Fasting plasma glucose (mmol/L).

Supplement Figure 2C. HbA1c (%).

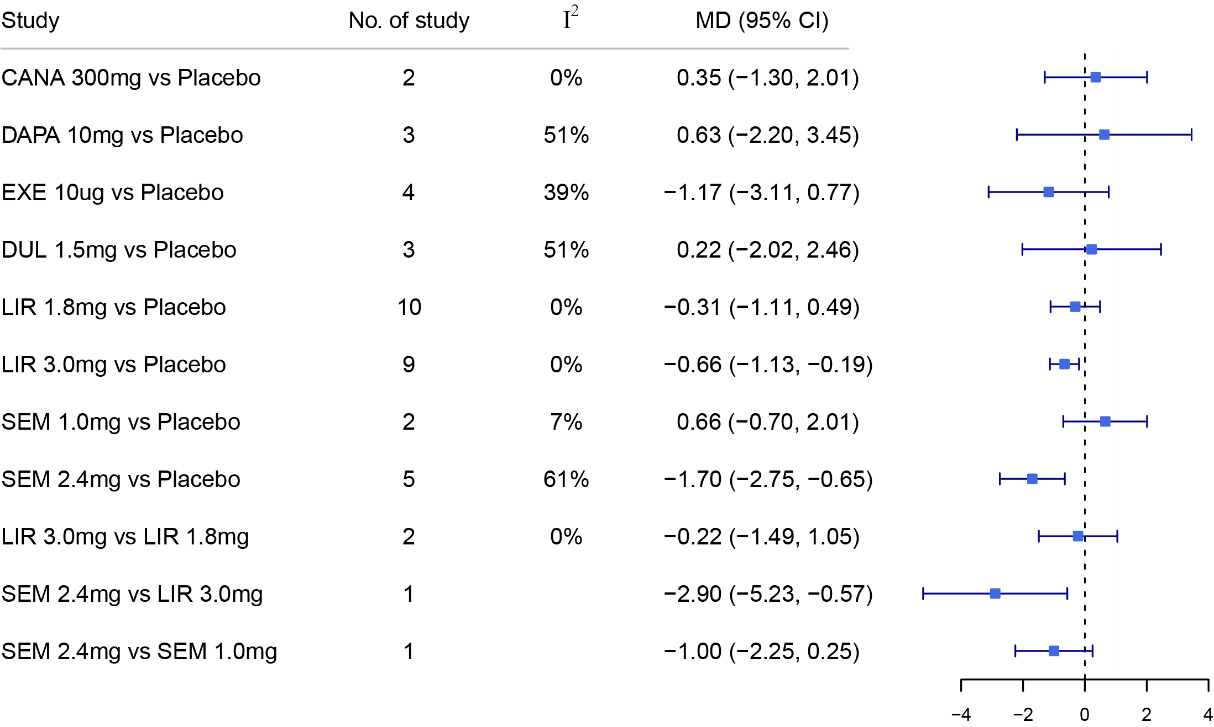

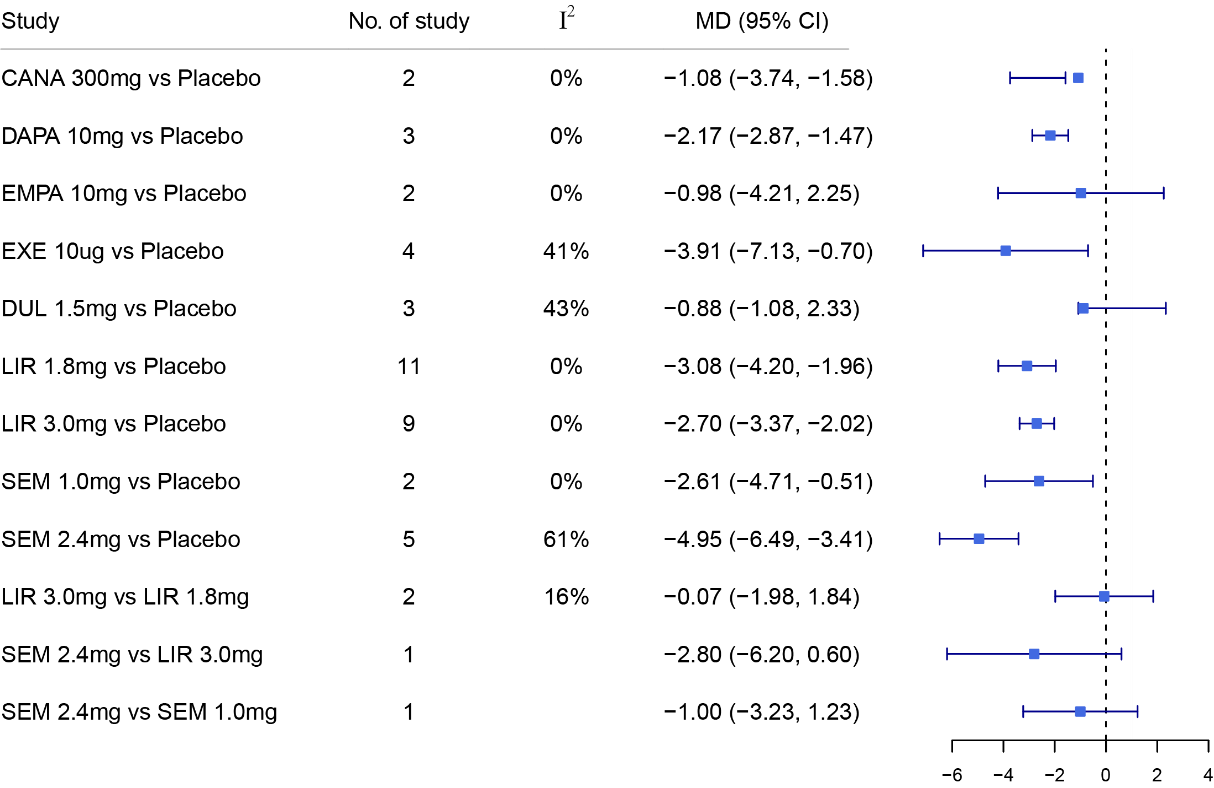

Supplement Figure 2F. Diastolic blood pressure (mmHg).

Supplement Figure 2E. Systolic blood pressure (mmHg).

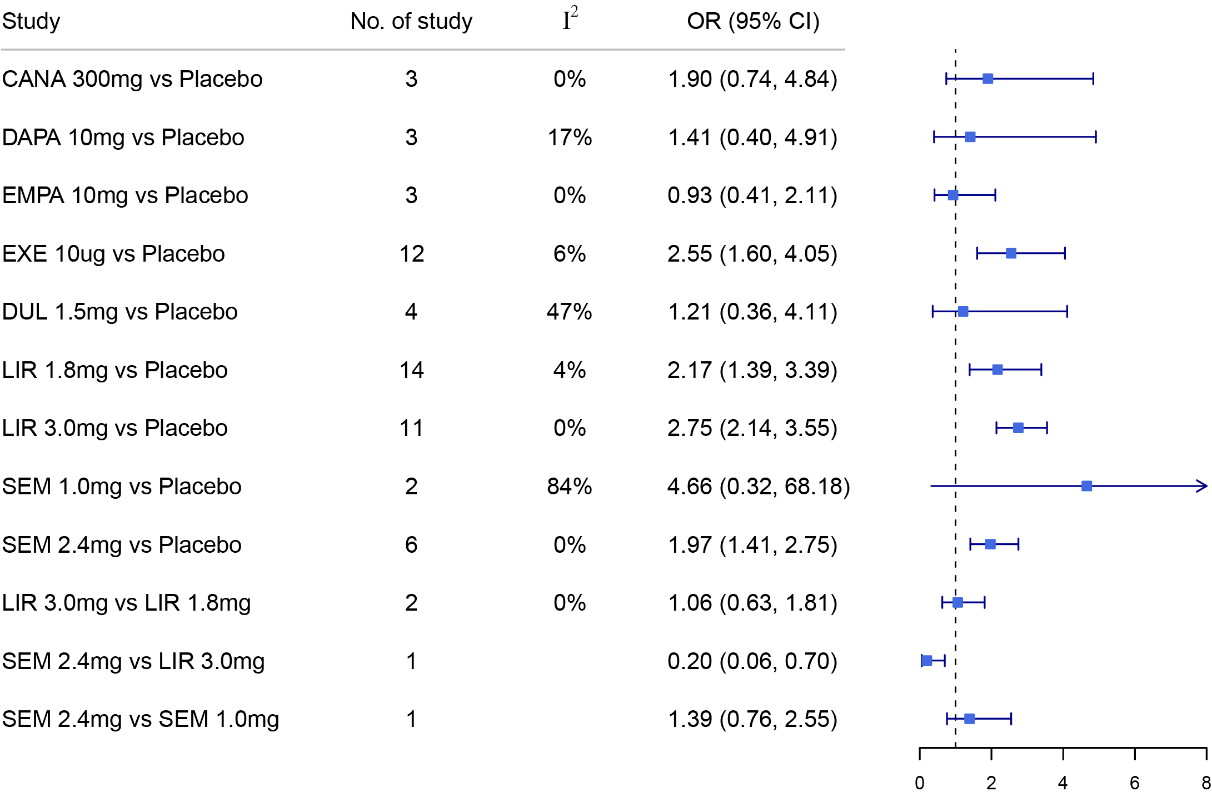

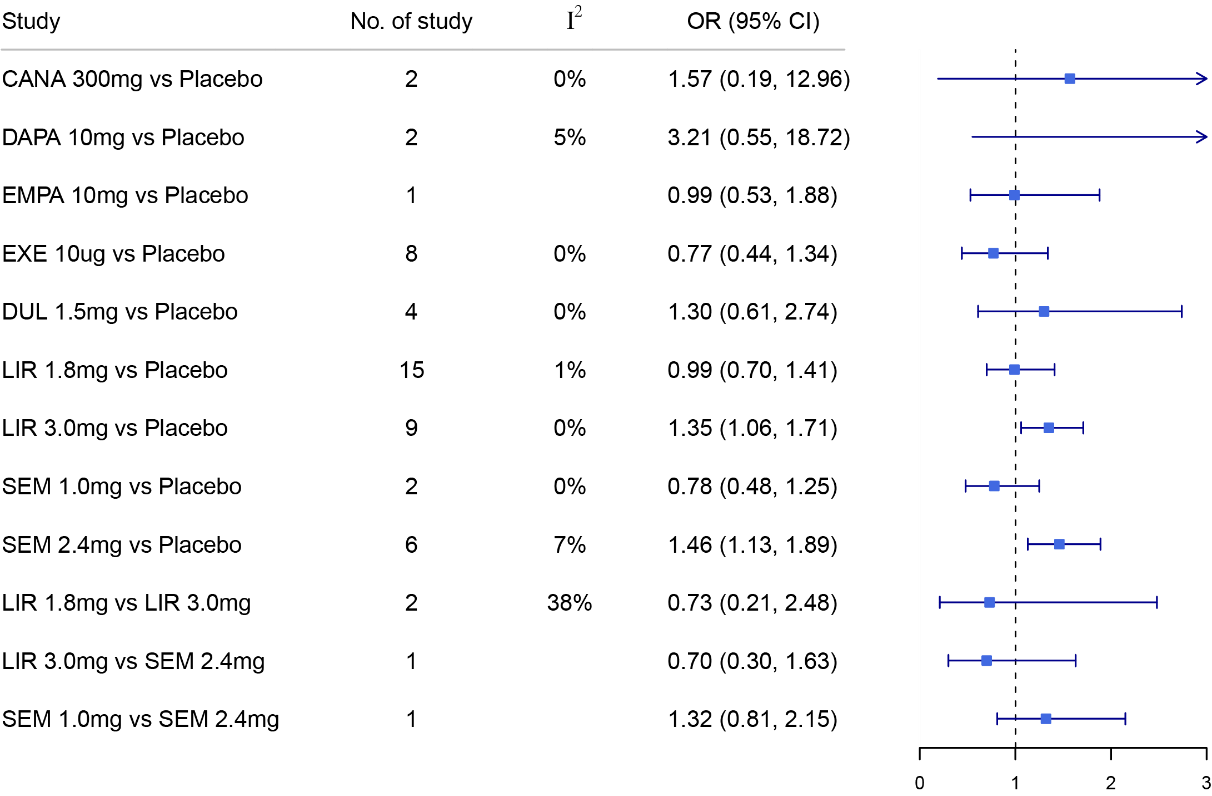

Supplement Figure 2H. discontinue due to adverse events.

Supplement Figure 2G. Serious adverse evens.

### Supplement Figure 3. Network meta-analysis of different interventions compared with placebo.

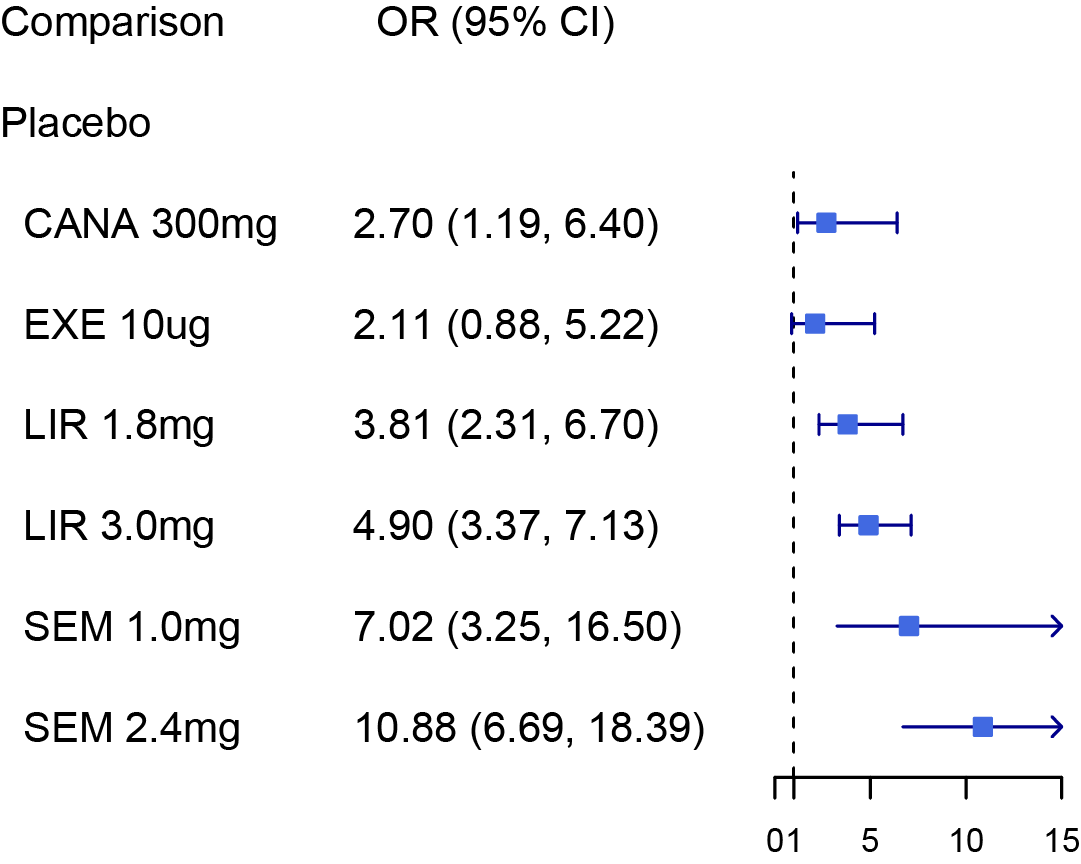

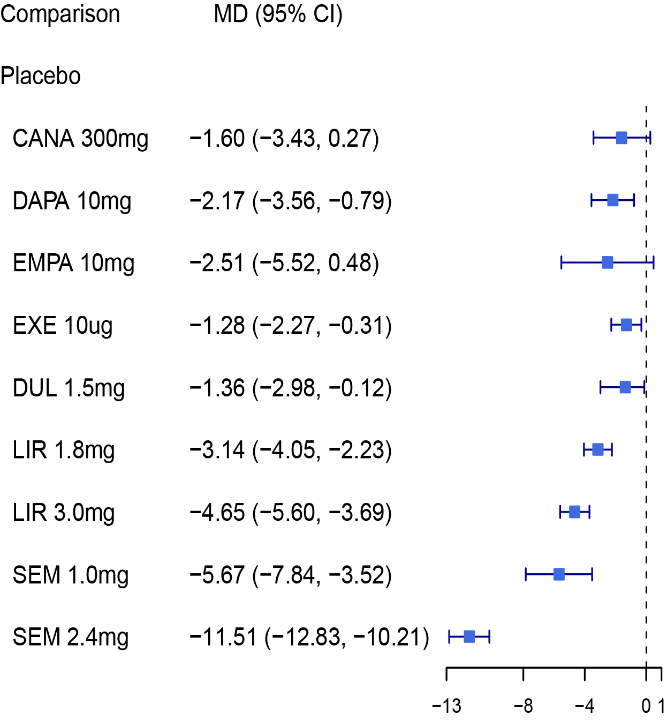

Achieving at least 5% weight loss

Body weight

Fasting plasma glucose (FPG)

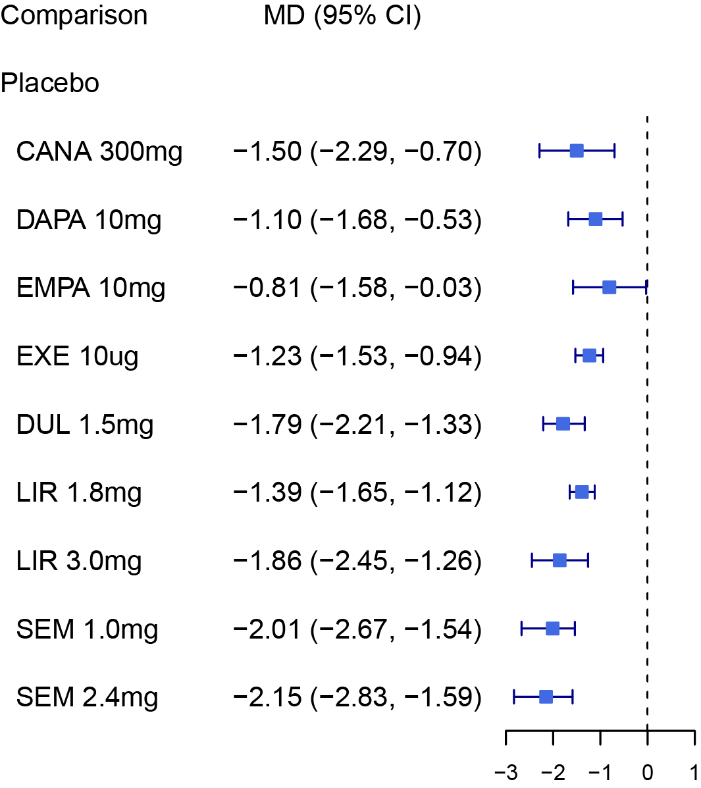

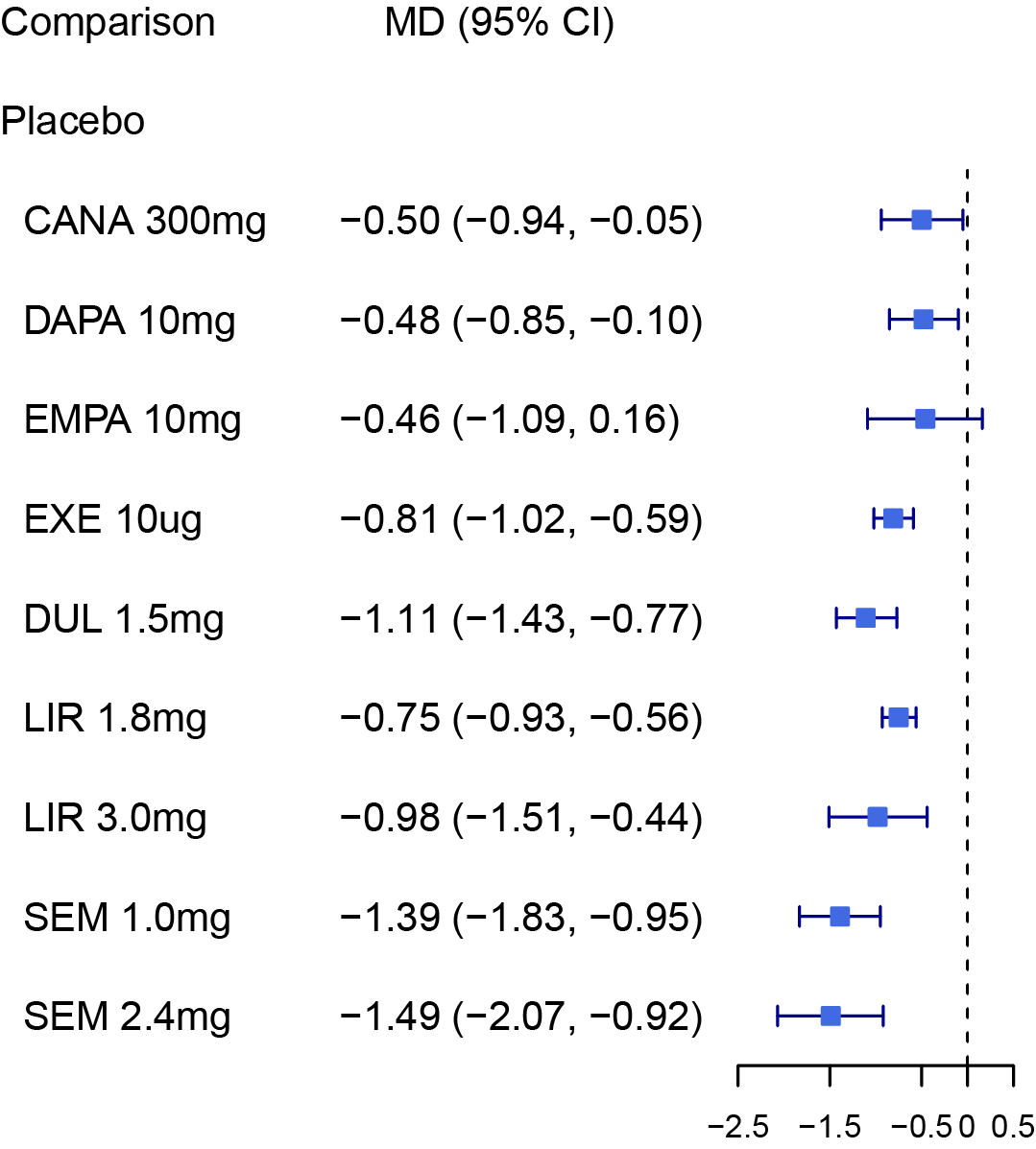

HbA1c

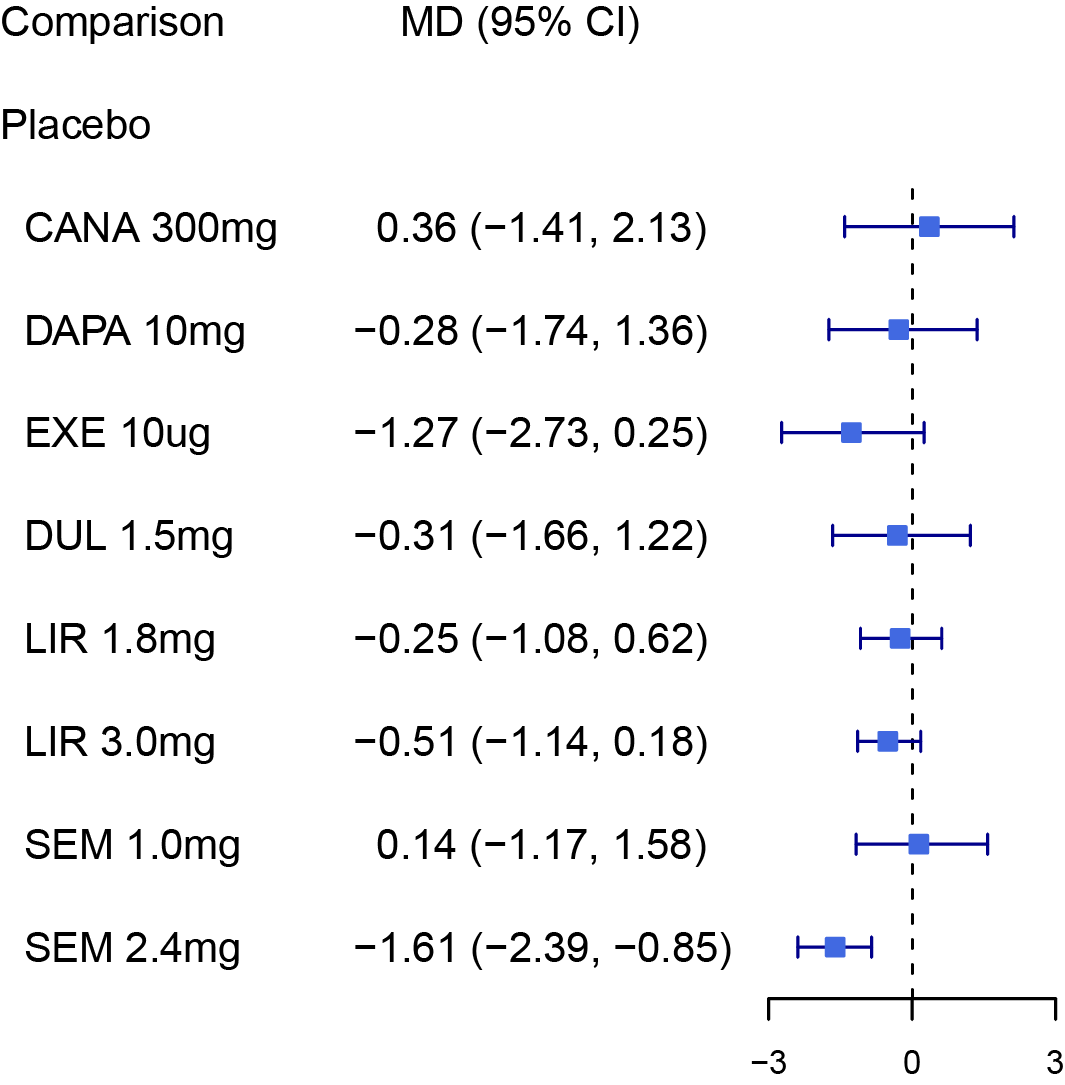

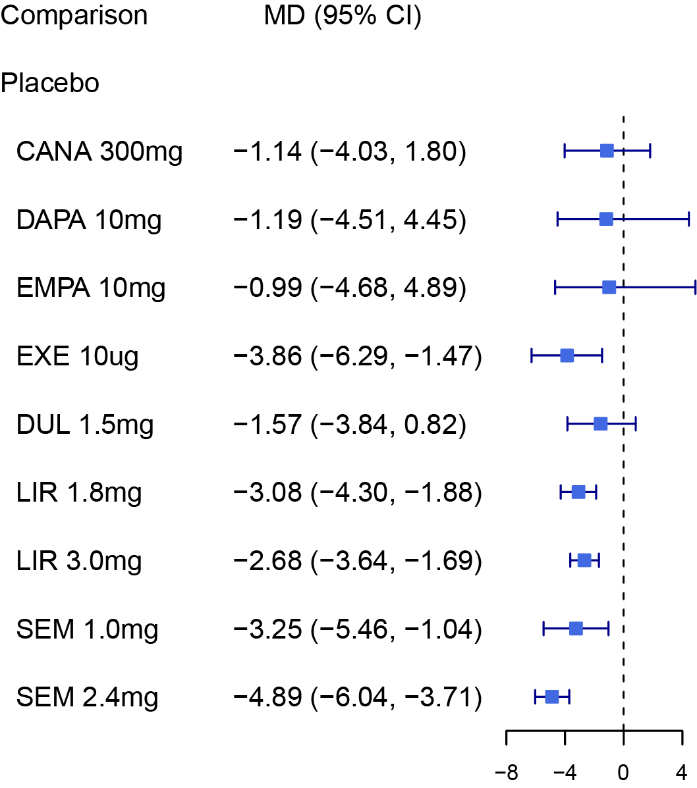

Diastolic blood pressure (DBP)

Systolic blood pressure (SBP)

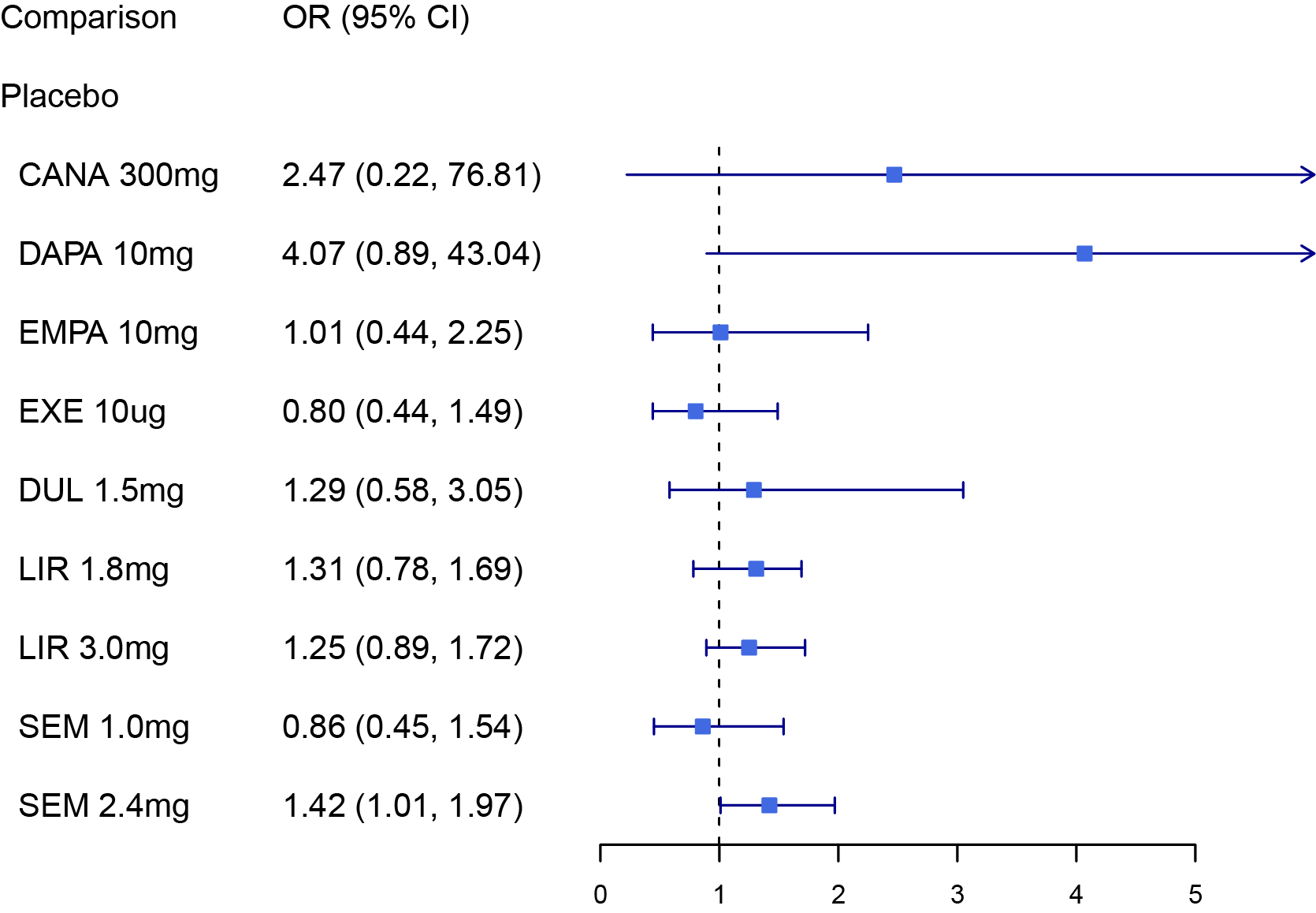

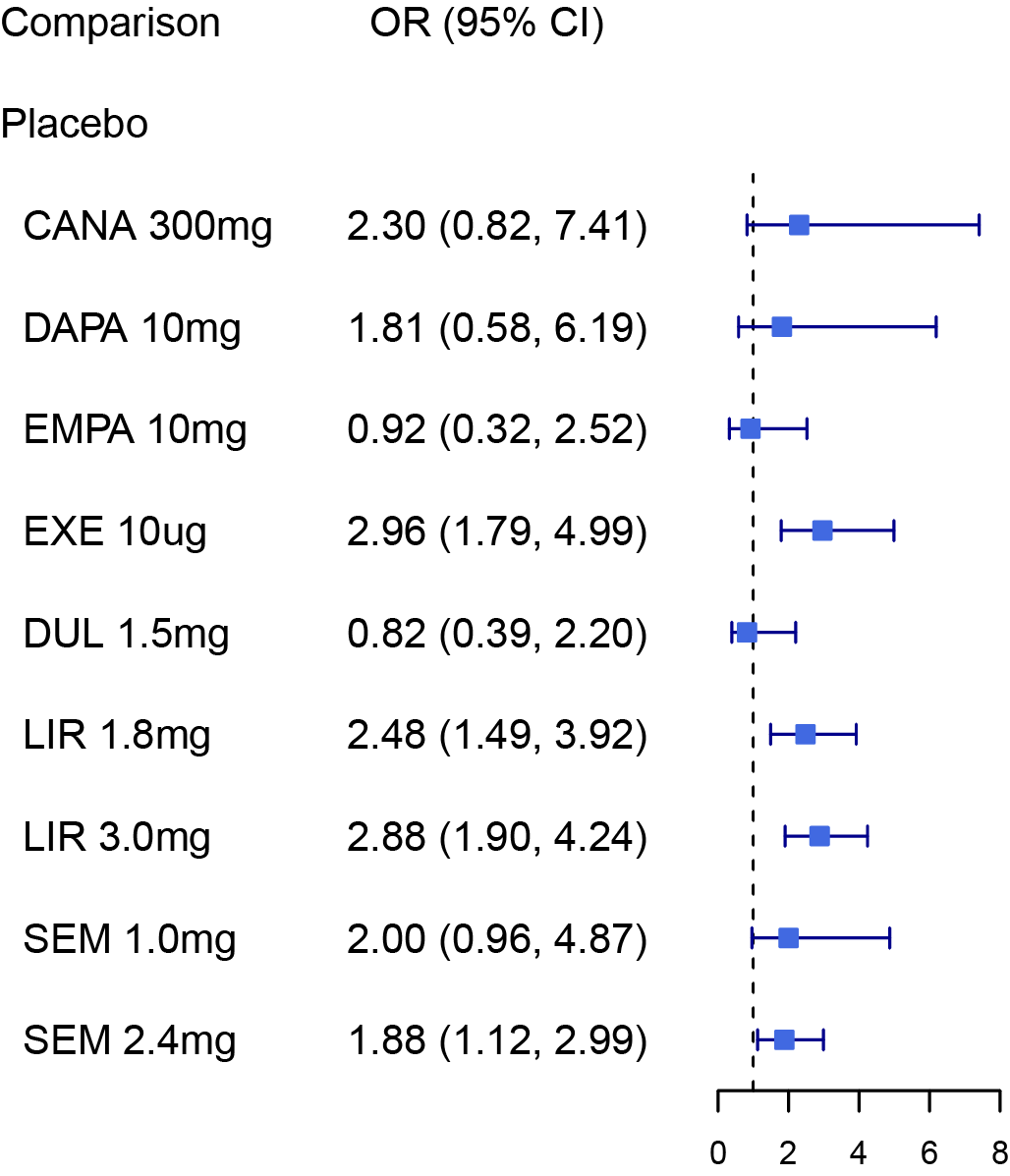

Discontinue due to adverse events

Serious adverse events (SAE)
